# Federated analysis of incubation period distributions using individual-level observed data and heterogeneous summary statistics

**DOI:** 10.64898/2026.06.01.26354607

**Authors:** Christian Morgenstern, Mark P. Khurana, Tristan Naidoo, Thom Rawson, Anne Cori, David A Duchene, Neil Ferguson, Moritz U.G. Kraemer, Samir Bhatt

## Abstract

The incubation period, the interval between pathogen exposure and symptom onset, is a critical epidemiological parameter for follow-up policy and outbreak response, yet individual-level exposure data remain scarce, especially early in outbreaks. For most priority pathogens, only summary statistics are available because sharing of individual-level data can be sensitive. Here we introduce a Bayesian hierarchical framework that jointly models individual-level observations and published summary statistics under a unified federated analysis framework. Simulation studies demonstrate that the method accurately recovers incubation period distributions across a range of data availability scenarios, generally outperforming approaches that use published summary statistics alone. Applying the framework to 18 pathogens, including 10 priority pathogens classified to have outbreak potential by the World Health Organization, we find substantial between-study heterogeneity in incubation period estimates, including by outbreak country for SARS-CoV-1, variants of concern for COVID-19, and exposure setting for typhoid fever. These estimates, together with the curated dataset and modelling framework in our associated R package `ddsynth`, provide a reproducible foundation for improved incubation period estimation and synthesis across pathogens of epidemic concern. Our framework enables robust and rapid estimation of incubation periods during new outbreaks.

## Introduction

The incubation period, the time from infection to symptom onset, is a fundamental epidemiological parameter and used extensively to chart the natural history of an infection or outbreak. The incubation period determines the duration of quarantine and active monitoring required to interrupt chains of transmission, informs the design of surveillance systems and contact tracing protocols, and governs the expected lag between an exposure event and its clinical manifestation in a population, as has been established across respiratory[1] and enteric pathogens[2]. These operational decisions depend not only on the mean incubation period but on the full probability distribution: for example, the proportion of cases exceeding a given quarantine window (the exceedance probability) determines the residual transmission risk that isolation policies leave unaddressed, the mass in the left tail governs how quickly exposed contacts become infectious before they can be identified, and the relationship between the incubation period and the generation time shapes whether non-pharmaceutical interventions can render an outbreak controllable [3–5]. Characterising the incubation period distribution accurately is therefore an integral component for any outbreak response.

Despite this, incubation period data are often not reported in a form that makes full distributional inference straightforward. The primary literature presents these data in a wide variety of formats, including individual-level exposure-onset records, grouped frequency tables, sample summary statistics such as means and percentiles, and fitted distributional parameters from data that are not publicly available [6]. This heterogeneity reflects persistent structural constraints: privacy regulations prevent public health authorities from sharing individual-level records, reporting standards in the primary literature remain inconsistent [7], and much of the evidence base rests on older studies, including human challenge experiments that may not reflect natural exposure in contemporary populations. While large prospective studies can provide precise estimates for a single pathogen, no single study captures the full range of variation across host populations, transmission routes, and pathogen variants, making cross-study synthesis necessary, albeit non-trivial.

Statistical meta-analysis addresses this by combining or pooling effect estimates across studies to produce an overall estimate [8, 9], while also quantifying between-study variability and uncertainty around the estimated average effect. Meta-analyses, however, typically discard much of the *within*-study distributional information and require study-level summaries to be converted to a common effect-size scale before pooling. Methods based on parametric survival analysis [1] and Bayesian inference [10] recover richer distributional information but are restricted to settings where individual-level data are available, and cannot incorporate the summary statistics that constitute the majority of the published evidence base. No existing approach jointly and exactly synthesises across the full range of data types present in the literature.

For most pathogens studied here, summary statistics are more commonly reported than individual-level records, and for several, including Nipah and MERS, individual-level data from publicly available sources alone would be insufficient to support between-study heterogeneity estimation.

Here we describe a Bayesian hierarchical framework for the joint inference of incubation period distributions from heterogeneous, distributed data. The published incubation-period evidence base is already decentralised, since raw observations typically remain with the original data holders for privacy and governance reasons, and what circulates are instead study-specific representations of those data, ranging from summary statistics such as means with standard deviations, medians with interquartile ranges or ranges, and frequency tables, up to and including individual exposure-onset records. We adopt the term ‘federated analysis’ to describe inference over a shared statistical model that consumes these heterogeneous representations directly, without requiring harmonisation to a common format. This mirrors the term’s use in multi-site clinical research [11, 12] and in proposed paradigms for distributed epidemic inference [13], but extends it from settings where every site shares the same type of derived quantity to settings where the form of the shared representation varies fundamentally between studies.

Conceptually, each study contributes the exact likelihood of its available representation under a parametric incubation-period distribution with study-specific parameters, themselves drawn from a pathogen-level distribution that captures between-study heterogeneity. Individual interval-censored exposure-onset pairs, frequency tables, and summary statistics therefore enter a single Bayesian hierarchical posterior. The framework supports five parametric families, including the commonly used log-normal, gamma and Weibull families and the more flexible Burr Type XII [14, 15] and generalised gamma distributions that improve characterisation of tail behaviour and outbreak-relevant exceedance risk. By construction, it decomposes apparent variability in reported incubation periods into genuine between-population variation and information loss due to study design and reporting format, and yields posterior estimates of policy-relevant quantities such as the predictive median, the 95th percentile and exceedance probabilities with calibrated uncertainty.

We validate our approach on simulated data with a known data-generating process and then apply it to data compiled from systematic reviews covering 18 epidemic and pandemic pathogens (Figure 1 and S2). These span viral haemorrhagic fevers, pandemic respiratory viruses, arboviral and vector-borne diseases, and environmental and zoonotic bacterial infections (Figure 1) [1, 2, 8, 9, 16–22]. For each pathogen, we infer the full posterior incubation period distributions and their between-study heterogeneity structure.

**Figure 1.**
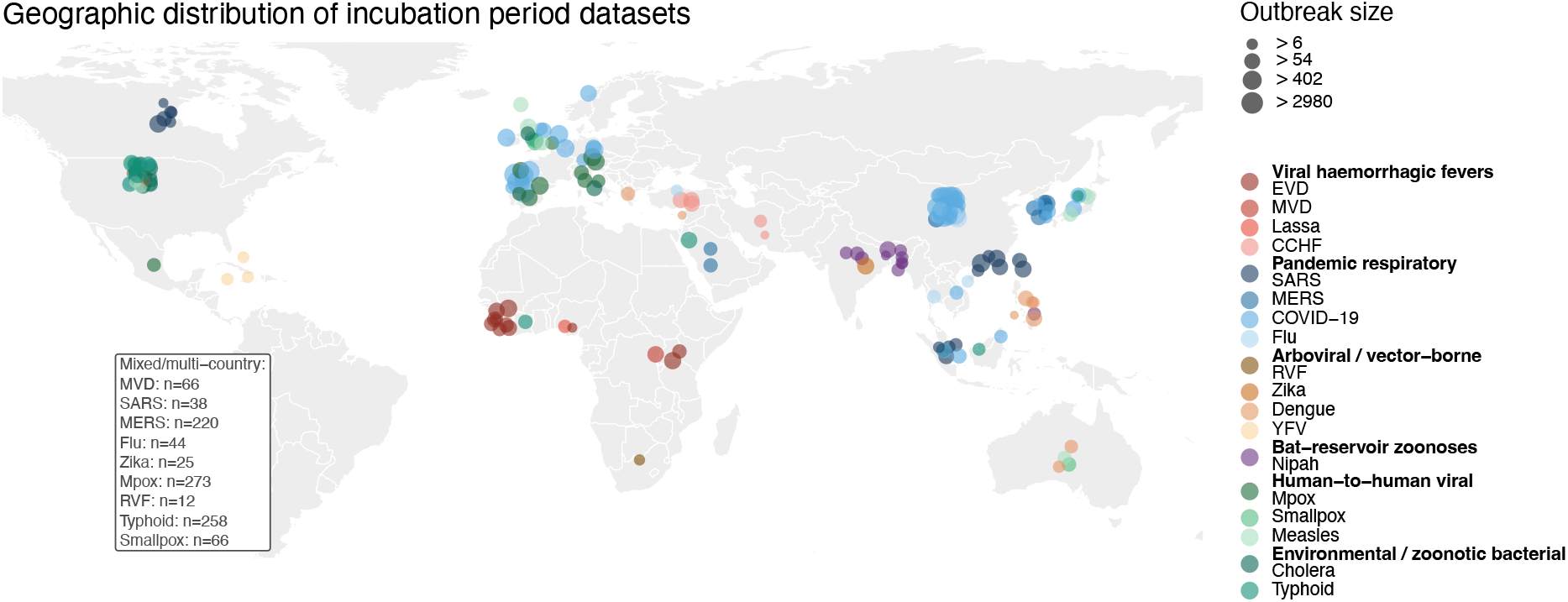
Each point represents one dataset, positioned at the centroid of the outbreak country and jittered to avoid overlap. Point size is proportional to the within-study sample size. Point colour denotes pathogen, grouped by epidemiological category as shown in the legend. Datasets from studies spanning multiple countries or with no single assignable location could not be placed geographically and are listed in the inset with their total sample size (*n*). In total, 225 datasets from 18 pathogens are included.

## Results

### Simulation study results

To evaluate model performance under realistic data conditions, we conducted a simulation study spanning five parametric families (log-normal, gamma, Weibull, Burr XII, and generalised gamma), four summary-statistic types (median + range, median + IQR, mean + SD, and frequency table), and a mixed-type condition in which different summary formats were available across studies. For each combination, we varied the number of contributing datasets (1–30+) and the within-study sample size (5–25+ observations), yielding 237 scenarios each evaluated over 100 simulation replicates. In each replicate, individual-level delay data were generated from a hierarchical model with known population parameters (*μ*_0_, *τ*, *ϕ* and, for the three-parameter families, an additional shape parameter *κ*), compressed to the relevant summary statistics, and analysed with the proposed model. Performance was assessed using 95% credible-interval coverage and median signed bias for all model parameters, and for the posterior predictive median (P50) and 95th percentile (P95) as the primary policy-relevant quantities, alongside the Weighted Interval Score (WIS) as a proper scoring rule for the full predictive distribution (Supplementary Information A.2.1). The coverage metric assesses how well the method is able to capture the true parameter within the credible interval and the bias metric of how far the estimate is away from the true parameter value.

The model showed well-calibrated predictive performance across most settings (Figure 2). Coverage of the predictive median was close to the nominal 95% level for all five distribution families and all summary-statistic types, with the greatest departures seen in the sparsest data settings (<10 datasets, ≤5 observations per study; Figure 2A). Bias in the predictive median was negligible across conditions (Figure 2A). Coverage of the 95th percentile was slightly more variable, with the most notable departures occurring in scenarios with only a single contributing dataset, where insufficient data precluded reliable estimation of the upper tail regardless of summary-statistic type (Figure 2B). Bias in the 95th percentile was generally modest and positive, consistent with a small tendency to overestimate the tail in data-sparse conditions (Figure 2B). Predictive accuracy as measured by the mean WIS, expressed in days and jointly penalising miscalibration and lack of sharpness, was broadly consistent across distribution families, with the notable exception of Burr Type XII, where three distinct performance clusters are visible (Supplementary Figure S3C). These clusters reflect the three values of the tail-heaviness parameter *κ* (2, 3, and 5) used in the Burr Type XII scenarios. Lower *κ* produces heavier-tailed distributions with wider predictive intervals and consequently higher WIS, while higher *κ* yields lighter tails and lower WIS. That the model accurately recovers the predictive distribution across all three *κ* values demonstrates its ability to capture substantial variation in tail behaviour; the lower WIS for higher *κ* reflects the intrinsically narrower predictive intervals of lighter-tailed distributions rather than differential model performance.

**Figure 2.**
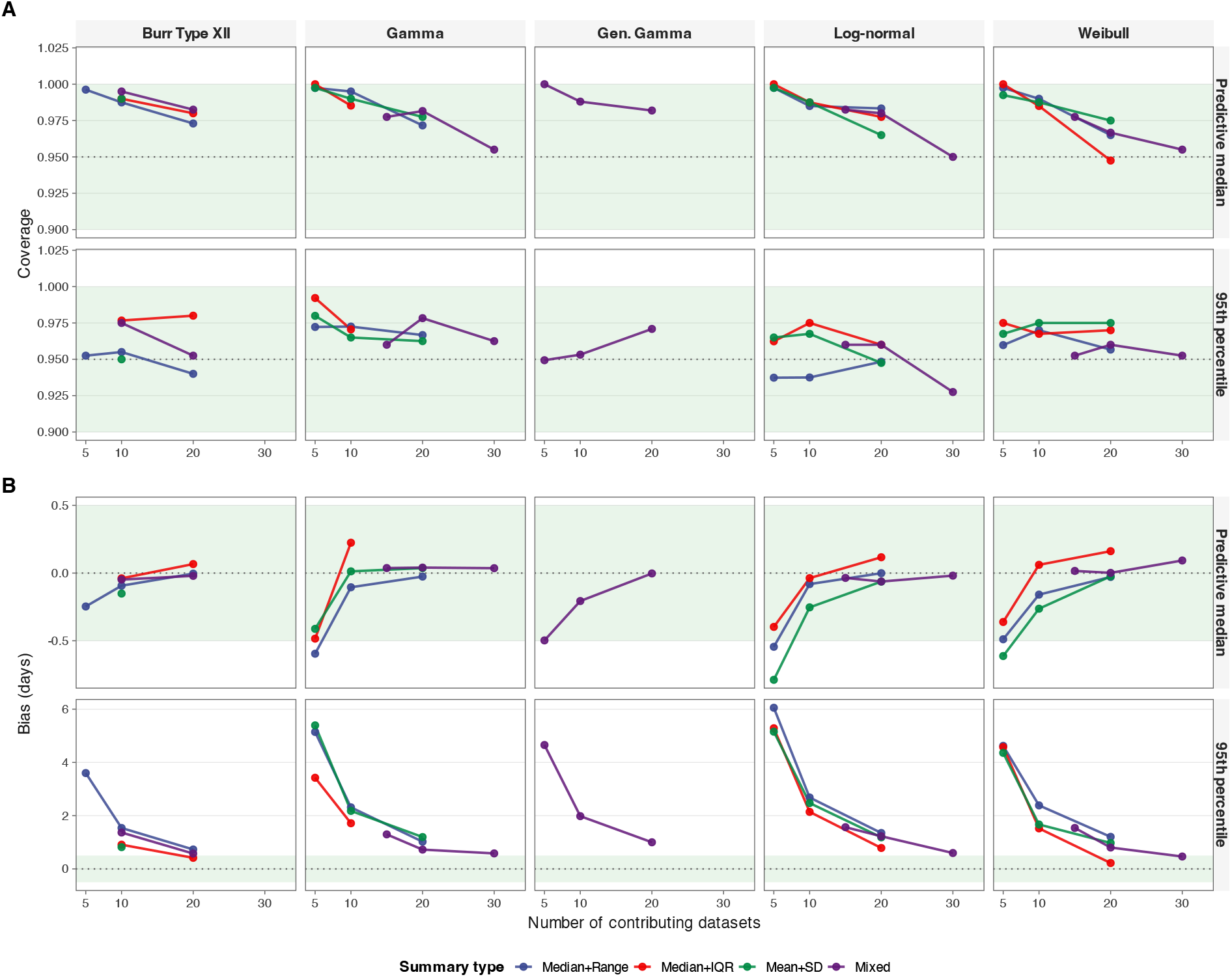
Simulation study predictive performance as a function of the number of contributing datasets. Results are averaged across within-study sample size and distributional sub-scenarios. Columns correspond to the five parametric families evaluated (Burr Type XII, gamma, generalised gamma, log-normal, Weibull); line colour indicates summary-statistic type, as shown in the legend. The green shaded region marks the acceptable zone (coverage: 0.90–1.00; bias: −0.5 to 0.5 days). The dotted line indicates the nominal target (0.95 for coverage panels; 0 for bias panels). Frequency-table summary type excluded; generalised gamma scenarios only available for the mixed summary-statistic type. (A) Empirical 95% credible interval coverage for the predictive median (P50; top row) and 95th percentile (P95; bottom row). (B) Median signed bias (days) for the predictive median (P50; top row) and 95th percentile (P95; bottom row).

Sensitivity of results to the ground truth values of *τ* (between-study heterogeneity) and *μ*_0_ (population mean log-delay) used to generate the simulated data is presented in Supplementary Figure S4. Coverage of the predictive median remained close to the nominal 95% level across all five distribution families and all parameter configurations. Coverage of the 95th percentile was broadly maintained but showed more variability across conditions, with coverage tending to be higher for larger *μ*_0_ and greater between-study heterogeneity. WIS was lower when the true delay was short and between-study heterogeneity was small, reflecting that tighter, shorter distributions are inherently easier to predict precisely; performance remained robust across all ground truth configurations.

### Collected data across pathogen groups

We compiled incubation period data from published epidemiological studies for 18 pathogens, yielding 225 datasets in total (Supplementary Table S6). For WHO priority pathogens [23], we identified relevant studies through systematic reviews conducted by the Pathogen Epidemiology Review Group (PERG) at Imperial College London, then returned to the underlying primary studies to extract data directly. For remaining pathogens we drew on available systematic reviews or, where none existed, conducted a PubMed search for “incubation period” combined with the pathogen name (typhoid fever, smallpox). In each case, if a study reported individual-level observations we encoded those preferentially; if only summary statistics were available we encoded those instead. Where a study reported only fitted distributional parameters, we searched the broader literature for the underlying individual-level data and encoded those directly, discarding the authors’ derived estimates in favour of the raw observations. Data reported only in figures were extracted by digital image processing. The 18 pathogens covered a broad range of transmission routes and disease severity (Supplementary Figure S2), spanning six groups: viral haemorrhagic fevers (Ebola, Marburg, Lassa, and Crimean-Congo haemorrhagic fever); arboviral and vector-borne diseases (Rift Valley fever, Zika, dengue, and yellow fever); pandemic respiratory pathogens (SARS, MERS, COVID-19, and influenza A/B); the bat-reservoir zoonosis Nipah; (non-respiratory) human-to-human viral diseases (mpox, smallpox, and measles); and environmental or zoonotic bacterial infections (cholera and typhoid fever). The number of datasets per pathogen ranged from 1 (Lassa) to 51 (COVID-19). Studies reported results in five summary formats: frequency tables, including exact-value and interval-censored (*n*=99 datasets), median with range (*n*=67), mean with standard deviation (*n*=37), and median with interquartile range (22). Within-study sample sizes ranged from 4 to 2,907 (median 33).

### Inferred incubation period distribution

Figure 3 presents the posterior predictive distributions for all 18 pathogens. All five parametric families were fitted to each dataset; the distribution selected by leave-one-out cross-validation (LOO) is displayed, with parameter estimates for the best fitting model displayed in Table 1. The Burr Type XII distribution provided the best fit for the majority of pathogens, including Ebola, MVD (Marburg virus disease), CCHF (Crimean-Congo haemorrhagic fever), SARS, Nipah, smallpox, cholera, and typhoid fever, reflecting the heavy-tailed nature of incubation period distributions across many infectious diseases.

**Figure 3.**
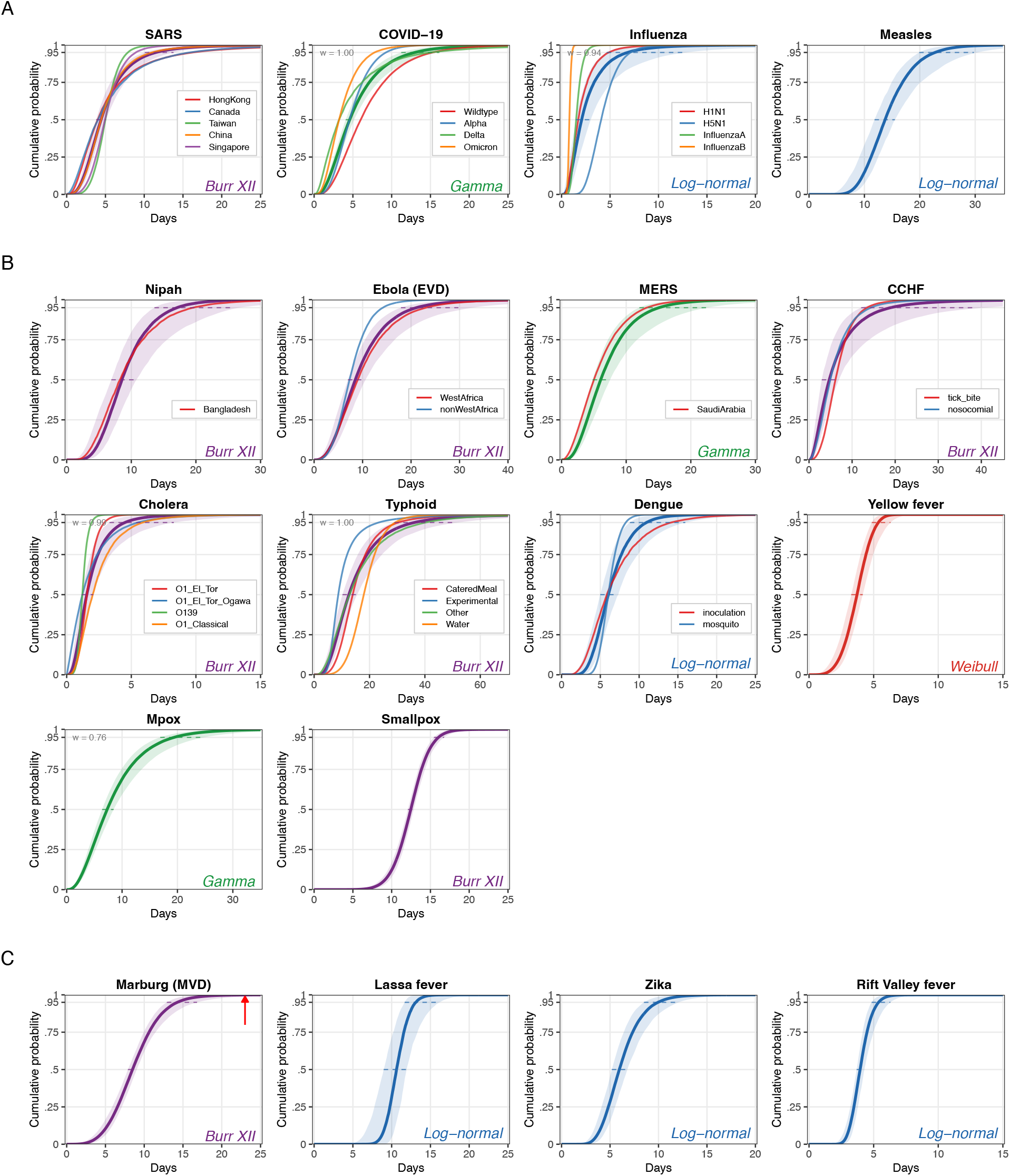
Posterior predictive incubation period distributions for 18 pathogens: All five parametric families (log-normal, gamma, Weibull, Burr XII, generalised gamma) were fitted to each pathogen; the best-fitting distribution according to leave-one-out cross-validation (LOO) is shown for each pathogen (the colour of the main inferred CDF and credible interval is given by the distribution). Solid lines represent the posterior median predictive cumulative distribution function (CDF); shaded ribbons represent the 95% credible interval. Where subgroup analyses were performed, each subgroup is shown as a separate CDF without credible intervals. Dashed lines indicate the 95% credible interval of the median and the 95th percentile. (A) Pathogens with the largest number of contributing datasets. (B) Pathogens with a moderate number of contributing datasets. (C) Data-sparse pathogens with few contributing studies. The red arrow in the Marburg panel indicates a case report describing a substantially longer incubation period that could not be incorporated into the analysis. Parameter estimates for the best-fitting model are reported in Table 1.

**Table 1.** Incubation period estimates by pathogen, grouped by pathogen type: Values are posterior medians with 95% credible intervals. The best-fitting parametric family, selected by leave-one-out cross-validation, is shown for each pathogen. *μ*_0_ is the population mean log-delay; *ϕ* is the within-study dispersion parameter; *κ* is the additional shape parameter estimated for Burr Type XII and generalised gamma distributions (shown as — for other families); *τ* is the between-study heterogeneity standard deviation (shown as — where less than 5 dataset were available). Subgroup analyses are shown in italics beneath each pathogen. The predictive median (days) and 95th percentile (P95, days) are derived from the posterior predictive distribution. For pathogens with five or more contributing datasets, between-study heterogeneity is propagated by sampling new study locations from 𝒩 (*μ*_0_,*τ*); for pathogens with fewer than five datasets, *τ* is not identifiable and predictions are computed at the mean of the observed study-level locations.

| Pathogen | Distribution | Median (days) | P95 (days) | $\mu_0$ | $\phi$ | $\kappa$ | $\tau$ |
| --- | --- | --- | --- | --- | --- | --- | --- |
| ***Viral haemorrhagic fevers*** |  |  |  |  |  |  |  |
| **Ebola (EVD)** | Burr XII | 8.9 (7.4, 11.4) | 19.5 (16.2, 24.9) | 2.51 (2.20, 2.92) | 2.67 (2.28, 3.18) | 2.14 (1.37, 3.98) | 0.28 (0.16, 0.55) |
| *West Africa* |  | 9.5 (7.6, 13.6) | 21.1 (16.8, 30.0) | 2.62 (2.24, 3.08) | 2.57 (2.17, 3.11) | 2.30 (1.42, 4.47) | 0.31 (0.15, 0.69) |
| *non-West Africa* |  | 7.3 (6.2, 8.6) | 14.2 (11.9, 17.8) | 2.33 (1.32, 3.25) | 2.94 (2.30, 3.96) | 2.55 (1.24, 6.01) | — |
| **Marburg (MVD)** | Burr XII | 8.5 (8.0, 9.0) | 14.3 (12.8, 16.5) | 2.41 (1.18, 3.55) | 3.68 (2.88, 4.78) | 2.99 (1.33, 7.23) | — |
| **Lassa fever** | Log-normal | 10.7 (9.2, 12.0) | 13.2 (11.6, 16.4) | 2.36 (0.90, 3.79) | 0.13 (0.07, 0.25) | — | — |
| **CCHF** | Burr XII | 5.9 (4.0, 10.7) | 14.6 (9.9, 26.6) | 1.82 (1.30, 2.47) | 2.55 (1.95, 3.44) | 1.60 (0.95, 3.61) | 0.63 (0.41, 1.07) |
| *Tick-bite* |  | 6.2 (5.4, 7.1) | 13.6 (11.7, 16.2) | 2.21 (1.02, 3.42) | 2.75 (2.01, 4.50) | 1.95 (0.79, 5.44) | — |
| *Nosocomial* |  | 4.9 (3.4, 6.9) | 14.2 (9.7, 26.4) | 2.12 (1.00, 3.18) | 1.89 (1.29, 2.76) | 2.44 (1.06, 6.02) | — |
| ***Pandemic respiratory*** |  |  |  |  |  |  |  |
| **COVID-19** | Gamma | 5.1 (4.5, 5.9) | 10.7 (9.4, 12.4) | 1.62 (1.50, 1.75) | 4.02 (3.95, 4.09) | — | 0.42 (0.34, 0.54) |
| *Wildtype* |  | 6.4 (5.6, 7.4) | 13.0 (11.4, 15.1) | 1.86 (1.73, 2.00) | 4.44 (4.33, 4.55) | — | 0.36 (0.27, 0.49) |
| *Alpha* |  | 4.6 (4.3, 4.9) | 9.3 (8.7, 9.9) | 1.60 (0.47, 2.72) | 4.49 (4.32, 4.66) | — | — |
| *Delta* |  | 4.5 (2.7, 12.6) | 9.5 (5.7, 26.6) | 1.30 (0.69, 1.91) | 3.99 (3.84, 4.14) | — | 0.75 (0.43, 1.45) |
| *Omicron* |  | 3.4 (2.9, 4.4) | 7.1 (6.0, 9.3) | 1.27 (1.07, 1.47) | 4.04 (3.86, 4.23) | — | 0.27 (0.15, 0.56) |
| **SARS** | Burr XII | 4.6 (4.0, 5.4) | 10.7 (9.3, 12.5) | 1.58 (1.37, 1.87) | 3.15 (2.60, 3.80) | 1.22 (0.88, 1.94) | 0.27 (0.18, 0.42) |
| *Hong Kong* |  | 4.7 (2.9, 12.6) | 13.1 (8.2, 35.2) | 1.33 (0.75, 1.93) | 3.10 (2.48, 3.92) | 0.89 (0.66, 1.22) | 0.57 (0.27, 1.33) |
| *Canada* |  | 5.0 (3.2, 10.6) | 13.6 (8.6, 29.0) | 2.19 (1.44, 2.92) | 1.91 (1.60, 2.31) | 3.06 (1.52, 7.12) | 0.53 (0.26, 1.19) |
| *Taiwan* |  | 5.0 (4.7, 5.3) | 7.8 (7.0, 9.2) | 1.78 (0.56, 2.90) | 4.65 (3.29, 6.65) | 2.20 (1.09, 5.01) | — |
| *China* |  | 4.5 (4.0, 5.0) | 11.1 (10.0, 12.4) | 1.64 (0.71, 2.67) | 2.87 (1.88, 5.12) | 1.25 (0.58, 3.60) | — |
| *Singapore* |  | 4.9 (4.4, 5.5) | 8.9 (7.9, 10.2) | 1.84 (1.03, 2.64) | 3.62 (2.78, 5.31) | 2.04 (0.86, 5.63) | — |
| **MERS** | Gamma | 6.4 (5.2, 8.6) | 13.5 (11.0, 18.3) | 1.89 (1.66, 2.12) | 3.99 (3.38, 4.68) | — | 0.31 (0.17, 0.62) |
| *Saudi Arabia* |  | 5.0 (3.9, 6.1) | 13.1 (11.0, 16.4) | 1.76 (0.62, 2.89) | 2.24 (1.41, 3.43) | — | — |
| **Influenza** | Log-normal | 2.6 (1.8, 4.2) | 4.6 (3.2, 7.6) | 0.74 (0.38, 1.10) | 0.35 (0.33, 0.38) | — | 0.63 (0.43, 0.99) |
| *H1N1* |  | 1.9 (1.3, 4.9) | 3.6 (2.3, 9.1) | 0.53 (-0.01, 1.08) | 0.37 (0.33, 0.41) | — | 0.52 (0.24, 1.24) |
| *H5N1* |  | 4.1 (3.7, 4.5) | 7.2 (6.3, 8.5) | 1.41 (0.67, 2.13) | 0.34 (0.27, 0.43) | — | — |
| *Influenza A* |  | 1.5 (1.4, 1.6) | 2.6 (2.3, 2.9) | 0.43 (-0.75, 1.62) | 0.33 (0.28, 0.39) | — | — |
| *Influenza B* |  | 0.8 (0.7, 0.9) | 1.1 (1.0, 1.2) | -0.21 (-1.33, 0.94) | 0.19 (0.15, 0.24) | — | — |
| ***Arboviral / vector-borne*** |  |  |  |  |  |  |  |
| **Dengue** | Log-normal | 6.3 (5.4, 7.8) | 8.8 (7.4, 10.9) | 1.79 (1.61, 1.98) | 0.20 (0.18, 0.22) | — | 0.31 (0.20, 0.52) |
| *Inoculation* |  | 6.7 (4.6, 12.7) | 9.5 (6.5, 18.0) | 1.78 (1.34, 2.20) | 0.21 (0.18, 0.25) | — | 0.49 (0.27, 1.03) |

| Pathogen | Distribution | Median (days) | $p_{95}$ (days) | $\mu_0$ | $\phi$ | $\kappa$ | $\tau$ |
| --- | --- | --- | --- | --- | --- | --- | --- |
| *Mosquito bite* |  | 6.0 (5.6, 6.4) | 8.3 (7.7, 9.1) | 1.80 (1.13, 2.52) | 0.20 (0.17, 0.23) | — | — |
| **Zika** | Log-normal | 6.0 (5.2, 6.8) | 9.9 (8.5, 11.9) | 1.93 (0.86, 3.21) | 0.31 (0.25, 0.39) | — | — |
| **Rift Valley fever** | Log-normal | 3.9 (3.6, 4.3) | 5.4 (4.8, 6.3) | 1.36 (0.24, 2.50) | 0.19 (0.14, 0.27) | — | — |
| **Yellow fever** | Weibull | 3.7 (3.3, 4.1) | 5.4 (4.9, 6.2) | 1.39 (0.47, 2.26) | 3.97 (2.88, 5.23) | — | — |
| **Bat-reservoir zoonoses** |  |  |  |  |  |  |  |
| **Nipah** | Burr XII | 9.2 (7.5, 12.3) | 13.4 (10.9, 18.0) | 2.31 (2.03, 2.61) | 5.72 (4.62, 7.24) | 1.96 (1.07, 4.11) | 0.34 (0.20, 0.63) |
| *Bangladesh* |  | 9.4 (6.7, 17.8) | 13.4 (9.3, 25.6) | 2.32 (1.85, 2.79) | 5.84 (4.66, 7.64) | 2.46 (1.03, 6.57) | 0.48 (0.25, 1.03) |
| **Human-to-human viral** |  |  |  |  |  |  |  |
| **Mpox** | Gamma | 7.6 (6.6, 9.1) | 17.9 (15.5, 21.4) | 2.11 (1.96, 2.26) | 3.00 (2.67, 3.35) | — | 0.26 (0.17, 0.45) |
| **Measles** | Log-normal | 14.0 (12.3, 16.5) | 20.5 (18.0, 24.2) | 2.62 (2.47, 2.76) | 0.23 (0.21, 0.25) | — | 0.20 (0.11, 0.41) |
| **Smallpox** | Burr XII | 12.5 (12.1, 12.8) | 16.0 (15.4, 16.9) | 2.64 (1.96, 3.29) | 8.23 (6.80, 10.08) | 2.20 (1.17, 4.61) | — |
| **Environmental / zoonotic bacterial** |  |  |  |  |  |  |  |
| **Cholera** | Burr XII | 1.7 (1.3, 2.5) | 3.6 (2.7, 5.3) | 0.66 (0.30, 1.05) | 3.12 (2.60, 3.75) | 1.68 (1.10, 2.84) | 0.46 (0.29, 0.80) |
| *O1 El Tor* |  | 1.6 (1.4, 1.7) | 2.9 (2.6, 3.3) | 0.73 (-0.26, 1.70) | 3.33 (2.63, 4.33) | 2.23 (1.20, 4.84) | — |
| *O1 El Tor Ogawa* |  | 1.2 (0.8, 1.8) | 5.5 (3.5, 9.5) | 1.39 (0.08, 2.54) | 1.17 (0.97, 1.42) | 4.04 (2.07, 8.59) | — |
| *O139* |  | 1.2 (1.1, 1.4) | 1.9 (1.6, 2.4) | 0.32 (-0.43, 1.08) | 4.99 (3.39, 8.37) | 1.62 (0.66, 4.70) | — |
| *O1 Classical* |  | 2.3 (1.6, 5.0) | 4.4 (2.9, 9.4) | 0.82 (0.30, 1.35) | 3.92 (3.12, 5.01) | 1.40 (0.79, 2.58) | 0.51 (0.25, 1.14) |
| **Typhoid** | Burr XII | 13.9 (11.4, 17.8) | 29.5 (24.0, 38.2) | 2.40 (2.19, 2.62) | 5.23 (4.68, 5.85) | 0.65 (0.53, 0.81) | 0.46 (0.34, 0.66) |
| *Catered meal* |  | 13.9 (13.0, 14.9) | 30.6 (26.9, 36.3) | 2.61 (1.83, 3.47) | 4.13 (3.21, 5.32) | 0.86 (0.53, 1.54) | — |
| *Experimental* |  | 8.7 (8.2, 9.2) | 23.3 (20.0, 27.9) | 1.93 (1.29, 2.67) | 6.37 (5.03, 8.03) | 0.38 (0.27, 0.53) | — |
| *Other* |  | 15.2 (10.5, 27.5) | 26.5 (18.3, 48.2) | 2.93 (2.45, 3.41) | 3.44 (2.97, 4.03) | 2.87 (1.58, 5.79) | 0.58 (0.36, 1.05) |
| *Water* |  | 17.9 (16.7, 19.2) | 29.2 (27.1, 31.8) | 3.01 (2.28, 3.72) | 4.78 (4.01, 5.76) | 1.58 (0.96, 2.94) | — |

Estimated incubation periods varied substantially across pathogens (Table 1). The shortest median incubation periods were observed for cholera (1.7 days, 95% CrI 1.3–2.5) and influenza subtypes (0.8–4.1 days), while the longest were observed for measles (14.0 days, 95% CrI 12.3–16.5) and typhoid fever (13.9 days, 95% CrI 11.4–17.8). The 95th percentile showed even greater variation, ranging from 1.1 days for influenza B to 29.5 days for typhoid fever.

Data-sparse pathogens are shown in panel C (Figure 3). For Marburg virus disease, the two available datasets yield a median estimate of 8.5 days (95% CrI 8.0–9.0) with a 95th percentile of 14.3 days (95% CrI 12.8–16.5); however, a case report describing a substantially longer incubation period (red arrow) could not be incorporated into the analysis as only two longer than expected incubation periods were reported[24]. Given the extreme sparsity of Marburg data, this suggests the right tail of the distribution may be underestimated, and quarantine periods derived from this estimate should be interpreted with caution. For Lassa fever, estimates are based on a single dataset describing nosocomial transmission, which is not the dominant route of acquisition; results should therefore be interpreted cautiously and may not generalise to community or zoonotic exposure settings.

### Heterogeneity analysis of incubation period distributions

Between-study heterogeneity (τ) was estimated for pathogens with five or more contributing datasets and was generally modest, with most pathogens yielding posterior median estimates in the range 0.2–0.5 (Table 1). The lowest heterogeneity was observed for measles (*τ* = 0.20, 95% CrI 0.11–0.41) and mpox (*τ* = 0.26, 95% CrI 0.17–0.45), consistent with well-characterised, methodologically similar incubation periods. Nipah showed moderate heterogeneity overall (*τ* = 0.34, 95% CrI 0.20–0.63), with the Bangladesh subgroup showing somewhat greater variation (*τ* = 0.48, 95% CrI 0.25–1.03), likely reflecting differences in exposure setting and case ascertainment across outbreaks. Higher heterogeneity was observed for CCHF (*τ* = 0.63, 95% CrI 0.41–1.07), partly attributable to differences in transmission route, as subgroup analyses separating tick-bite from nosocomial acquisition substantially reduced residual heterogeneity. For dengue, the inoculation subgroup showed higher heterogeneity (*τ* = 0.49, 95% CrI 0.27–1.03) than the overall estimate (*τ* = 0.31, 95% CrI 0.20–0.52), while *τ* was not estimated for the mosquito bite subgroup independently, reflecting the more limited number of studies in that subgroup. For cholera, the overall *τ* = 0.46 (95% CrI 0.29–0.80) reflects variation across strains, which showed markedly different median incubation periods ranging from 1.2 days for the O139 strain to 2.3 days for O1 Classical; within-strain *τ* was not estimated for most subgroups due to limited data per strain. Typhoid fever showed substantial heterogeneity across exposure pathways (*τ* = 0.46, 95% CrI 0.34–0.66 overall), consistent with a dose-response effect, whereby experimental challenge studies, which use controlled and typically higher doses, yielded the shortest median incubation period (8.7 days, 95% CrI 8.2–9.2), while waterborne exposures, which tend to deliver lower infectious doses, yielded the longest (17.9 days, 95% CrI 16.7–19.2), with foodborne exposures from catered meals intermediate at 13.9 days (95% CrI 13.0–14.9). For COVID-19, the overall *τ* = 0.42 (95% CrI 0.34–0.54) was accompanied by a clear progressive shortening of the incubation period across variants, from Wildtype (6.4 days, 95% CrI 5.6–7.4) through Alpha (4.6 days, 95% CrI 4.3–4.9) and Delta (4.5 days, 95% CrI 2.7–12.6) to Omicron (3.4 days, 95% CrI 2.9–4.4). Within-variant heterogeneity was low for Wildtype (*τ* = 0.36, 95% CrI 0.27–0.49), reflecting the large number of contributing studies, and for Omicron (*τ* = 0.27, 95% CrI 0.15–0.56), but remained substantial for Delta (*τ* = 0.75, 95% CrI 0.43–1.45), which was based on only seven studies. For SARS, geographic subgroup analyses revealed higher heterogeneity in China (*τ* = 0.63, 95% CrI 0.43–0.99) and Singapore (*τ* = 0.52, 95% CrI 0.24–1.24) relative to Hong Kong (*τ* = 0.31, 95% CrI 0.17–0.62), suggesting real differences in case ascertainment or exposure patterns across settings.

### The value of federated analysis

To quantify the contribution of each data format to inference, we compared three analysis arms: individual-level data only (*I*), summary-statistics only (*S*), and a federated arm (*F*) that combined both formats within the same unified likelihood. The summary-statistics arm is itself a form of federated analysis, since the underlying observations remain decentralised with the original data holders, and only published summaries are accessible. The federated arm extends this by additionally incorporating shared individual-level records.

We first considered the information gain, defined as the ratio of the 95% credible interval width obtained under arm *I* or arm *S* to that under *F*; values greater than one indicate greater precision in the federated arm. For the population-level mean *μ*_0_ (Figure 4A), we found consistent information gain across all pathogens. For most pathogens the gain over the summary-statistics arm was smaller than the gain over the individual-data arm, because more datasets were available as summary statistics than as individual records, and the summary-statistics arm therefore already benefited from a larger evidence base. Typhoid fever was an exception, with 18 individual-level studies but only 4 summary-statistics datasets, so here the gain over the individual-level data arm was instead the smaller. For the 95th percentile (Figure 4B) the picture was less consistent. Summary statistics, and in particular the mean with standard deviation and the median with interquartile range, carried limited information about the distribution’s right tail. As a consequence, the federated arm yielded only marginally narrower, and in a few cases marginally wider, credible intervals for the 95th percentile than the individual-data arm alone.

**Figure 4.**
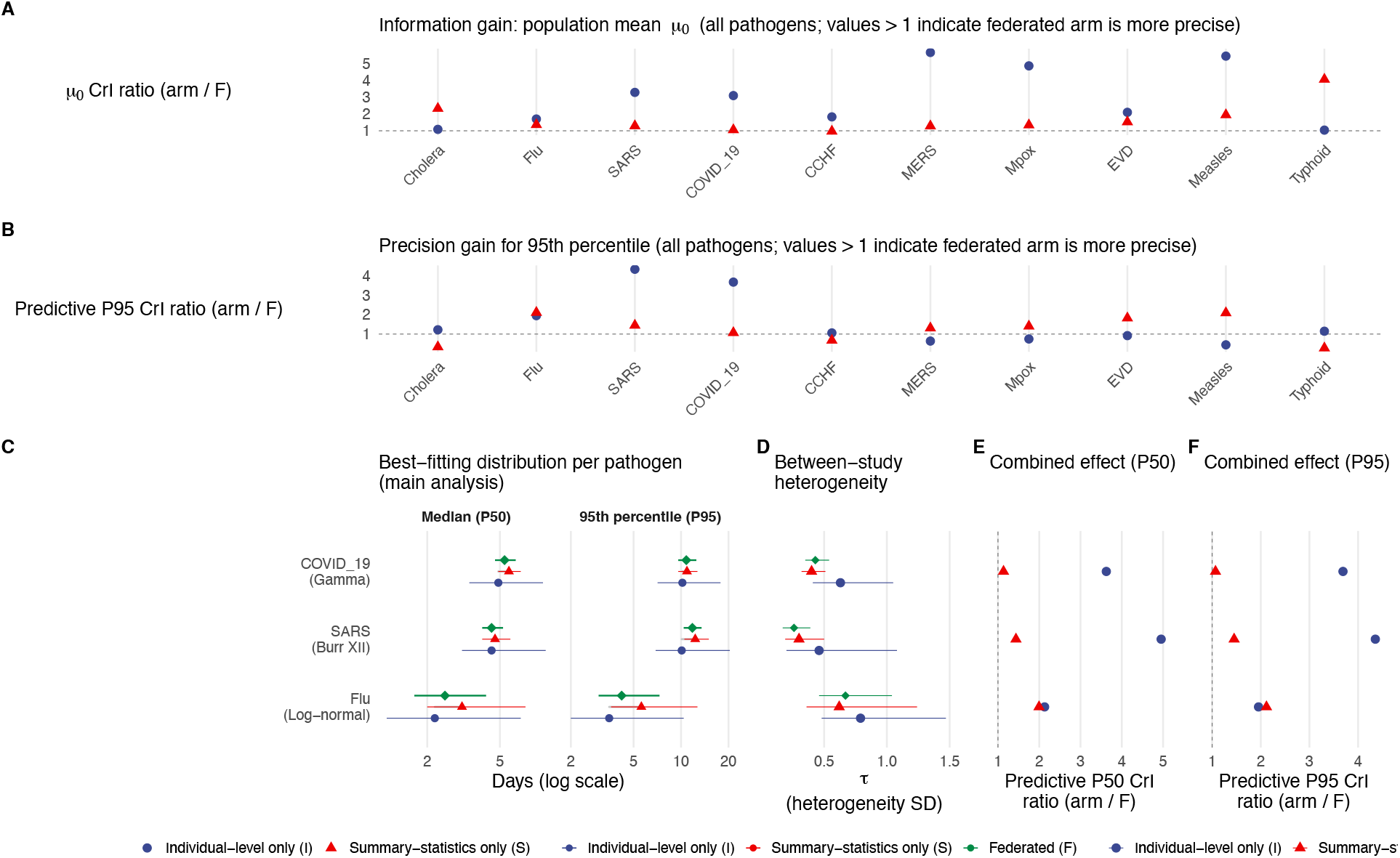
Comparison of data format arms across all pathogens: Panels A and B show all pathogens with available fits; panels C–F are restricted to pathogens where between-study heterogeneity (*τ*) was estimable in all three arms (n ≥5 datasets per arm). Three arms are compared: individual-level data only (*I*), summary statistics only (*S*), and federated (*F* = *I* + *S*). Panels A and B show ratios of 95% credible interval (CrI) widths relative to arm F for the population-level location parameter *μ*_0_ (panel A) and the posterior predictive 95th percentile (panel B); values greater than 1 indicate arm F is more precise. Panel C shows posterior predictive medians and 95th percentiles with 95% CrIs under the best-fitting distribution per pathogen (selected by LOO), with grey segments connecting the I and S estimates. Panel D shows posterior estimates of between-study heterogeneity *τ* per arm. Panels E and F show ratios of 95% CrI widths for the posterior predictive median (P50) and 95th percentile (P95), respectively, combining the gain in *μ*_0_ precision with the contribution of *τ* uncertainty.

Three pathogens, COVID-19 (Omicron), SARS (Hong Kong) and influenza, had sufficient datasets in each arm to estimate the between-study heterogeneity *τ* separately under *I, S* and *F* (Figure 4D). For these we could assess the federated arm through the predictive median and predictive 95th percentile, which reflect both the gain in *μ*_0_ precision and the additional uncertainty from *τ* (Figure 4E, F). Across all three pathogens the federated arm yielded consistently narrower predictive intervals than either single-format arm, demonstrating that the overall effect of incorporating more studies, even when those studies contributed only summary statistics, was a consistent improvement in policy-relevant predictive precision.

### Comparison to classical meta-analysis

To benchmark the Bayesian hierarchical model against standard practice, we fitted a classical random-effects meta-analysis using the restricted maximum-likelihood estimator to the 12 pathogens with five or more usable datasets (Supplementary Table S4, Methods 4). Across all 12 pathogens, *I*^2^ was high, ranging from 61.6% (Nipah) to 99.3% (COVID-19), indicating that between-study heterogeneity dominates total variance and that a single pooled mean is not a reliable summary for any of the pathogens considered. The Bayesian model consistently estimated higher between-study heterogeneity than classical meta-analysis across all pathogens. The largest absolute discrepancy was observed for Nipah, where the meta-analysis yielded *τ* = 0.016 compared to *τ* = 0.33 (95% CrI 0.27–0.86) from the best-fitting Bayesian model; here, the meta-analysis is restricted to reported or inferred conventional summary statistics, while the Bayesian model additionally incorporates frequency table data directly through the full likelihood, making more efficient use of the available evidence. More broadly, this systematic underestimation of *τ* by classical meta-analysis likely reflects the information loss inherent in approximating study-level distributions by their means and standard deviations, which discards information about distributional shape that helps distinguish within-study variability from between-study heterogeneity.

## Discussion

We have presented a Bayesian hierarchical framework for inference of the incubation period distribution that can jointly synthesise individual-level observations and heterogeneous summary statistics under a unified likelihood. The approach substantially advances on traditional methods for estimating incubation period distributions that rely on individual-level data, are predominantly pooled and rarely include hierarchical structure. Our method also improves on classical meta-analysis, which discards within-study distributional information because it is based solely on summary statistics, characterises only the central estimate of the distribution rather than the full distribution, and, as demonstrated here, systematically underestimates between-study heterogeneity (Supplementary Table S1). By extending the distributional family to include the Burr Type XII and generalised gamma, the framework can capture the heavy-tailed behaviour observed in the majority of pathogens studied more flexibly. This has direct consequences for quarantine policy as a heavier right tail implies a longer period of monitoring required to contain a given proportion of cases, and estimates based on lighter-tailed distributions will systematically underestimate this window.

The practical necessity of this Bayesian hierarchical approach is illustrated by the composition of the assembled dataset (Supplementary Table S5). For 9 of 18 pathogens, including Ebola, CCHF, SARS, MERS, Nipah, mpox, influenza, RVF, and Zika, summary statistics provide the majority of contributing datasets, and individual-level data alone would leave each pathogen below the ten-dataset threshold identified by the simulation study as required for reliable inference. For Nipah in particular, only a single frequency-table dataset is available as individual-level data; the remaining ten datasets exist only as summary statistics. By contrast, pathogens with large bodies of historic individual-level data, such as cholera and typhoid fever, benefit less from summary statistic incorporation in terms of dataset counts, though summary statistics still contribute meaningfully to subgroup analyses where individual dataset counts are sparse.

Our simulation study identified practical operating conditions for the framework. Performance is robust with ten or more contributing studies of moderate size, but degraded in sparse settings, particularly for the 95th percentile and for the generalised gamma, which presents identifiability challenges in contexts with few datasets. Fewer studies may suffice, however, if at least one contains substantial individual-level data. These thresholds inform which of the 18 pathogen estimates should be treated as well-supported and which warrant greater caution.

The relationship between the incubation period distribution and outbreak controllability has been characterised formally by Fraser et al. [3], who showed that when the left tail of the incubation period is short relative to the generation time, a substantial proportion of transmission occurs before symptom onset, limiting the effectiveness of symptom-based contact tracing; our estimates reflect this directly. Influenza subtypes show the shortest incubation periods (0.8–4.1 days across subtypes), consistent with well-documented pre-symptomatic infectiousness and the limited effectiveness of contact tracing as a sole control measure for seasonal influenza [25, 26]. By contrast, a pathogen such as measles (14.0 days, 95% CrI 12.3–16.5) has a long incubation period that, in principle, provides a window for intervention, although this window must be weighed against the practical challenges of identifying and tracing contacts over longer periods.

Our estimates for well-studied pathogens are broadly consistent with existing systematic reviews and large-scale studies such as ComCor [27], providing external validation of the framework. For COVID-19, the progressive shortening of the incubation period across variants, from Wildtype (6.4 days) to Omicron (3.4 days), mirrors trends reported elsewhere and reflects genuine biological differences between variants as well as differences in the underlying evidence base[27]. The substantial residual heterogeneity within Delta (τ = 0.75), based on only seven contributing studies, highlights how rapidly evolving pathogen-context combinations can produce sparse and variable evidence even for intensively studied pathogens.

For cholera, the high overall heterogeneity (*τ* = 0.46) and markedly different incubation periods across strains reflect genuine biological and epidemiological differences, but the limited number of studies per strain means that strain-specific estimates carry a substantial risk of misspecification. Additionally, the dataset is sensitive to the inclusion of outlier observations, such as rectal swab samples from asymptomatic patients, which may not reflect the typical symptomatic onset of disease [28]. These issues underscore a broader limitation of the evidence base, as for most pathogens, the available data are sparse, outdated, and derived from study designs, including human challenge experiments, that may not reflect natural exposure in contemporary populations.

For data-sparse pathogens, narrow credible intervals should not be interpreted as precision. Where few studies are available, the model fits the available data well but cannot capture variation that is simply absent from the evidence base. The Marburg case illustrates this, as the reported estimate is precise relative to the data, but a case report describing a substantially longer incubation period suggests that the right tail may be underestimated, and that derived quarantine recommendations should be treated with corresponding caution. Many key epidemiological parameters are similar between Marburg virus disease and Ebola virus disease [29], and this is reflected in the comparable central incubation period estimates across the two pathogens (8.5 days for MVD versus 8.9 days for EVD overall). For EVD, the West Africa outbreak provides a large and well-characterised evidence base [30], yielding a 95th percentile of 21.1 days (95% CrI 16.8–30.0) for that subgroup, which is consistent with the longer incubation periods described in the MVD case report. Together, this suggests that the central estimate for MVD is broadly plausible but that the right tail of the distribution, and consequently the recommended quarantine window, is likely underestimated due to the sparsity of available data. While the simulation study identified a threshold of ten or more contributing datasets as a practical safeguard against right-truncation bias in upper-tail estimates, as illustrated by the Marburg case report describing a substantially longer incubation period than the sparse available data could capture, exposure uncertainty arising from recalled or estimated rather than precisely documented exposure events remains an unmeasured source of bias that cannot be corrected without additional information on the underlying exposure distribution.

To assess the sensitivity of our results to the factorised likelihood used throughout the primary analysis, we fitted an alternative model in which the joint density of the three reported order statistics was used for all datasets contributing a median with range (summary type 1) or median with interquartile range (summary type 2) (full details are given in the Supplementary Methods, the joint and factorised formulations are compared in Supplementary Table S3). Posterior median estimates of the predicted median incubation period were essentially unaffected across all pathogens and distributions, with differences well below the precision of the reported source data. Estimates of the 95th percentile differed by at most a quarter of a day. Credible intervals were marginally wider under the joint model, consistent with the expectation that the factorised likelihood inflates the effective information content of order-statistic summaries by treating correlated statistics as independent. The practical impact on inference is therefore negligible for the sample sizes and summary statistics present in this review, and we retain the factorised formulation as the primary model on the grounds of computational robustness and efficiency.

The framework has several remaining limitations. It does not account for dose-response relationships, nor for pathogens such as anthrax, where the incubation period is known to be strongly dose-dependent [31]. However, distributional synthesis of this kind is of limited utility without explicit dose information. The assumption that all studies within a pathogen share a common distributional family is a modelling simplification that may not hold when studies span radically different exposure contexts or populations. Similarly, the model assumes a shared within-study dispersion parameter across studies, which may introduce bias when study designs differ substantially. Many older studies rely on recalled or estimated exposure dates rather than precisely documented events, introducing measurement error in the reported incubation period that cannot be accounted for within the model. Future extensions of the framework could incorporate study-level quality weights to address the potential for publication bias, given that studies reporting incubation periods are not a random sample of outbreak investigations and may systematically over-represent well-documented clusters with identifiable exposure events.

Beyond incubation periods, the same structural data heterogeneity also arises for serial intervals, generation times, onset-to-hospitalisation delays, and hospitalisation-to-death delays. The ddsynth R package provides both the curated incubation period dataset and a general methodological framework applicable to any delay distribution of interest and compatible with the federated epidemic inference architectures now being proposed for multi-site outbreak response [13]; for other delays, such as serial intervals or onset-to-hospitalisation times, compatible datasets are available from the epireview[32] and grEPI[33] packages.

## Methods

### Bayesian hierarchical model for incubation period federated analysis

Incubation period data reported in the literature are heterogeneous in format as different studies report different summary statistics (median and range, median and interquartile range, mean and standard deviation, or individual-level frequency counts), use different sample sizes, and may exhibit genuine between-study variation arising from differences in case ascertainment, study population, or exposure definition.

Classical random-effects meta-analysis can synthesise estimates of a single central quantity (such as the median) across studies, but does not characterise the full incubation-period distribution. Bayesian synthetic likelihood (BSL) approaches [34, 35] offer greater flexibility by replacing the intractable likelihood with a simulation-based approximation, but at substantially higher computational cost and with reduced flexibility in accommodating heterogeneous summary statistic types across studies. To make full use of all available evidence, we developed a Bayesian hierarchical model that directly incorporates each study’s reported individual-level data or summary statistic through an analytic parametric likelihood. The model simultaneously accommodates five summary types: (1) median and range; (2) median and interquartile range; (3) mean and standard deviation; (4) exact-value frequency tables; and (5) interval-censored frequency tables. Studies reporting different summary types can therefore be synthesised within a single joint model without any pre-processing or conversion of the reported statistics.

The model is hierarchical, with up to four parameters of primary interest (Figure 5; Table 2). The population-level log-scale mean *μ*_0_ characterises the typical incubation period across studies. The between-study standard deviation *τ* quantifies heterogeneity in the location of the incubation period distribution across studies, capturing genuine epidemiological variation between outbreak settings. The shape parameter *ϕ* governs the dispersion of individual observations within each study. For the Burr XII [14, 15] and generalised gamma families, a second shape parameter *κ* provides additional flexibility in tail behaviour. Study-specific location parameters loc_*d*_ arise as random deviations from *μ*_0_, scaled by *τ*.

**Figure 5.**
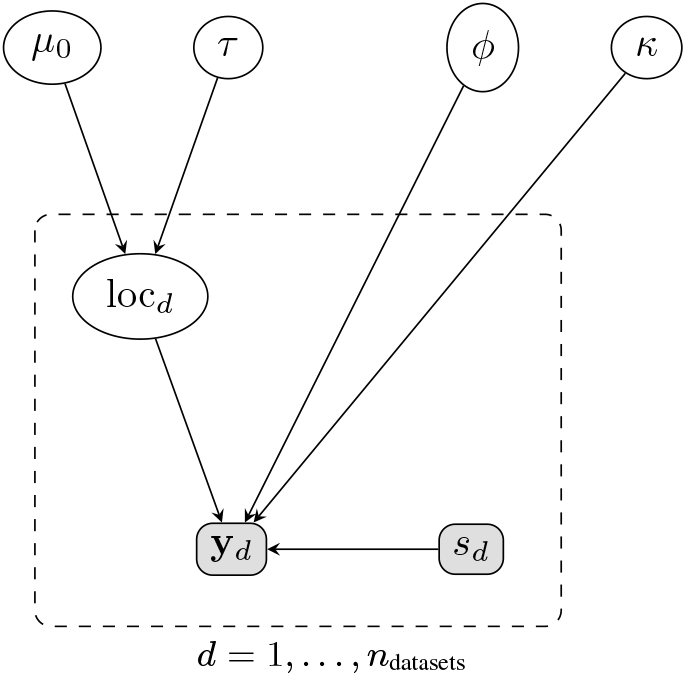
Hierarchical model for Bayesian incubation period synthesis. Population-level hyperparameters (*μ*_0_,*τ, ϕ*, and, for three-parameter families, *κ*) govern study-specific location parameters loc_*d*_, which in turn determine the likelihood of each study’s observed summary statistic **y**_*d*_. The observed summary type *s*_*d*_ selects the appropriate likelihood for each study, allowing datasets reporting different summary statistics to be synthesised jointly within a single model. Shaded nodes are observed; unshaded nodes are latent.

**Table 2.** Parameters of the Bayesian hierarchical incubation period model.

| Parameter | Interpretation | Role | Type |
| --- | --- | --- | --- |
| $\mu_0$ | Population log-scale mean | Typical incubation period across studies | Hyperparameter |
| $\tau = e^{\log \tau}$ | Between-study SD | Heterogeneity across outbreak settings | Hyperparameter |
| $\phi = e^{\log \phi}$ | Shape/dispersion | Spread of individual observations within studies | Hyperparameter |
| $\kappa = e^{\log \kappa}$ | Second shape (Burr XII, gen. gamma only) | Tail behaviour; unused by log-normal, gamma, Weibull | Hyperparameter |
| $\text{loc}_d$ | Study-level location | Study-specific mean on the log scale | Latent variable |

We fitted five parametric distributional families (log-normal, gamma, Weibull, Burr Type XII, and generalised gamma), each of which is a right-skewed, non-negative distribution appropriate for time-to-event data. The log-normal, gamma, and Weibull families have each been previously used to characterise incubation period data [7]. The log-normal is characterised by its log-scale mean and standard deviation; the gamma and Weibull by a log-scale location and a shape parameter. The Burr Type XII and generalised gamma are three-parameter families that extend this framework with an additional shape parameter *κ*: the Burr Type XII is parameterised by a scale 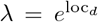 and two shape parameters (*c* = *ϕ, k* = *κ*), while the generalised gamma uses the Prentice parameterisation with location *μ* = loc_*d*_, log-dispersion *σ* = *ϕ*, and shape *Q* = *κ* [36, 37]. The three two-parameter families can be recovered as special or limiting cases of the generalised gamma (*Q* = 1 yields the Weibull; *Q* = *σ* yields the log-normal as *Q* → 0; 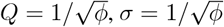 yields the gamma). Model selection between the five families was performed using Leave-one-out cross validation [38]. Full details of the likelihood construction, prior specification, and posterior predictive summaries are given in the following section.

**Population level:** *μ*_0_, *τ, ϕ, κ* (priors defined in text)

**Study level:** loc_*d*_ ~ 𝒩 (*μ*_0_, *τ*^2^)

**Observed summary: y**_*d*_ | loc_*d*_, *ϕ, κ, s*_*d*_ ~ *p*(**y**_*d*_ | loc_*d*_, *ϕ, κ, s*_*d*_)

where *s*_*d*_ ∈ *{*1, 2, 3, 4, 5*}* is the *observed* summary type reported by study *d* (shaded node), and the underlying individual-level distribution *f* (·; loc_*d*_, *ϕ, κ*) is one of:

- Log − normal (loc_*d*_, *ϕ*),
- Gamma (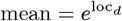, shape = *ϕ*),
- Weibull (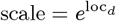, shape = *ϕ*),
- Burr XII (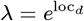, *c* = *ϕ, k* = *κ*),
- Gen. Gamma (*μ* = loc_*d*_, *σ* = *ϕ, Q* = *κ*).

*Note*: Shaded nodes denote observed quantities; unshaded nodes are latent. For the log-normal, gamma, and Weibull families, *κ* does not enter the likelihood. The centred form loc_*d*_ ~ 𝒩 (*μ*_0_,*τ* ^2^) is shown for clarity; a non-centred reparameterisation loc_*d*_ = *μ*_0_ + *τ* ·*z*_*d*_, *z*_*d*_ ~ 𝒩 (0, 1), is used in the Stan implementation to improve HMC sampling efficiency [39].

### Bayesian hierarchical model with parametric joint individual-level data and summary-statistic likelihood

In line with the model described in the previous section, we have hyperparameters *μ*_0_ ∈ ℝ, *τ >* 0, *ϕ >* 0, and, for the Burr XII and generalised gamma families, a second shape parameter *κ >* 0. We model study-level random effects as *z*_*d*_ for *d* = 1, ···, *n*_datasets_ so that the study specific location becomes loc_*d*_ = *μ*_0_ + *τ* ·*z*_*d*_. This non-centred parameterisation improves HMC sampling efficiency by decorrelating the global hyperparameters from the study-level deviations [40]. The underlying distribution of dataset *d* is 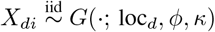, where *G* is one of five distributional families, with

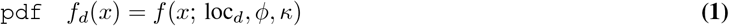

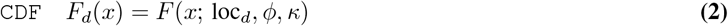

The five distributions are parameterised as follows, with loc_*d*_ controlling the scale of the distribution on the log scale, *ϕ* controlling its shape, and, where applicable, *κ* providing additional flexibility in tail behaviour:

- **Log-normal**: Log-normal(loc_*d*_, *ϕ*), where loc_*d*_ is the log-scale mean and *ϕ* = *σ* is the log-scale standard deviation.
- **Gamma**: 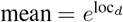, shape = *ϕ*, 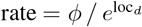.
- **Weibull**: 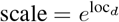, shape = *ϕ*.
- **Burr Type XII**: 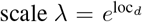, first shape *c* = *ϕ*, second shape *k* = *κ*, with CDF *F* (*x*) = 1 1+ (*x/λ*)*c* −*k*. Finite moments require *κϕ > r* for the *r*-th moment; in particular, a finite mean requires *κϕ >* 1 and a finite variance requires *κϕ >* 2.
- **Generalised gamma** (Prentice parameterisation [36, 37]): location *μ* = loc_*d*_, log-dispersion *σ* = *ϕ*, shape *Q* = *κ*, with derived shape *γ* = 1*/Q*^2^ and standardised log-variate *w* = (log *x* − *μ*)*/σ*. The CDF is *F* (*x*) = Γ_reg_ (*γ e*^*Qw*^; *γ*), where Γ_reg_ denotes the regularised lower incomplete gamma function. The log-normal, Weibull, and gamma arise as special or limiting cases.

For the log-normal, gamma, and Weibull families, *κ* does not enter the likelihood and is assigned a weakly informative prior that does not influence inference. We have observed summary statistics **y**_*d*_ for each dataset, and the summary type *s*_*d*_ determines which likelihood is used:

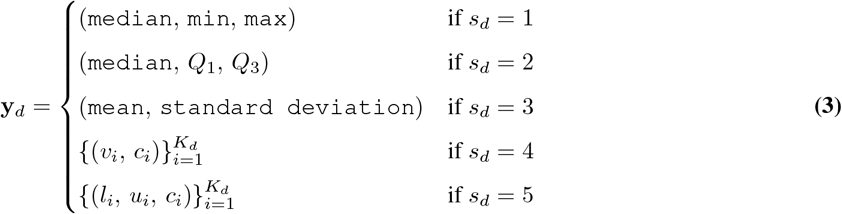

For *s*_*d*_ = 4, the data are a frequency table with *K*_*d*_ entries. Each entry (*v*_*i*_, *c*_*i*_) records a distinct observed value *v*_*i*_ (e.g. incubation period in days) and its count *c*_*i*_ ≥1; the total sample size is 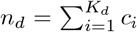. This representation is equivalent to the full list of *n*_*d*_ individual observations, but exploits the grouping for computational efficiency. For *s*_*d*_ = 5, the data are provided as an interval-censored frequency table of *K*_*d*_ entries. Each entry (*l*_*i*_, *u*_*i*_, *c*_*i*_) records the lower bound *l*_*i*_, upper bound *u*_*i*_ ≥*l*_*i*_, and count *c*_*i*_ ≥1 of individuals whose true value is known only to lie in the interval [*l*_*i*_, *u*_*i*_]. When *l*_*i*_ = *u*_*i*_, the observation is exact; when *l*_*i*_ *< u*_*i*_, it is interval-censored. This generalises *s*_*d*_ = 4, which corresponds to the special case *l*_*i*_ = *u*_*i*_ = *v*_*i*_ for all *i*.

#### Priors

We use the following priors:

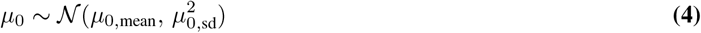

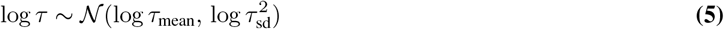

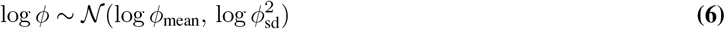

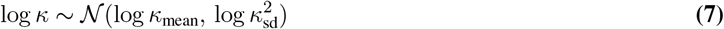

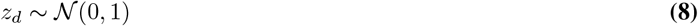

with joint prior

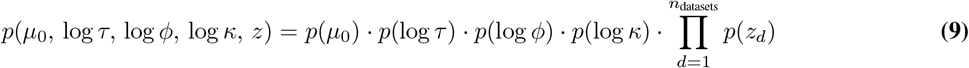

All prior hyperparameters are passed as data to the Stan model. Because *ϕ* and *κ* have different interpretations across distributions, the default values of log *ϕ*_mean_ and log *κ*_mean_ are set separately for each family to place the prior mass over a plausible range (Table 3). All other hyperparameters share the same defaults regardless of the chosen distribution.

**Table 3.** Distribution-specific default priors for log *ϕ* and log *κ*.

| Distribution | $\phi / \kappa$ meaning | log $\phi$ prior | | log $\kappa$ prior | | Prior median ($\phi, \kappa$) |
| --- | --- | --- | --- | --- | --- | --- |
|  |  | mean | sd | mean | sd |  |
| Log-normal | $\phi = \sigma$ (log-SD); $\kappa$ unused | -0.7 | 0.5 | — | — | (0.50, —) |
| Gamma | $\phi = \text{shape}$; $\kappa$ unused | 2.5 | 0.5 | — | — | (12.2, —) |
| Weibull | $\phi = \text{shape}$; $\kappa$ unused | 1.0 | 0.5 | — | — | (2.7, —) |
| Burr XII | $\phi = c$ (shape 1); $\kappa = k$ (shape 2) | 0.7 | 0.5 | 1.0 | 0.5 | (2.0, 2.7) |
| Gen. Gamma | $\phi = \sigma$ (log-disp.); $\kappa = Q$ (shape) | -0.5 | 0.5 | 0.0 | 0.5 | (0.6, 1.0) |

We note an identifiability issue of *τ* with few studies. When *n*_datasets_ *<* 5, the between-study heterogeneity parameter *τ* cannot be reliably identified from the data, and its posterior is largely determined by the prior [41–43]. In this setting, predicted quantities are computed at the mean of the estimated study-level location parameters, 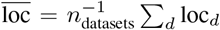, rather than sampling new locations from 𝒩 (*μ*_0_,*τ* ^2^, to avoid prior-dominated inflation of predictive intervals. In the results we do not report *τ* estimates for the case of *n*_datasets_ *<* 5.

#### Order-statistics likelihood for median and range

Let the order statistics of the unobserved sample be 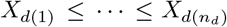. For a general order statistic *X*_*d*(*k*)_, the pdf is

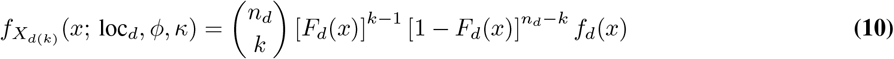

We define the index function for each dataset as follows:

- median index: *k*_med,*d*_ = *l*(*n*_*d*_ + 1)*/*2*J*
- min index: *k*_min_ = 1
- max index: *k*_max_ = *n*_*d*_

and hence median = 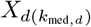, min = *X*_*d*(1)_, 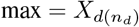. The likelihood is

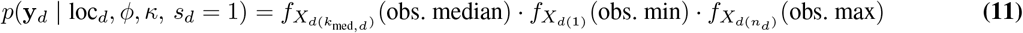

The log-likelihood for the order statistic involves terms of the form (*k*− 1) log *F*_*d*_(*x*) and (*n*_*d*_− *k*) log[1 −*F*_*d*_(*x*)]. A naive implementation that first evaluates *F*_*d*_(*x*) and then takes logarithms is numerically unstable when *F*_*d*_(*x*) is close to 0 or 1, and clipping *F*_*d*_(*x*) to a small interval away from these boundaries produces zero gradients at the clip boundaries, which can impair HMC sampling. We therefore evaluate log *F*_*d*_(*x*) and log[1 − *F*_*d*_(*x*)] directly using the numerically stable log-CDF and log-complementary-CDF functions provided by Stan (`_lcdf` and `_lccdf`), which preserve correct gradient information throughout the parameter space. Terms with a zero coefficient (i.e. *k* = 1 or *k* = *n*_*d*_) are omitted to avoid 0 *×* (−∞) = `NaN` in the gradient computation.

#### Order-statistics likelihood for median and interquartile range

The construction of the likelihood for the median and IQR follows the same pattern as in the previous section, except that we define

- quartile 1 index: 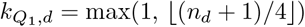
- quartile 3 index: 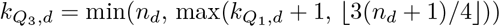

and the likelihood is

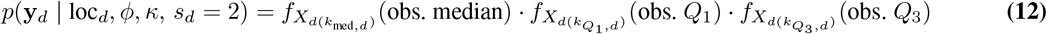

#### Likelihood for mean and standard deviation

Let *μ*_*d*_ = 𝔼 [*X*_*d*_] and 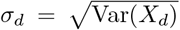 for the chosen distribution and parameters loc_*d*_, *ϕ, κ*. The analytical forms are:

- **Log-normal**: 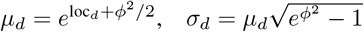
- **Gamma**: 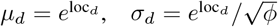
- **Weibull**: 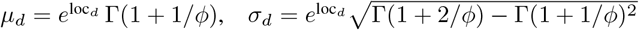
- **Burr XII** (with 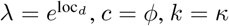; requires *kc >* 2):

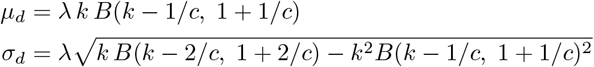

where *B*(·, ·) denotes the beta function. When *kc* ≤ 2 the variance is infinite and the target density is set to −∞ for this summary type.
- **Generalised gamma** (with *γ* = 1*/κ*^2^):

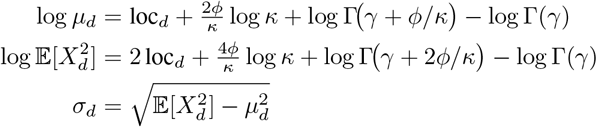

All computations are performed on the log scale for numerical stability.

By the central limit theorem and the delta method, the observed sample mean 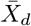 and sample standard deviation *S*_*d*_ have approximate sampling distributions

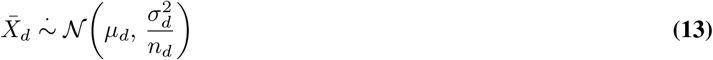

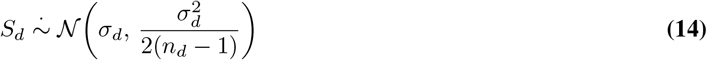

and the likelihood becomes

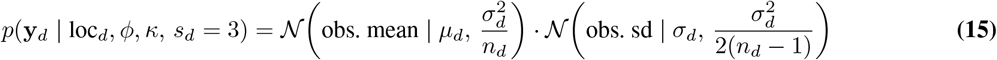

The Gaussian sampling distributions above are asymptotic approximations that become increasingly accurate as *n*_*d*_ grows. For small samples (e.g. *n*_*d*_ *<* 20) from heavily skewed distributions, the true sampling distribution of 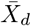 may exhibit noticeable skewness and that of *S*_*d*_ may be poorly approximated by a symmetric distribution. In practice, studies reporting mean and standard deviation typically have moderate to large sample sizes, as small studies more commonly report the median and range or IQR [44]. Where small-sample mean–SD studies are included, the resulting likelihood contribution should be interpreted with appropriate caution.

#### Likelihood for frequency tables (*s_d_* = 4)

When *s*_*d*_ = 4, the dataset is provided as a frequency table with *K*_*d*_ entries 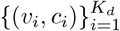, where *v*_*i*_ is the *i*-th distinct observed value and *c*_*i*_ is its count, with 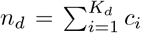. This is equivalent to direct likelihood evaluation on all *n*_*d*_ individual observations, exploiting the grouping for computational efficiency:

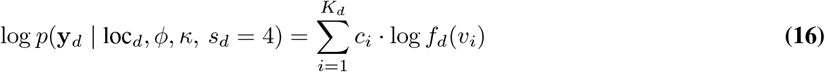

#### Likelihood for interval-censored frequency tables (*s_d_* = 5)

When *s*_*d*_ = 5, each individual is known only to have a value in the interval [*l*_*i*_, *u*_*i*_]. The probability that a single observation falls in this interval is *F*_*d*_(*u*_*i*_) −*F*_*d*_(*l*_*i*_), so the log-likelihood contribution of the *c*_*i*_ individuals in interval *i* is *c*_*i*_ · log[*F*_*d*_(*u*_*i*_) − *F*_*d*_(*l*_*i*_)]. When *l*_*i*_ = *u*_*i*_, the interval degenerates to a point, and we recover the exact-observation case. The full log-likelihood is therefore

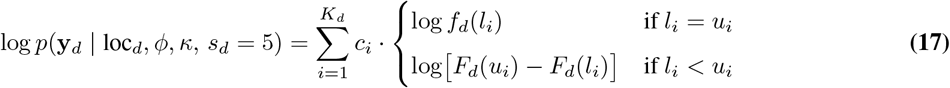

Direct evaluation of log[*F*_*d*_(*u*_*i*_) −*F*_*d*_(*l*_*i*_)] suffers from problematic cancellation when *F*_*d*_(*u*_*i*_) ≈ *F*_*d*_(*l*_*i*_), i.e. when the interval is narrow relative to the distribution’s scale. We therefore compute this quantity as

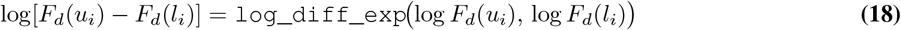

where `log_diff_exp`(*a, b*) = log(*e*^*a*^ −*e*^*b*^) is evaluated in a numerically stable manner, with log *F*_*d*_(·) and log[1 −*F*_*d*_(·)] obtained via Stan’s `_lcdf` and `_lccdf` functions as described in the previous section.

#### Full likelihood

The likelihood for each dataset selects the appropriate term based on the summary type *s*_*d*_:

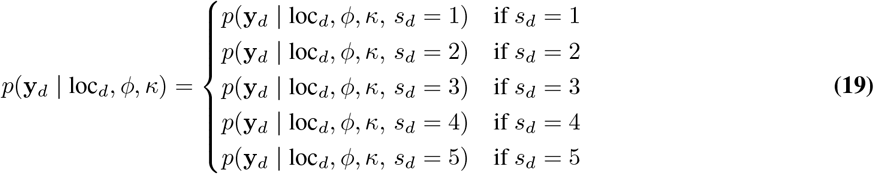

and the full likelihood across all datasets is

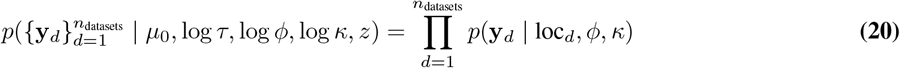

Note that datasets of different summary types can be combined freely within a single model fit, as each contributes its own likelihood term to the joint posterior.

### Alternative data synthesis methods

#### Classical random-effects meta-analysis

To benchmark the Bayesian hierarchical framework against standard practice, we fitted a classical random-effects meta-analysis to all pathogens with five or more usable datasets (12 pathogens in total), pooling log-transformed mean incubation periods using the `metamean` function from the `meta` R package[45]. Between-study variance was estimated using the restricted maximum-likelihood estimator, and all pooled estimates and heterogeneity statistics were computed under a random-effects model. The *I*š statistic and the between-study standard deviation *τ* (on the log scale) were reported for each pathogen alongside the pooled mean and 95% confidence interval back-transformed to days.

Because studies report incubation period data in heterogeneous formats, study-level means and standard deviations were derived as follows. For studies reporting a mean and standard deviation directly, these values were used without transformation. For studies reporting a median with interquartile range or median with range, the mean and standard deviation were approximated using the Luo and Shi methods respectively [46, 47]. For exact-value frequency tables, the mean and standard deviation were computed directly from the individual counts. For interval-censored frequency tables, interval midpoints were used to approximate individual observations, with the standard deviation estimated by combining the between-midpoint variance and the expected within-interval variance under a uniform distribution. Datasets in which all observations were reported as upper bounds only, providing no lower-bound information, were excluded from the meta-analysis. Subgroup analyses were conducted for pathogens with two or more distinct subgroups, using the same random-effects estimator applied to the subgroup-stratified data.

#### Inference using Bayesian synthetic likelihood

As an alternative to the parametric likelihood approach described above, incubation period distributions can be inferred using Bayesian synthetic likelihood (BSL) [34, 35]. BSL is a likelihood-free inference method for models with intractable likelihoods but tractable simulation. It approximates the distribution of informative summary statistics by a multivariate normal distribution estimated from simulated data, yielding a synthetic likelihood that replaces the true likelihood within an MCMC framework. Unlike approximate Bayesian computation, which relies on distance-based acceptance or kernel weighting, BSL directly models the joint distribution of summary statistics, offering improved efficiency and stability in moderate-dimensional settings.

In practice, BSL is substantially more computationally expensive than the analytic likelihood approach and requires the same summary statistic type to be available across all contributing datasets, limiting its applicability to the heterogeneous data structures encountered here. For these reasons, BSL is not used in the primary analysis. However, for users who prefer this approach, ddsynth provides BSL-based inference for the log-normal, gamma, and Weibull families via the BSL R package [48], supporting inference of the population-level log-mean (*μ*_0_), between-study heterogeneity (*τ*), and within-study dispersion (*ϕ*).

### Evaluation of simulation study

Model performance was assessed using four metrics computed across 100 replicates per scenario. Coverage was defined as the empirical proportion of replicates in which the true parameter value or predictive quantile fell within the 95% posterior credible interval, with a nominal target of 0.95. Bias was defined as the median signed error (posterior median minus true value) across replicates, with a target of zero. Predictive accuracy was assessed using the Weighted Interval Score (WIS) [49, 50], a proper scoring rule expressed in days that jointly penalises miscalibration and lack of sharpness; lower values indicate better predictive performance. MCMC convergence was assessed per replicate using the potential scale reduction factor 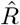 and the effective sample size [51]; replicates with 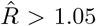 or minimum effective sample size below 100 for any parameter were excluded from performance summaries. Coverage and bias were computed for the population-level log-mean (*μ*_0_), between-study heterogeneity (*τ*), within-study dispersion (*ϕ*), and, for the Burr XII and generalised gamma families, the additional shape parameter (*κ*), as well as for the posterior predictive median (P50) and 95th percentile (P95) as the primary policy-relevant quantities. Full details are provided in the further methods section in the Supplementary information A.2, including full performance summaries, in Supplementary Tables S1 and S2 and Supplementary Figures 2 and S4.

### Data collection from the literature

Data for the 18 pathogens were collected from the literature using three approaches. For WHO priority pathogens, we drew on systematic reviews conducted by the Pathogen Epidemiology Review Group (PERG) at Imperial College London. Published or pre-print reviews were available for Marburg virus disease, Ebola virus disease, Lassa fever, SARS, Zika, and Nipah [16–21]; unpublished PERG reviews were used for MERS, Rift Valley fever, and CCHF.

For the remaining pathogens we used published systematic reviews: influenza [1], COVID-19 [8], dengue [22, 52], yellow fever [22], measles (Public Health Authority of Canada, unpublished, data available via WHO grEPI [33]), cholera [2], and mpox [9]. No systematic reviews of the incubation period were identified for smallpox or typhoid fever; for these two pathogens we conducted a PubMed search from database inception to 1 April 2026 using the terms “SMALLPOX AND INCUBATION” and “TYPHOID FEVER AND INCUBATION” respectively, with title and abstract screening, full-text review, and data extraction performed by a single reviewer.

From each review, we obtained references to the underlying primary studies and extracted either the reported summary statistics or, where available, individual-level data. Where both were available, individual-level data were preferred to maximise inferential precision. For studies reporting data only in figures, values were extracted by digital image processing using Claude Sonnet 4.6 and manual verification. In all cases, care was taken to ensure that individual-level observations and summary statistics were not extracted from the same study simultaneously, to avoid duplication.

### Declaration of generative AI and AI-assisted technologies in the manuscript preparation process

During the preparation of this work the author(s) used Claude Sonnet 4.6 to correct grammar, digital data extraction from figures, and refine & review code. After using this tool/service, the author(s) reviewed and edited the content as needed and take(s) full responsibility for the content of the published article.

## Declarations

### Data & Code availability

All data are publicly available and reference in the supplementary information. The curated data and code are available in the R package `ddsynth` https://github.com/cm401/ddsynth.

### Author contributions

C.M. conceptualised this systematic review. C.M., M.P.K, M.U.G.K. and S.B. formally analysed, visualised, and validated the data. C.M. was responsible for the software infrastructure. S.B. acquired funding. C.M., S.B., and N.M.F were responsible for project administration. S.B. and N.M.F supervised the project. C.M., M.P.K, and S.B. wrote the original manuscript draft. All authors were responsible for the methodology, review, and editing of the manuscript. All authors debated, discussed, edited, and approved the final version of the manuscript. All authors had final responsibility for the decision to submit the manuscript for publication.

