## Supplementary Information for "Federated analysis of incubation period distributions using individual-level observed data and heterogeneous summary statistics"

This Supplementary Information provides additional methodological detail, extended simulation results, and pathogen-specific findings to support the main text. Figure S1 provides a schematic overview of the model. Appendix A describes the posterior predictive CDF computation, the simulation study evaluation framework, theoretical limitations of the generalised gamma distribution for summary-statistic data, and the joint order-statistic likelihood. Appendix B presents simulation study sensitivity analyses, a comparison of the factorised and joint likelihood approaches, and a comparison to classical meta-analysis. Appendix C provides pathogen-specific results for each of the 18 pathogens included in the analysis.

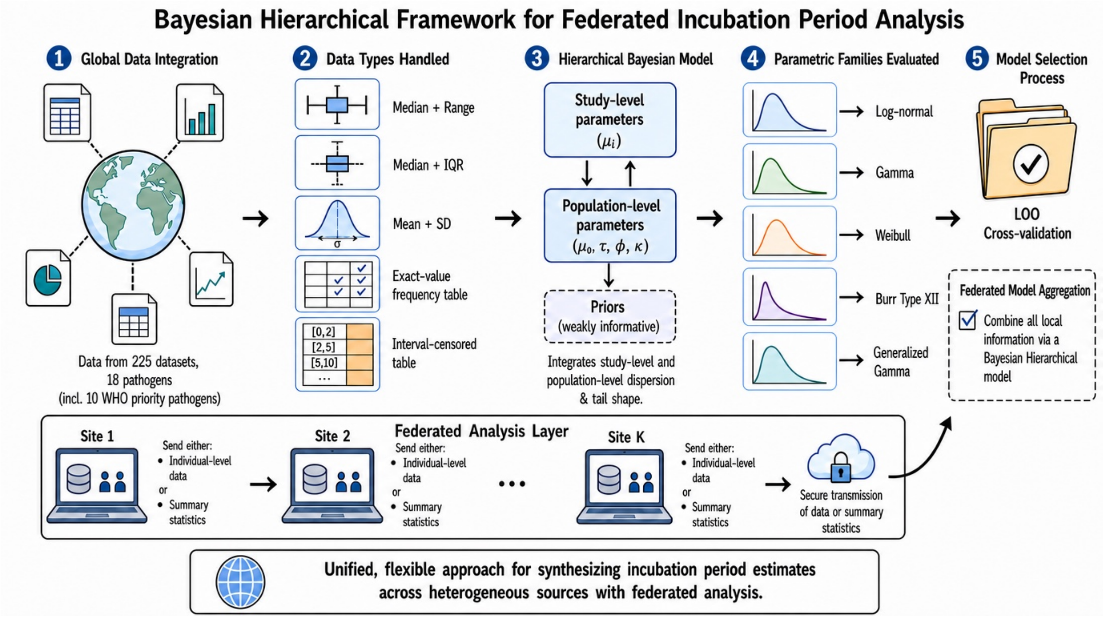

**Supplementary Figure S1.** Schematic overview of the study

The 18 pathogens we consider are organised into six epidemiological groups based on shared transmission route and clinical presentation (Figure S2). Pathogens classified as WHO Research and Development Blueprint priorities are highlighted. Crimean-Congo haemorrhagic fever is retained within the viral haemorrhagic fevers group on clinical grounds, notwithstanding its tick-borne transmission route.

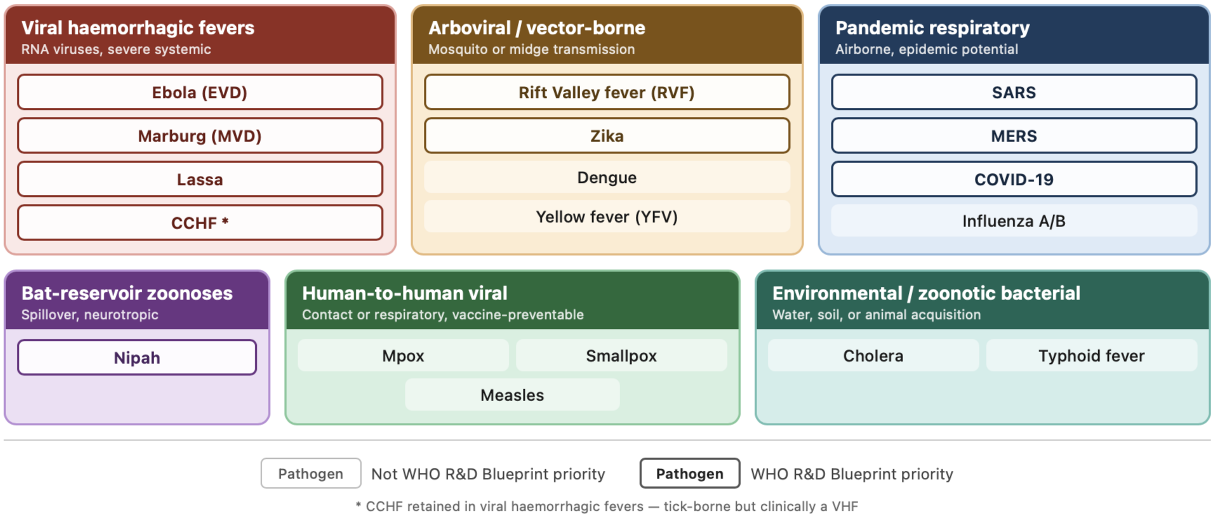

**Supplementary Figure S2.** Grouping of the 18 pathogens included in the analysis by epidemiological category. Groups are defined by shared transmission route and clinical presentation. Filled boxes indicate pathogens classified as WHO Research and Development Blueprint priorities. Asterisk denotes Crimean-Congo haemorrhagic fever, retained within the viral haemorrhagic fevers group on clinical grounds despite tick-borne transmission.

### Appendix A: Further methods

In the further methods section we give a brief description on the computation of the posterior predictive CDFs and how we evaluate the simulation study. The theoretical limitation of the generalised gamma distribution for summary-statistics data section details some of the limitations of the generalised gamma and under which circumstances it cannot be used (cases which have been excluded from the simulation study). We describe the joint order-statistics likelihood in full detail to complement the methods in the main text which describe the factorised likelihood.

#### A.1. Posterior predictive CDF

For each posterior draw, the predictive cumulative distribution function (CDF) was computed as a mixture over study-level random effects. Specifically, for each draw of the population-level parameters $(\mu_0, \tau, \phi)$, $L = 2000$ study-level location parameters were sampled from $\mathcal{N}(\mu_0, \tau)$ and the corresponding CDFs averaged to give a single mixture CDF for that draw. The shaded credible band represents the $2.5^{th}$–$97.5^{th}$ percentile range of these mixture CDFs across posterior draws. We note that for generalised gamma we use L = 200 to improve computation time required.

Reference lines indicating the predicted median and $95^{th}$ percentile of the incubation period were derived from the same posterior draws. For each draw, the x-value at which the mixture CDF crossed 0.5 (median) or 0.95 ($95^{th}$ percentile) was identified by linear interpolation; the dashed and dotted lines show the 95% prediction interval of these x-values across draws, and are therefore directly consistent with the shaded credible band.

When fewer than five studies contributed to the analysis, the between-study heterogeneity parameter $\tau$ cannot be reliably estimated from the data and is largely determined by its prior [41, 42]. In this setting, the predictive CDF was evaluated at the mean of the estimated study-level location parameters rather than sampling new locations from $\mathcal{N}(\mu_0, \tau)$, to avoid prior-dominated inflation of the predictive intervals.

#### A.2. Simulation study evaluation

***A.2.1. Weighted Interval Score.*** We assessed the calibration and sharpness of the posterior predictive distribution using the Weighted Interval Score (WIS) [49, 50]. The WIS is a proper scoring rule that simultaneously rewards calibrated coverage and narrow prediction intervals, and converges to the Continuous Ranked Probability Score (CRPS) as the number of intervals grows.

**Definition** Let $y$ be an observation drawn from the true marginal delay distribution, let $m$ be the predicted median, and let $\{(l_k, u_k)\}_{k=1}^{K}$ be a set of central prediction intervals at level $(1-\alpha_k) \times 100\%$ with lower and upper bounds $l_k$ and $u_k$ respectively. The Interval Score for a single interval at level $\alpha_k$ is

$$\mathrm{IS}_{\alpha_k}(l_k, u_k, y) = (u_k - l_k) + \frac{2}{\alpha_k}\max(l_k - y,\, 0) + \frac{2}{\alpha_k}\max(y - u_k,\, 0), \tag{21}$$

which penalises the interval width plus an additional penalty of $2/\alpha_k$ per unit by which $y$ falls outside the interval. The WIS across $K$ intervals is then

$$\mathrm{WIS}(m, \{l_k, u_k\}_{k=1}^{K}, y) = \frac{1}{K+0.5}\left[\frac{1}{2}|y-m| + \sum_{k=1}^{K}\frac{\alpha_k}{2}\,\mathrm{IS}_{\alpha_k}(l_k, u_k, y)\right]. \tag{22}$$

The $\frac{1}{2}|y-m|$ term is the score for the median forecast, and the normalising constant $K + 0.5$ places the WIS on the same scale as the Mean Absolute Error when $K = 0$.

**Prediction intervals used** We used $K = 4$ central prediction intervals at the 50%, 80%, 90%, and 95% levels, giving non-coverage probabilities $\alpha_k \in \{0.50,\, 0.20,\, 0.10,\, 0.05\}$ and a quantile grid of

$$\{0.025,\ 0.05,\ 0.10,\ 0.25,\ 0.50,\ 0.75,\ 0.90,\ 0.95,\ 0.975\}. \tag{23}$$

**Posterior predictive quantiles** The predicted quantiles $(m, l_k, u_k)$ are posterior predictive quantiles of the delay for a new study drawn from the fitted hierarchical model. They represent the full marginal distribution of a delay observation, integrating over both the study-specific random effect $\mu_d \sim \mathcal{N}(\mu_0, \tau^2)$ and the within-study variation.

For each simulation replicate we drew $B = 500$ posterior parameter samples $(\hat{\mu}_0^{(b)}, \hat{\tau}^{(b)}, \hat{\phi}^{(b)})$ from the Stan model. For each sample we drew $M = 1{,}000$ delay values from the marginal predictive distribution by first sampling a study location $\mu_d^{(b,m)} \sim \mathcal{N}(\hat{\mu}_0^{(b)}, (\hat{\tau}^{(b)})^2)$ and then sampling a delay from the chosen parametric distribution with location $\mu_d^{(b,m)}$ and shape $\hat{\phi}^{(b)}$. The predictive quantile at probability $p$ was taken as the median across posterior samples of the empirical $p$-quantile of the $M$ delay draws, yielding a single set of predicted quantiles that is robust to Monte Carlo noise.

**Evaluation against the true marginal distribution** To evaluate the score, we drew $n_{\text{test}} = 200$ test observations from the true marginal delay distribution used to generate each simulation scenario. Each test observation was obtained by first sampling a study-specific location $\mu_d \sim \mathcal{N}(\mu_0, \tau^2)$ using the true hyperparameters and then sampling a delay from the true parametric distribution, thus matching the population that the predictive distribution targets. The WIS was computed for each test observation and averaged:

$$\overline{\text{WIS}} = \frac{1}{n_{\text{test}}} \sum_{i=1}^{n_{\text{test}}} \text{WIS}\big(m, \{l_k, u_k\}_{k=1}^{K}, y_i\big) . \tag{24}$$

**Relative WIS** To enable comparison across scenarios with different delay scales, we also report the *relative WIS*, defined as

$$\text{relWIS} = \frac{\overline{\text{WIS}}}{\tilde{y}}, \tag{25}$$

where $\tilde{y}$ is the true marginal median of the delay distribution under the scenario's generating parameters. A perfect forecast has $\text{relWIS} = 0$; values below 1 indicate that the mean absolute prediction error is smaller than the median delay.

**Reported summaries** For each scenario (100 simulation replicates) we report the median $\overline{\text{WIS}}$ and median relWIS across replicates. Replicates flagged by a heuristic check (see Section A.3) are excluded from these summaries.

#### A.3. Theoretical limitations of the generalised gamma for summary-statistic data

The generalised gamma (GG) distribution in the Prentice parameterisation has three parameters: a log-location $\mu \in \mathbb{R}$, a log-scale $\sigma > 0$, and a shape parameter $Q \neq 0$, with $\gamma = 1/Q^2$. Its cumulative distribution function is

$$F(x;\, \mu, \sigma, Q) = I\big(\gamma\, e^{Qw};\, \gamma\big)\,, \qquad w = \frac{\log x - \mu}{\sigma}, \tag{26}$$

where $I(\cdot\,;\gamma)$ denotes the regularised incomplete gamma function. Three scenarios that illustrate fundamental identifiability constraints are discussed below. Because each limitation follows directly from the structure of the model and the information content of the available summaries, simulation evidence would add little beyond what can be established theoretically; these scenarios are therefore excluded from the simulation study.

***A.3.1. Very few studies ($D = 3$).*** In the hierarchical model, individual study log-location parameters $\delta_d$ are drawn from $\mathcal{N}(\mu_0, \tau^2)$. Estimating the between-study heterogeneity $\tau$ reliably requires enough studies to observe variation in $\delta_d$; as a practical guideline we require $D \geq 5$ before treating $\tau$ as data-identified rather than prior-dominated.

For the GG this constraint is more acute than for two-parameter distributions. The likelihood contribution of a single study is determined by at most a few order statistics (minimum, median, maximum for a typical study), and the Fisher information for $Q$ from a single order statistic of a sample of size $n$ is $O(1/n)$ but is additionally attenuated because $Q$ enters the likelihood only through the gamma-CDF argument $\gamma\, e^{Qw}$, a non-linear function whose sensitivity to $Q$ is low near $Q = 1$ (the Weibull boundary) and near $Q \to 0$ (the log-normal boundary). With $D = 3$ studies, the Fisher information for $Q$ accumulated across studies is insufficient to shift the marginal posterior for $Q$ appreciably away from its prior, so the posterior for $Q$ is essentially prior-determined regardless of the observed summaries. This identifiability failure is a structural consequence of the GG likelihood rather than an exceptional edge case, though its severity varies across three-parameter families. The Burr Type XII is also a three-parameter distribution but avoids this specific failure mode. Its tail-index parameter $\kappa$ enters the survival function as a direct power-law exponent whose sensitivity does not vanish at special boundary values, so the Fisher information for $\kappa$ from upper-tail order statistics remains non-negligible even when $D$ is small.

***A.3.2. Small per-study sample size ($N = 5$).*** When each study contributes only $N = 5$ observations, the study is summarised by its minimum $X_{(1)}$, median $X_{(3)}$, and maximum $X_{(5)}$. These correspond to the population quantiles at approximately the 17th, 50th, and 83rd percentiles.

The shape parameter $Q$ of the GG controls the behaviour of the distribution in the extreme tails — it governs how the density behaves as $x \to 0^+$ and $x \to \infty$ relative to the lognormal. For $N = 5$, the available order statistics span only a central $\sim 66\%$ of the distribution, a range over which the GG and the lognormal (or Weibull) are largely indistinguishable for realistic parameter values. Formally, the asymptotic variance of the $k$-th order statistic from a sample of size $n$ is

$$\mathrm{Var}\big(X_{(k)}\big) \approx \frac{p(1-p)}{n\left[f(F^{-1}(p))\right]^2}, \qquad p = \frac{k}{n+1}. \tag{27}$$

For $N = 5$ this variance is large enough that the likelihood surface is essentially flat in $Q$: changes in $Q$ of order $0.5$–$1.0$ produce differences in the expected order statistics that lie well within one standard error of those statistics. The within-study information about $Q$ is therefore negligible, and the hierarchical pooling across studies cannot recover a parameter that the data do not contain.

***A.3.3. Mean and standard deviation summaries only (summary type 3).*** If each study reports only its sample mean $\bar{x}$ and standard deviation $s$, the likelihood is constructed from these two moment summaries. The raw moments of the GG are

$$\mathbb{E}[X^r] = \mathrm{e}^{r\mu} \cdot \frac{\Gamma(\gamma + r\sigma/Q)}{\Gamma(\gamma)}, \tag{28}$$

so the mean and variance are

$$\mathbb{E}[X] = \mathrm{e}^{\mu} \cdot \frac{\Gamma(\gamma + \sigma/Q)}{\Gamma(\gamma)}, \tag{29}$$

$$\mathrm{Var}(X) = \mathrm{e}^{2\mu} \left[\frac{\Gamma(\gamma + 2\sigma/Q)}{\Gamma(\gamma)} - \left(\frac{\Gamma(\gamma + \sigma/Q)}{\Gamma(\gamma)}\right)^2\right]. \tag{30}$$

These two equations involve all three parameters $(\mu, \sigma, Q)$. However, the squared coefficient of variation $\mathrm{CV}^2 = \mathrm{Var}(X)/[\mathbb{E}(X)]^2$ simplifies to a function of $\sigma/Q$ and $\gamma = 1/Q^2$ only:

$$\mathrm{CV}^2 = \frac{\Gamma(\gamma + 2\sigma/Q)\,\Gamma(\gamma)}{\Gamma(\gamma + \sigma/Q)^2} - 1. \tag{31}$$

Given observed $(\bar{x},\, s)$, the CV is fixed, which constrains $(\sigma, Q)$ to a one-dimensional curve; the mean then determines $\mu$ as a function of the chosen point on that curve. The system is therefore *underdetermined by one degree of freedom*: there exists an entire manifold of parameter triples $(\mu, \sigma, Q)$ consistent with any observed (mean, SD) pair. The third parameter $Q$ is structurally non-identifiable from two moment summaries alone, and its posterior will simply reflect the prior.

#### A.4. Joint order-statistic likelihood

***Definition.*** Studies reporting a median with range (summary type 1) or interquartile range (summary type 2) contribute three order statistics drawn from the same underlying sample of $n$ observations – the minimum $x_{(i_1)}$, a central quantile $x_{(i_2)}$, and either the maximum or upper quartile $x_{(i_3)}$, with $x_{(i_1)} < x_{(i_2)} < x_{(i_3)}$. Because all three statistics arise from the same sample, they are positively correlated, such that knowledge of the minimum is informative about the median and knowledge of the median is informative about the spread. The likelihood for three jointly observed order statistics from $n$ i.i.d. draws from a distribution with density $f(\,\cdot\,; \boldsymbol{\theta})$ and CDF $F(\,\cdot\,; \boldsymbol{\theta})$ is

$$\mathcal{L}_{\text{joint}}(\boldsymbol{\theta}) \propto \frac{n!}{(i_1-1)!\,(i_2-i_1-1)!\,(i_3-i_2-1)!\,(n-i_3)!}$$
$$\times F\big(x_{(i_1)}\big)^{i_1-1} \cdot f\big(x_{(i_1)}\big)$$
$$\times \big[F\big(x_{(i_2)}\big) - F\big(x_{(i_1)}\big)\big]^{i_2-i_1-1} \cdot f\big(x_{(i_2)}\big)$$
$$\times \big[F\big(x_{(i_3)}\big) - F\big(x_{(i_2)}\big)\big]^{i_3-i_2-1} \cdot f\big(x_{(i_3)}\big)$$
$$\times \big[1 - F\big(x_{(i_3)}\big)\big]^{n-i_3}. \tag{32}$$

Reading from left to right, the four power terms represent the probability that $i_1 - 1$ unobserved observations fall below the smallest reported statistic, the probability that $i_2 - i_1 - 1$ fall in the gap between the first and second reported statistics, the probability that $i_3 - i_2 - 1$ fall in the gap between the second and third, and the probability that $n - i_3$ fall above the largest. Any term with a zero exponent equals one and is omitted in the implementation to avoid evaluating $0 \times (-\infty)$ during automatic differentiation. For numerical stability, each gap probability is computed on the log scale as $\log[F(x_{(j)}) - F(x_{(i)})] = \texttt{log\_diff\_exp}(\log F(x_{(j)}), \log F(x_{(i)}))$.

***The factorised likelihood as an approximation.*** The primary analysis uses a factorised likelihood in which each of the three reported order statistics contributes its marginal density independently. For the $k$-th order statistic from a sample of $n$, this marginal is

$$\mathcal{L}_{\text{marg}}(x_{(k)};\, \boldsymbol{\theta}) \propto \binom{n}{k} f\big(x_{(k)}\big)\ F\big(x_{(k)}\big)^{k-1}\ \big[1 - F\big(x_{(k)}\big)\big]^{n-k}, \tag{33}$$

and the factorised likelihood is $\mathcal{L}_{\text{fact}} = \prod_j \mathcal{L}_{\text{marg}}(x_{(i_j)})$. Although each factor in equation Eq. (33) is the correct marginal density for a single order statistic, their product ignores the positive correlation induced by the shared sample from which all three statistics were drawn. Multiplying the marginals is equivalent to treating the minimum, median, and maximum (or quartiles) as if they had been observed from three separate, independent samples of size $n$, an assumption that inflates the effective information content of the data and can produce posterior distributions that are too narrow.

The joint and factorised likelihoods are proportional (and therefore yield identical posteriors) only when all gap exponents are simultaneously zero. For summary type 1, this requires $k_{\text{med}} = 2$ and $n = 3$; for all larger sample sizes the two likelihoods differ, with the discrepancy increasing with $n$.

***Advantages and disadvantages.*** The joint likelihood is statistically correct; it uses the full dependence structure of the three reported order statistics and therefore produces better-calibrated posterior uncertainty. In terms of raw computation per HMC leapfrog step, the joint model requires seven distribution-function evaluations per type-1/2 dataset, compared with nine for the factorised model (three log-pdfs, three log-CDFs, and one log-CCDF versus three of each), so the per-step arithmetic cost is marginally lower. The two additional operations in the joint model (a normalising-constant evaluation via log-gamma functions and two log-difference-of-CDF evaluations for the gap terms) are negligible by comparison.

The principal computational disadvantage of the joint model lies in its effect on posterior geometry rather than arithmetic cost. The gap terms $[F(x_{(i_2)}) - F(x_{(i_1)})]^{i_2-i_1-1}$ approach zero, and their logarithm approaches $-\infty$, when two reported statistics are closely spaced relative to the distribution scale or when the exponent $i_2 - i_1 - 1$ is large. Either situation produces steep likelihood gradients that force the HMC sampler to adopt a smaller leapfrog step size, which increases the number of gradient evaluations required per effective sample. The factorised model couples statistics only through the shared parameters and therefore has a smoother posterior surface. In practice, the joint model is expected to run within approximately twice the wall-clock time of the factorised model for the datasets in this analysis; a worst-case slowdown of three to four times is plausible for pathogens with large reported sample sizes and tightly clustered summary statistics. A substantially smaller mean leapfrog step size in the sampler diagnostics is the empirical signature of this effect.

The factorised model is simpler to implement, more robust to atypical data configurations, and easier to extend to additional summary types. Its principal limitation is the systematic overcounting of information described above.

***Situations in which the two likelihoods differ most.*** The magnitude of the discrepancy between the joint and factorised likelihoods is governed by the gap exponents. For summary type 1, these are $k_{\text{med}} - 2$ (gap between minimum and median) and $n - k_{\text{med}} - 1$ (gap between median and maximum), where $k_{\text{med}} = \lfloor (n+1)/2 \rfloor$. Both exponents grow linearly with $n$, so the two likelihoods agree closely for small samples ($n \leq 5$, where each exponent is at most two) and diverge substantially for large ones ($n \geq 30$, where exponents exceed thirteen). Analogous expressions hold for summary type 2, with $k_{Q_1}$, $k_{\text{med}}$, and $k_{Q_3}$ replacing $i_1$, $i_2$, and $i_3$.

Beyond sample size, the likelihoods also diverge when reported statistics span a narrow range relative to the distribution scale. In this setting, the gap probability $F(x_{(i_2)}) - F(x_{(i_1)})$ is small, the corresponding gap term imposes a tight constraint on the shape parameter, and the joint likelihood places substantially less weight on parameter configurations that cannot reproduce the narrow spread than the factorised model does. Conversely, when the reported statistics span most of the distribution's support (as is common for heavy-tailed distributions or small samples) the gap probabilities are close to their expected values under the marginals and the two approaches yield similar posterior summaries.

Datasets contributing summary types 3 (mean and standard deviation), 4 (exact frequency table), and 5 (interval-censored frequency table) are unaffected; the likelihood formulations are identical for those summary types in both models.

### Appendix B: Further results

#### B.1. Simulation study sensitivity analysis

**Supplementary Table S1.** Simulation study performance by distribution family and summary type. Coverage values are empirical 95% credible interval coverage (%; nominal target: 95%). Predictive bias is the median signed error (posterior median − true value) across replicates. IQD: integrated quadratic distance; WIS: weighted interval score (lower is better). Metrics are averaged over numbers of datasets and within-study sample sizes.

| | | Parameter coverage (%) | | | Predictive coverage (%) | | Predictive bias (days) | | Scoring rules | |
|---|---|---|---|---|---|---|---|---|---|---|
| Summary type | $N$ | $\mu_0$ | $\tau$ | $\phi$ | P50 | P95 | P50 | P95 | IQD | WIS |
| **Log-normal** | | | | | | | | | | |
| Median + Range | 14 | 98.8 | 85.6 | 93.5 | 98.8 | 94.2 | -0.14 | +2.48 | 0.007 | 1.82 |
| Median + IQR | 12 | 99.0 | 82.0 | 40.4 | 98.8 | 96.6 | -0.04 | +2.14 | 0.007 | 1.83 |
| Mean + SD | 12 | 98.1 | 83.9 | 89.8 | 98.3 | 96.0 | -0.25 | +2.47 | 0.008 | 1.83 |
| Freq. table | 2 | 100.0 | 68.0 | 94.0 | 34.5 | 31.5 | +0.13 | -4.09 | 0.023 | 2.05 |
| Mixed | 14 | 97.3 | 88.6 | 89.1 | 97.2 | 95.1 | -0.02 | +1.15 | 0.002 | 1.95 |
| **Gamma** | | | | | | | | | | |
| Median + Range | 14 | 98.7 | 82.3 | 82.0 | 98.6 | 97.0 | -0.09 | +1.97 | 0.004 | 2.08 |
| Mean + SD | 12 | 98.7 | 79.7 | 70.2 | 98.8 | 96.9 | -0.05 | +2.18 | 0.004 | 2.07 |
| Freq. table | 2 | 100.0 | 69.0 | 85.5 | 41.5 | 33.0 | +0.37 | -4.42 | 0.012 | 2.31 |
| Mixed | 14 | 97.5 | 89.3 | 74.2 | 97.3 | 96.9 | +0.04 | +0.72 | 0.001 | 2.19 |
| **Weibull** | | | | | | | | | | |
| Median + Range | 14 | 98.3 | 84.9 | 90.8 | 98.1 | 96.1 | -0.16 | +2.09 | 0.004 | 1.84 |
| Median + IQR | 12 | 98.5 | 75.9 | 30.8 | 97.8 | 97.1 | +0.06 | +1.52 | 0.004 | 1.86 |
| Mean + SD | 12 | 98.7 | 84.2 | 94.6 | 98.5 | 97.2 | -0.26 | +1.67 | 0.004 | 1.85 |
| Freq. table | 2 | 100.0 | 74.5 | 94.0 | 47.5 | 30.5 | +0.27 | -3.94 | 0.010 | 2.02 |
| Mixed | 14 | 96.7 | 90.3 | 86.9 | 96.6 | 95.6 | +0.04 | +0.87 | 0.001 | 1.85 |
| **Burr XII** | | | | | | | | | | |
| Median + Range | 26 | 98.2 | 86.2 | 95.7 | 98.5 | 94.8 | -0.09 | +1.36 | 0.007 | 1.30 |
| Median + IQR | 4 | 98.5 | 86.2 | 47.2 | 98.8 | 97.8 | +0.01 | +0.66 | 0.003 | 1.29 |
| Mean + SD | 1 | 100.0 | 85.0 | 100.0 | 99.0 | 95.0 | -0.15 | +0.82 | 0.005 | 1.23 |
| Freq. table | 4 | 97.8 | 90.3 | 97.5 | 99.0 | 95.0 | -0.05 | +1.39 | 0.005 | 1.31 |
| Mixed | 6 | 95.3 | 84.3 | 88.3 | 98.7 | 96.0 | -0.04 | +0.67 | 0.003 | 1.33 |
| **Gen. Gamma** | | | | | | | | | | |
| Mixed | 13 | 97.3 | 86.7 | 94.8 | 98.6 | 95.6 | -0.15 | +1.67 | 0.003 | 1.81 |

*Note:* Coverage: empirical proportion of replicates where the true value fell inside the 95% posterior credible interval (nominal target: 95%). Predictive bias: median signed error (posterior median − true value) across replicates. IQD: integrated quadratic distance between true and estimated predictive

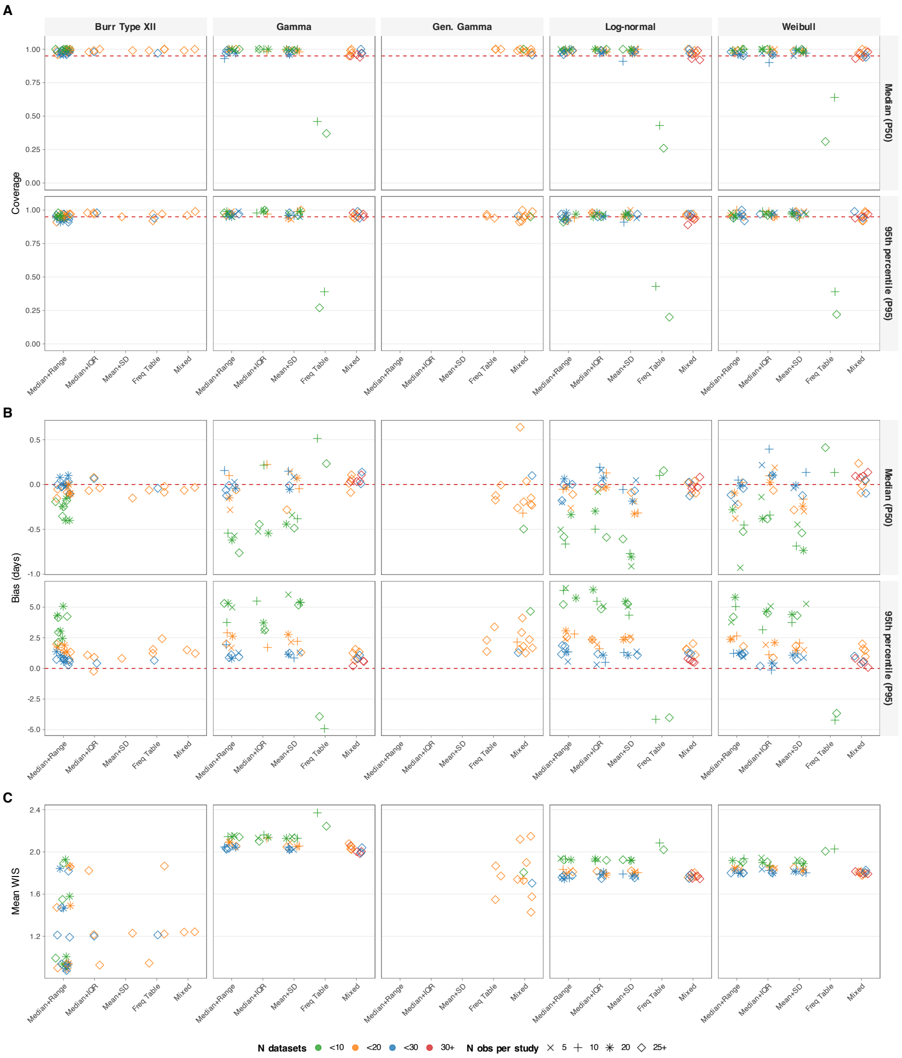

**Supplementary Figure S3. Supplementary simulation study performance across distribution families, summary-statistic types, number of contributing datasets, and within-study sample size.** Each point represents one scenario, summarised across 100 simulation replicates. Columns correspond to the five parametric families evaluated (Burr Type XII, gamma, generalised gamma, log-normal, Weibull); points on the x-axis indicate the summary-statistic type available to the model. Point colour denotes the number of contributing datasets and point shape denotes the within-study sample size, as indicated in the legend. Dashed red lines indicate the nominal target (0.95 for coverage panels; 0 for bias panels). (A) Empirical 95% credible interval coverage for the predictive median (P50; top row) and 95th percentile (P95; bottom row). (B) Median signed bias (days) for the predictive median (P50; top row) and 95th percentile (P95; bottom row). (C) Mean Weighted Interval Score (WIS). Generalised gamma scenarios only shown for frequency tables and mixed data summaries including frequency tables.

CDFs. WIS: weighted interval score (lower is better for IQD and WIS). Gen. Gamma scenarios with fewer than 10 identifiable replicates excluded prior to aggregation.

**Supplementary Table S2.** Simulation study performance by summary type, number of datasets, and within-study sample size (pooled over distribution families). Coverage values are empirical 95% credible interval coverage (%; nominal target: 95%). Predictive bias is the median signed error (posterior median − true value) across replicates. IQD: integrated quadratic distance; WIS: weighted interval score (lower is better).

| | | | Parameter coverage (%) | | | Predictive coverage (%) | | Predictive bias (days) | | Scoring rules | |
|---|---|---|---|---|---|---|---|---|---|---|---|
| $N$ datasets | $n$ obs | $N$ | $\mu_0$ | $\tau$ | $\phi$ | P50 | P95 | P50 | P95 | IQD | WIS |
| **Median + Range** | | | | | | | | | | | |
| <10 | 5 | 3 | 100.0 | 72.2 | 81.0 | 99.7 | 95.6 | -0.57 | +4.98 | 0.012 | 1.99 |
| <10 | 10 | 3 | 100.0 | 79.0 | 85.7 | 100.0 | 96.3 | -0.54 | +5.05 | 0.011 | 2.00 |

*continued on next page*

*continued from previous page*

| | | | Parameter coverage (%) | | | Predictive coverage (%) | | Predictive bias (days) | | Scoring rules | |
|---|---|---|---|---|---|---|---|---|---|---|---|
| $N$ datasets | $n$ obs | $N$ | $\mu_0$ | $\tau$ | $\phi$ | P50 | P95 | P50 | P95 | IQD | WIS |
| <10 | 20 | 7 | 99.4 | 78.3 | 95.1 | 99.6 | 95.4 | -0.34 | +5.07 | 0.014 | 1.63 |
| <10 | ≥25 | 7 | 99.9 | 82.7 | 95.6 | 99.7 | 95.1 | -0.35 | +4.20 | 0.013 | 1.62 |
| <20 | 5 | 3 | 98.7 | 82.3 | 80.0 | 99.0 | 97.3 | -0.28 | +2.40 | 0.004 | 1.92 |
| <20 | 10 | 3 | 99.0 | 84.3 | 85.7 | 99.0 | 97.0 | +0.02 | +2.81 | 0.004 | 1.92 |
| <20 | 20 | 7 | 98.3 | 85.4 | 92.7 | 99.0 | 95.4 | -0.09 | +1.89 | 0.005 | 1.56 |
| <20 | ≥25 | 7 | 99.1 | 88.4 | 95.0 | 98.7 | 95.0 | -0.11 | +1.80 | 0.004 | 1.55 |
| <30 | 5 | 3 | 97.7 | 83.3 | 82.0 | 98.0 | 96.3 | -0.20 | +0.95 | 0.002 | 1.88 |
| <30 | 10 | 3 | 97.0 | 85.7 | 85.7 | 96.3 | 93.7 | +0.05 | +1.35 | 0.002 | 1.87 |
| <30 | 20 | 7 | 97.3 | 89.6 | 92.0 | 97.0 | 94.3 | -0.00 | +1.07 | 0.002 | 1.53 |
| <30 | ≥25 | 15 | 97.3 | 89.9 | 94.9 | 97.5 | 95.5 | -0.02 | +0.94 | 0.002 | 1.62 |
| **Median + IQR** | | | | | | | | | | | |
| <10 | 5 | 2 | 100.0 | 68.5 | 27.0 | 100.0 | 97.5 | -0.11 | +5.08 | 0.010 | 1.94 |
| <10 | 10 | 2 | 100.0 | 75.0 | 25.0 | 100.0 | 98.5 | -0.42 | +4.32 | 0.009 | 1.89 |
| <10 | 20 | 2 | 100.0 | 74.9 | 89.5 | 100.0 | 96.0 | -0.34 | +5.54 | 0.010 | 1.92 |
| <10 | ≥25 | 2 | 100.0 | 76.5 | 81.5 | 100.0 | 95.5 | -0.49 | +4.67 | 0.010 | 1.91 |
| <20 | 5 | 2 | 99.5 | 73.0 | 5.0 | 100.0 | 97.5 | +0.07 | +1.94 | 0.004 | 1.83 |
| <20 | 10 | 2 | 98.5 | 82.0 | 6.0 | 97.5 | 98.0 | +0.11 | +1.36 | 0.004 | 1.83 |
| <20 | 20 | 2 | 99.0 | 84.5 | 66.5 | 99.0 | 96.5 | -0.00 | +2.22 | 0.003 | 1.82 |
| <20 | ≥25 | 5 | 97.8 | 86.4 | 53.2 | 98.6 | 97.2 | -0.05 | +0.91 | 0.004 | 1.52 |
| <30 | 5 | 2 | 97.5 | 66.0 | 0.0 | 97.0 | 98.0 | +0.19 | +0.28 | 0.003 | 1.80 |
| <30 | 10 | 2 | 98.0 | 81.0 | 0.0 | 93.5 | 96.0 | +0.29 | +0.18 | 0.003 | 1.81 |
| <30 | 20 | 2 | 98.0 | 84.0 | 31.0 | 97.0 | 96.0 | +0.09 | +0.76 | 0.002 | 1.80 |
| <30 | ≥25 | 3 | 98.0 | 92.3 | 38.0 | 97.7 | 96.7 | +0.07 | +0.42 | 0.002 | 1.58 |
| **Mean + SD** | | | | | | | | | | | |
| <10 | 5 | 3 | 99.7 | 75.3 | 78.3 | 99.7 | 97.7 | -0.45 | +5.27 | 0.011 | 2.00 |
| <10 | 10 | 3 | 99.7 | 76.3 | 87.0 | 99.7 | 97.7 | -0.69 | +4.34 | 0.011 | 1.98 |
| <10 | 20 | 3 | 99.7 | 75.3 | 91.3 | 99.3 | 97.7 | -0.74 | +5.40 | 0.011 | 1.98 |
| <10 | ≥25 | 3 | 99.7 | 79.3 | 97.0 | 99.7 | 95.3 | -0.54 | +5.17 | 0.010 | 1.99 |
| <20 | 5 | 3 | 99.7 | 84.3 | 70.7 | 99.7 | 97.3 | -0.19 | +2.15 | 0.004 | 1.91 |
| <20 | 10 | 3 | 99.3 | 83.7 | 86.0 | 99.0 | 96.7 | -0.16 | +2.21 | 0.004 | 1.90 |
| <20 | 20 | 3 | 98.0 | 83.7 | 91.3 | 98.0 | 95.7 | -0.24 | +2.35 | 0.004 | 1.89 |
| <20 | ≥25 | 4 | 98.8 | 82.8 | 94.5 | 98.8 | 97.2 | -0.22 | +1.57 | 0.004 | 1.74 |
| <30 | 5 | 3 | 97.3 | 80.0 | 63.0 | 98.0 | 95.7 | +0.05 | +1.28 | 0.002 | 1.87 |
| <30 | 10 | 3 | 96.0 | 90.7 | 80.7 | 95.7 | 94.3 | +0.14 | +1.18 | 0.002 | 1.88 |
| <30 | 20 | 3 | 97.7 | 89.0 | 87.0 | 97.3 | 97.3 | -0.05 | +1.08 | 0.002 | 1.87 |
| <30 | ≥25 | 3 | 97.0 | 91.3 | 93.3 | 98.0 | 97.3 | -0.07 | +1.08 | 0.001 | 1.86 |
| **Freq. table** | | | | | | | | | | | |
| <10 | 10 | 3 | 100.0 | 70.0 | 88.3 | 51.0 | 40.3 | +0.13 | -4.22 | 0.017 | 2.16 |
| <10 | ≥25 | 3 | 100.0 | 71.0 | 94.0 | 31.3 | 23.0 | +0.23 | -3.93 | 0.013 | 2.09 |
| <20 | ≥25 | 3 | 98.3 | 89.0 | 97.3 | 99.7 | 95.3 | -0.06 | +1.55 | 0.006 | 1.34 |
| <30 | ≥25 | 1 | 96.0 | 94.0 | 98.0 | 97.0 | 94.0 | -0.04 | +0.65 | 0.002 | 1.21 |
| **Mixed** | | | | | | | | | | | |
| <10 | ≥25 | 1 | 100.0 | 82.3 | 92.4 | 100.0 | 94.9 | -0.50 | +4.65 | 0.009 | 1.81 |
| <20 | 10 | 1 | 98.3 | 86.4 | 98.3 | 100.0 | 94.9 | -0.32 | +2.15 | 0.004 | 1.75 |
| <20 | ≥25 | 20 | 97.8 | 88.4 | 88.6 | 98.2 | 95.6 | -0.05 | +1.55 | 0.003 | 1.77 |
| <30 | ≥25 | 27 | 97.2 | 87.3 | 87.9 | 97.8 | 96.5 | +0.00 | +0.74 | 0.001 | 2.00 |
| ≥30 | ≥25 | 12 | 95.0 | 91.3 | 77.5 | 95.3 | 94.8 | +0.06 | +0.56 | 0.001 | 1.86 |

*Note:* Coverage: empirical proportion of replicates where the true value fell inside the 95% posterior credible interval (nominal target: 95%). Predictive bias: median signed error (posterior median − true value) across replicates. IQD: integrated quadratic distance between true and estimated predictive CDFs. WIS: weighted interval score (lower is better for IQD and WIS). Gen. Gamma scenarios with fewer than 10 identifiable replicates excluded prior to aggregation.

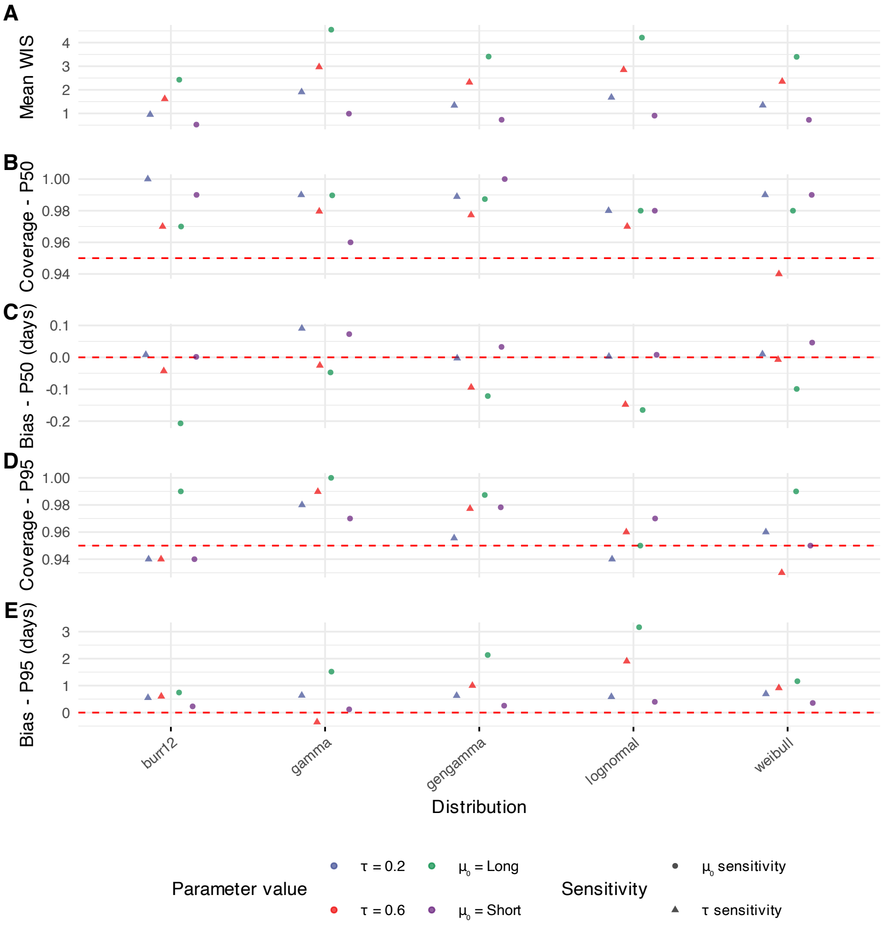

**Supplementary Figure S4.** Enter Caption

##### B.2. Comparison of factorised and joint likelihood

**Supplementary Table S3.** Comparison of posterior summaries under the factorised and joint order-statistic likelihoods (*filtered* analysis). Each pair of rows corresponds to one pathogen–distribution combination; the upper row gives results under the factorised model and the lower row under the joint model. $N$: total number of datasets; $n_{12}$: datasets contributing a type-1 (median + range) or type-2 (median + IQR) summary, i.e. those for which the two likelihoods differ. All posterior summaries are reported as median (2.5%ile, 97.5%ile). $\tau$ is shown as $--$ when $N < 5$ (prior-dominated). $\Delta$: joint minus factorised posterior median of the predictive quantity (days). † No type-1/2 datasets present; the two likelihoods are identical and the joint row repeats the factorised estimates.

| | | | | | Posterior summary — median (95% CI) | | | | | Δ (joint − fact.) | |
|---|---|---|---|---|---|---|---|---|---|---|---|
| Pathogen | Distribution | $N$ | $n_{12}$ | Model | Median (days) | $p_{95}$ (days) | $\mu_0$ | $\phi$ | $\tau$ | Median | $p_{95}$ |
| **Marburg (MVD)** | Log-normal | 2 | 2 | Factorised | 8.3 (7.8, 8.8) | 15.5 (13.8, 18.1) | 2.12 (0.98, 3.27) | 0.38 (0.31, 0.48) | — (n<5) | | |
| | | | | Joint | 8.3 (7.7, 8.9) | 15.5 (13.8, 18.0) | 2.13 (0.97, 3.29) | 0.38 (0.32, 0.46) | — (n<5) | +0.05 | +0.02 |
| | Gamma | 2 | 2 | Factorised | 8.4 (7.9, 8.9) | 14.6 (13.0, 16.5) | 1.95 (0.96, 3.27) | 7.61 (5.10, 12.35) | — (n<5) | | |
| | | | | Joint | 8.4 (7.8, 9.0) | 14.8 (13.4, 16.6) | 2.16 (1.00, 3.32) | 7.23 (5.12, 10.16) | — (n<5) | +0.01 | +0.25 |
| | Weibull | 2 | 2 | Factorised | 8.6 (8.1, 9.2) | 14.0 (12.9, 15.6) | 2.23 (1.06, 3.34) | 3.01 (2.46, 3.62) | — (n<5) | | |
| | | | | Joint | 8.6 (8.0, 9.3) | 14.1 (12.9, 15.6) | 2.24 (1.05, 3.36) | 2.99 (2.48, 3.58) | — (n<5) | -0.01 | +0.04 |
| | Burr XII | 2 | 2 | Factorised | 8.5 (8.0, 9.0) | 14.3 (12.8, 16.5) | 2.41 (1.18, 3.55) | 3.68 (2.88, 4.78) | — (n<5) | | |
| | | | | Joint | 8.5 (7.9, 9.1) | 14.4 (13.0, 16.6) | 2.40 (1.16, 3.51) | 3.64 (2.90, 4.66) | — (n<5) | +0.02 | +0.16 |
| **Lassa fever** | Log-normal | 1 | 0 | Factorised | 10.7 (9.2, 12.0) | 13.2 (11.6, 16.4) | 2.36 (0.90, 3.79) | 0.13 (0.07, 0.25) | — (n<5) | | |
| | | | | Joint† | 10.7 (9.2, 12.0) | 13.2 (11.6, 16.4) | 2.36 (0.90, 3.79) | 0.13 (0.07, 0.25) | — (n<5) | — | — |
| | Gamma | 1 | 0 | Factorised | 10.5 (9.0, 12.0) | 13.6 (11.8, 16.3) | 2.35 (0.92, 3.82) | 37.21 (15.79, 89.96) | — (n<5) | | |
| | | | | Joint† | 10.5 (9.0, 12.0) | 13.6 (11.8, 16.3) | 2.35 (0.92, 3.82) | 37.21 (15.79, 89.96) | — (n<5) | — | — |
| | Weibull | 1 | 0 | Factorised | 10.7 (9.3, 12.0) | 12.7 (11.4, 15.2) | 2.38 (0.89, 3.82) | 8.60 (4.26, 14.76) | — (n<5) | | |
| | | | | Joint† | 10.7 (9.3, 12.0) | 12.7 (11.4, 15.2) | 2.38 (0.89, 3.82) | 8.60 (4.26, 14.76) | — (n<5) | — | — |
| | Burr XII | 1 | 0 | Factorised | 10.7 (9.3, 12.0) | 12.9 (11.4, 15.9) | 2.43 (0.93, 3.89) | 10.45 (5.15, 21.96) | — (n<5) | | |
| | | | | Joint† | 10.7 (9.3, 12.0) | 12.9 (11.4, 15.9) | 2.43 (0.93, 3.89) | 10.45 (5.15, 21.96) | — (n<5) | — | — |
| | Gen. gamma | 1 | 0 | Factorised | 10.7 (9.2, 12.0) | 12.6 (11.4, 15.2) | 2.38 (0.89, 3.86) | 0.11 (0.05, 0.23) | — (n<5) | | |
| | | | | Joint† | 10.7 (9.2, 12.0) | 12.6 (11.4, 15.2) | 2.38 (0.89, 3.86) | 0.11 (0.05, 0.23) | — (n<5) | — | — |
| **Zika** | Log-normal | 2 | 1 | Factorised | 6.0 (5.2, 6.8) | 9.9 (8.5, 11.9) | 1.93 (0.86, 3.21) | 0.31 (0.25, 0.39) | — (n<5) | | |
| | | | | Joint | 6.0 (5.2, 6.8) | 9.8 (8.4, 11.7) | 1.94 (0.87, 3.18) | 0.30 (0.25, 0.37) | — (n<5) | +0.04 | -0.10 |
| | Gamma | 2 | 1 | Factorised | 6.0 (5.4, 7.0) | 9.5 (8.4, 11.5) | 2.02 (0.96, 2.73) | 11.58 (7.14, 14.70) | — (n<5) | | |
| | | | | Joint | 6.1 (5.3, 7.0) | 9.7 (8.4, 11.5) | 1.96 (0.92, 3.22) | 10.70 (7.30, 15.92) | — (n<5) | +0.05 | +0.19 |
| **Measles** | Log-normal | 11 | 6 | Factorised | 14.0 (12.3, 16.5) | 20.5 (18.0, 24.2) | 2.62 (2.47, 2.76) | 0.23 (0.21, 0.25) | 0.20 (0.11, 0.41) | | |
| | | | | Joint | 14.0 (12.3, 16.7) | 20.4 (17.9, 24.4) | 2.61 (2.47, 2.76) | 0.23 (0.21, 0.25) | 0.21 (0.11, 0.43) | -0.04 | -0.07 |
| | Gamma | 11 | 6 | Factorised | 13.8 (13.4, 14.2) | 20.3 (19.5, 21.0) | 2.65 (2.62, 2.67) | 16.46 (15.47, 17.14) | 0.08 (0.05, 0.12) | | |
| | | | | Joint | 14.0 (12.4, 16.7) | 20.1 (17.7, 24.0) | 2.64 (2.50, 2.79) | 18.76 (15.97, 21.86) | 0.20 (0.11, 0.42) | +0.18 | -0.20 |
| | Weibull | 11 | 6 | Factorised | 14.3 (12.7, 16.7) | 20.0 (17.8, 23.4) | 2.73 (2.60, 2.86) | 4.37 (4.01, 4.74) | 0.18 (0.10, 0.39) | | |
| | | | | Joint | 14.1 (12.5, 16.5) | 19.8 (17.5, 23.1) | 2.71 (2.58, 2.85) | 4.36 (4.01, 4.73) | 0.18 (0.09, 0.38) | -0.19 | -0.25 |
| | Burr XII | 11 | 6 | Factorised | 14.0 (12.3, 16.7) | 20.1 (17.7, 24.1) | 2.65 (2.48, 2.83) | 7.38 (6.12, 9.04) | 0.20 (0.11, 0.42) | | |
| | | | | Joint | 14.0 (12.3, 16.6) | 20.1 (17.6, 24.0) | 2.65 (2.48, 2.82) | 7.43 (6.14, 9.06) | 0.20 (0.11, 0.42) | -0.02 | -0.03 |

*continued on next page*

*continued from previous page*

| | | | | | Posterior summary — median (95% CI) | | | | | Δ (joint − fact.) | |
|---|---|---|---|---|---|---|---|---|---|---|---|
| Pathogen | Distribution | $N$ | $n_{12}$ | Model | Median (days) | $p_{95}$ (days) | $\mu_0$ | $\phi$ | $\tau$ | Median | $p_{95}$ |
| **Mpox** | Log-normal | 16 | 10 | Factorised | 7.3 (6.3, 8.8) | 19.4 (16.7, 23.4) | 1.95 (1.80, 2.11) | 0.59 (0.56, 0.63) | 0.27 (0.17, 0.46) | | |
| | | | | Joint | — | — | — | — | — | — | — |
| | Gamma | 16 | 10 | Factorised | 7.6 (6.6, 9.1) | 17.9 (15.5, 21.4) | 2.11 (1.96, 2.26) | 3.00 (2.67, 3.35) | 0.26 (0.17, 0.45) | | |
| | | | | Joint | — | — | — | — | — | — | — |
| | Weibull | 16 | 10 | Factorised | 7.8 (6.8, 9.3) | 17.3 (15.0, 20.8) | 2.22 (2.07, 2.37) | 1.84 (1.72, 1.95) | 0.27 (0.17, 0.44) | | |
| | | | | Joint | — | — | — | — | — | — | — |
| | Burr XII | 16 | 10 | Factorised | 7.6 (6.7, 9.0) | 17.6 (15.3, 20.9) | 2.70 (2.37, 3.11) | 2.18 (1.97, 2.42) | 0.26 (0.17, 0.43) | | |
| | | | | Joint | — | — | — | — | — | — | — |
| **Rift Valley fever** | Log-normal | 2 | 1 | Factorised | 3.9 (3.6, 4.3) | 5.4 (4.8, 6.3) | 1.36 (0.24, 2.50) | 0.19 (0.14, 0.27) | — (n<5) | | |
| | | | | Joint | — | — | — | — | — | — | — |
| | Gamma | 2 | 1 | Factorised | 4.0 (3.8, 4.2) | 5.8 (5.5, 6.2) | 1.08 (0.31, 2.34) | 16.92 (13.80, 20.80) | — (n<5) | | |
| | | | | Joint | — | — | — | — | — | — | — |
| | Weibull | 2 | 1 | Factorised | 4.1 (3.7, 4.5) | 5.5 (5.0, 6.2) | 1.44 (0.28, 2.56) | 4.98 (3.57, 6.63) | — (n<5) | | |
| | | | | Joint | — | — | — | — | — | — | — |
| | Burr XII | 2 | 1 | Factorised | 3.9 (3.6, 4.3) | 5.2 (4.7, 6.1) | 1.42 (0.27, 2.55) | 8.66 (5.46, 13.94) | — (n<5) | | |
| | | | | Joint | — | — | — | — | — | — | — |
| **Yellow fever** | Log-normal | 3 | 0 | Factorised | 3.5 (3.1, 4.0) | 5.7 (4.9, 7.1) | 1.27 (0.37, 2.16) | 0.29 (0.22, 0.39) | — (n<5) | | |
| | | | | Joint† | 3.5 (3.1, 4.0) | 5.7 (4.9, 7.1) | 1.27 (0.37, 2.16) | 0.29 (0.22, 0.39) | — (n<5) | — | — |
| | Gamma | 3 | 0 | Factorised | 3.6 (3.2, 4.0) | 5.4 (4.8, 6.4) | 1.31 (0.40, 2.22) | 13.79 (8.17, 22.22) | — (n<5) | | |
| | | | | Joint† | 3.6 (3.2, 4.0) | 5.4 (4.8, 6.4) | 1.31 (0.40, 2.22) | 13.79 (8.17, 22.22) | — (n<5) | — | — |
| | Weibull | 3 | 0 | Factorised | 3.7 (3.3, 4.1) | 5.4 (4.9, 6.2) | 1.39 (0.47, 2.26) | 3.97 (2.88, 5.23) | — (n<5) | | |
| | | | | Joint† | 3.7 (3.3, 4.1) | 5.4 (4.9, 6.2) | 1.39 (0.47, 2.26) | 3.97 (2.88, 5.23) | — (n<5) | — | — |
| | Burr XII | 3 | 0 | Factorised | 3.7 (3.2, 4.1) | 5.4 (4.8, 6.5) | 1.50 (0.57, 2.37) | 5.07 (3.53, 7.26) | — (n<5) | | |
| | | | | Joint† | 3.7 (3.2, 4.1) | 5.4 (4.8, 6.5) | 1.50 (0.57, 2.37) | 5.07 (3.53, 7.26) | — (n<5) | — | — |
| | Gen. gamma | 3 | 0 | Factorised | 3.7 (3.2, 4.2) | 5.4 (4.8, 6.3) | 1.37 (0.45, 2.25) | 0.26 (0.19, 0.36) | — (n<5) | | |
| | | | | Joint† | 3.7 (3.2, 4.2) | 5.4 (4.8, 6.3) | 1.37 (0.45, 2.25) | 0.26 (0.19, 0.36) | — (n<5) | — | — |
| **SARS** | Log-normal | 19 | 8 | Factorised | 4.6 (4.0, 5.3) | 11.3 (9.8, 13.2) | 1.49 (1.34, 1.62) | 0.55 (0.51, 0.58) | 0.26 (0.17, 0.42) | | |
| | | | | Joint | 4.6 (4.0, 5.4) | 11.2 (9.8, 13.2) | 1.49 (1.34, 1.63) | 0.55 (0.51, 0.58) | 0.26 (0.17, 0.42) | +0.00 | -0.02 |
| | Gamma | 19 | 8 | Factorised | 4.8 (4.2, 5.6) | 10.8 (9.4, 12.6) | 1.63 (1.49, 1.77) | 3.33 (2.95, 3.76) | 0.27 (0.18, 0.42) | | |
| | | | | Joint | 4.8 (4.2, 5.6) | 10.8 (9.4, 12.7) | 1.63 (1.49, 1.78) | 3.34 (2.96, 3.76) | 0.27 (0.17, 0.43) | +0.00 | -0.01 |
| | Weibull | 19 | 8 | Factorised | 5.0 (4.3, 5.9) | 10.7 (9.3, 12.6) | 1.75 (1.60, 1.90) | 1.91 (1.78, 2.04) | 0.28 (0.19, 0.44) | | |
| | | | | Joint | 4.9 (4.3, 5.8) | 10.6 (9.2, 12.6) | 1.75 (1.60, 1.89) | 1.90 (1.78, 2.03) | 0.28 (0.18, 0.45) | -0.03 | -0.07 |
| | Burr XII | 19 | 8 | Factorised | 4.6 (4.0, 5.4) | 10.7 (9.3, 12.5) | 1.58 (1.37, 1.87) | 3.15 (2.60, 3.80) | 0.27 (0.18, 0.42) | | |
| | | | | Joint | 4.6 (4.0, 5.4) | 10.7 (9.2, 12.5) | 1.58 (1.37, 1.86) | 3.15 (2.62, 3.79) | 0.27 (0.18, 0.43) | -0.01 | -0.02 |
| **Nipah** | Log-normal | 11 | 9 | Factorised | 9.1 (7.3, 12.4) | 14.3 (11.4, 19.6) | 2.14 (1.89, 2.38) | 0.27 (0.24, 0.31) | 0.36 (0.22, 0.66) | | |
| | | | | Joint | 9.1 (7.3, 12.5) | 14.2 (11.4, 19.6) | 2.14 (1.90, 2.39) | 0.27 (0.24, 0.31) | 0.36 (0.21, 0.66) | +0.00 | -0.04 |

*continued on next page*

*continued from previous page*

| | | | | | Posterior summary — median (95% CI) | | | | | Δ (joint − fact.) | |
|---|---|---|---|---|---|---|---|---|---|---|---|
| Pathogen | Distribution | $N$ | $n_{12}$ | Model | Median (days) | $p_{95}$ (days) | $\mu_0$ | $\phi$ | $\tau$ | Median | $p_{95}$ |
| | Gamma | 11 | 9 | Factorised | 8.9 (8.5, 15.0) | 13.8 (12.9, 22.5) | 2.16 (2.10, 2.36) | 14.16 (11.07, 14.67) | 0.33 (0.27, 0.86) | | |
| | | | | Joint | 9.2 (7.5, 12.2) | 13.8 (11.3, 18.5) | 2.18 (1.96, 2.41) | 14.25 (11.08, 18.04) | 0.34 (0.19, 0.62) | +0.24 | +0.02 |
| | Weibull | 11 | 9 | Factorised | 9.5 (7.8, 12.6) | 13.7 (11.1, 18.1) | 2.29 (2.06, 2.51) | 4.09 (3.53, 4.66) | 0.33 (0.19, 0.62) | | |
| | | | | Joint | 9.5 (7.8, 12.6) | 13.6 (11.1, 17.9) | 2.29 (2.06, 2.51) | 4.13 (3.60, 4.71) | 0.32 (0.18, 0.61) | -0.04 | -0.11 |
| | Burr XII | 11 | 9 | Factorised | 9.2 (7.5, 12.3) | 13.4 (10.9, 18.0) | 2.31 (2.03, 2.61) | 5.72 (4.62, 7.24) | 0.34 (0.20, 0.63) | | |
| | | | | Joint | 9.2 (7.5, 12.0) | 13.4 (11.0, 17.7) | 2.32 (2.05, 2.61) | 5.68 (4.62, 7.13) | 0.32 (0.18, 0.61) | -0.02 | -0.00 |
| **MERS** | Log-normal | 10 | 7 | Factorised | 6.1 (4.9, 8.3) | 15.2 (12.1, 20.8) | 1.75 (1.50, 1.99) | 0.55 (0.51, 0.60) | 0.34 (0.19, 0.65) | | |
| | | | | Joint | 6.1 (5.0, 8.4) | 15.2 (12.2, 20.9) | 1.76 (1.52, 1.99) | 0.55 (0.51, 0.60) | 0.33 (0.18, 0.65) | +0.02 | +0.07 |
| | Gamma | 10 | 7 | Factorised | 6.4 (5.2, 8.6) | 13.5 (11.0, 18.3) | 1.89 (1.66, 2.12) | 3.99 (3.38, 4.68) | 0.31 (0.17, 0.62) | | |
| | | | | Joint | 6.4 (5.2, 8.5) | 13.4 (10.9, 18.1) | 1.89 (1.65, 2.11) | 3.96 (3.37, 4.64) | 0.31 (0.17, 0.62) | -0.03 | -0.06 |
| | Weibull | 10 | 7 | Factorised | 6.6 (5.5, 8.7) | 13.0 (10.7, 17.0) | 2.01 (1.79, 2.23) | 2.18 (1.99, 2.38) | 0.29 (0.16, 0.58) | | |
| | | | | Joint | 6.6 (5.4, 8.6) | 12.9 (10.6, 17.0) | 2.01 (1.79, 2.22) | 2.17 (1.98, 2.37) | 0.28 (0.15, 0.58) | -0.06 | -0.09 |
| | Burr XII | 10 | 7 | Factorised | 6.4 (5.2, 8.5) | 13.2 (10.7, 17.7) | 2.25 (1.85, 2.72) | 2.70 (2.34, 3.19) | 0.32 (0.18, 0.62) | | |
| | | | | Joint | 6.4 (5.2, 8.6) | 13.3 (10.7, 18.1) | 2.21 (1.82, 2.67) | 2.72 (2.35, 3.21) | 0.32 (0.17, 0.63) | -0.04 | +0.06 |
| **Ebola (EVD)** | Log-normal | 11 | 2 | Factorised | 8.5 (7.1, 11.0) | 20.6 (17.1, 26.7) | 2.10 (1.89, 2.31) | 0.54 (0.50, 0.57) | 0.29 (0.16, 0.57) | | |
| | | | | Joint | 8.5 (7.1, 10.9) | 20.6 (17.1, 26.5) | 2.10 (1.90, 2.30) | 0.54 (0.50, 0.57) | 0.28 (0.16, 0.55) | +0.01 | -0.00 |
| | Gamma | 11 | 2 | Factorised | 9.0 (7.5, 11.7) | 20.0 (16.6, 25.9) | 2.25 (2.05, 2.46) | 3.47 (3.03, 3.95) | 0.29 (0.16, 0.56) | | |
| | | | | Joint | 9.0 (7.5, 11.5) | 20.0 (16.6, 25.6) | 2.26 (2.05, 2.45) | 3.47 (3.02, 3.95) | 0.29 (0.16, 0.55) | +0.01 | +0.03 |
| | Weibull | 11 | 2 | Factorised | 9.3 (7.7, 12.1) | 19.8 (16.4, 25.7) | 2.37 (2.17, 2.58) | 1.93 (1.79, 2.07) | 0.30 (0.17, 0.57) | | |
| | | | | Joint | 9.2 (7.6, 11.8) | 19.7 (16.3, 25.2) | 2.37 (2.16, 2.57) | 1.93 (1.79, 2.07) | 0.29 (0.17, 0.56) | -0.03 | -0.06 |
| | Burr XII | 11 | 2 | Factorised | 8.9 (7.4, 11.4) | 19.5 (16.2, 24.9) | 2.51 (2.20, 2.92) | 2.67 (2.28, 3.18) | 0.28 (0.16, 0.55) | | |
| | | | | Joint | 8.9 (7.4, 11.4) | 19.4 (16.2, 24.8) | 2.51 (2.19, 2.90) | 2.67 (2.29, 3.18) | 0.28 (0.16, 0.54) | -0.03 | -0.05 |
| **Smallpox** | Log-normal | 4 | 0 | Factorised | 12.3 (11.9, 12.6) | 16.5 (15.8, 17.3) | 2.51 (1.85, 3.16) | 0.18 (0.16, 0.20) | — (n<5) | | |
| | | | | Joint† | 12.3 (11.9, 12.6) | 16.5 (15.8, 17.3) | 2.51 (1.85, 3.16) | 0.18 (0.16, 0.20) | — (n<5) | — | — |
| | Gamma | 4 | 0 | Factorised | 12.3 (12.0, 12.7) | 16.3 (15.6, 17.0) | 2.53 (1.87, 3.19) | 32.15 (25.11, 40.51) | — (n<5) | | |
| | | | | Joint† | 12.3 (12.0, 12.7) | 16.3 (15.6, 17.0) | 2.53 (1.87, 3.19) | 32.15 (25.11, 40.51) | — (n<5) | — | — |
| | Weibull | 4 | 0 | Factorised | 12.6 (12.1, 13.0) | 16.1 (15.6, 16.7) | 2.59 (1.90, 3.23) | 5.95 (5.24, 6.70) | — (n<5) | | |
| | | | | Joint† | 12.6 (12.1, 13.0) | 16.1 (15.6, 16.7) | 2.59 (1.90, 3.23) | 5.95 (5.24, 6.70) | — (n<5) | — | — |
| | Burr XII | 4 | 0 | Factorised | 12.5 (12.1, 12.8) | 16.0 (15.4, 16.9) | 2.64 (1.96, 3.29) | 8.23 (6.80, 10.08) | — (n<5) | | |
| | | | | Joint† | 12.5 (12.1, 12.8) | 16.0 (15.4, 16.9) | 2.64 (1.96, 3.29) | 8.23 (6.80, 10.08) | — (n<5) | — | — |
| | Gen. gamma | 4 | 0 | Factorised | 12.4 (11.9, 13.0) | 16.0 (15.2, 17.0) | 2.55 (1.88, 3.20) | 0.17 (0.15, 0.20) | — (n<5) | | |
| | | | | Joint† | 12.4 (11.9, 13.0) | 16.0 (15.2, 17.0) | 2.55 (1.88, 3.20) | 0.17 (0.15, 0.20) | — (n<5) | — | — |
| **CCHF** | Log-normal | 11 | 4 | Factorised | 5.8 (3.9, 10.6) | 15.5 (10.5, 28.3) | 1.54 (1.13, 1.96) | 0.60 (0.55, 0.66) | 0.64 (0.41, 1.08) | | |
| | | | | Joint | 5.8 (3.9, 10.7) | 15.4 (10.4, 28.8) | 1.54 (1.12, 1.97) | 0.60 (0.55, 0.65) | 0.65 (0.42, 1.10) | +0.02 | -0.03 |
| | Gamma | 11 | 4 | Factorised | 6.0 (4.1, 10.9) | 15.0 (10.2, 27.3) | 1.72 (1.32, 2.14) | 2.61 (2.12, 3.16) | 0.63 (0.40, 1.07) | | |

*continued on next page*

*continued from previous page*

| | | | | | Posterior summary — median (95% CI) | | | | | Δ (joint − fact.) | |
|---|---|---|---|---|---|---|---|---|---|---|---|
| Pathogen | Distribution | $N$ | $n_{12}$ | Model | Median (days) | $p_{95}$ (days) | $\mu_0$ | $\phi$ | $\tau$ | Median | $p_{95}$ |
| | | | | Joint | 6.1 (4.1, 11.1) | 14.9 (10.2, 27.3) | 1.72 (1.31, 2.14) | 2.67 (2.19, 3.21) | 0.64 (0.41, 1.08) | +0.05 | -0.03 |
| | Weibull | 11 | 4 | Factorised | 6.1 (4.2, 11.0) | 14.9 (10.2, 26.6) | 1.83 (1.43, 2.24) | 1.65 (1.47, 1.84) | 0.63 (0.40, 1.06) | | |
| | | | | Joint | 6.1 (4.1, 11.4) | 14.7 (9.9, 27.5) | 1.82 (1.39, 2.24) | 1.67 (1.49, 1.85) | 0.65 (0.42, 1.10) | +0.01 | -0.16 |
| | Burr XII | 11 | 4 | Factorised | 5.9 (4.0, 10.7) | 14.6 (9.9, 26.6) | 1.82 (1.30, 2.47) | 2.55 (1.95, 3.44) | 0.63 (0.41, 1.07) | | |
| | | | | Joint | 5.9 (4.0, 10.8) | 14.5 (9.8, 26.6) | 1.78 (1.28, 2.40) | 2.64 (2.03, 3.51) | 0.64 (0.41, 1.08) | -0.01 | -0.11 |
| **Typhoid** | Log-normal | 22 | 4 | Factorised | 13.7 (11.4, 17.2) | 27.9 (23.1, 35.1) | 2.53 (2.33, 2.72) | 0.43 (0.41, 0.45) | 0.43 (0.32, 0.62) | | |
| | | | | Joint | 13.8 (11.5, 17.3) | 28.0 (23.2, 35.2) | 2.53 (2.34, 2.72) | 0.43 (0.41, 0.45) | 0.43 (0.31, 0.62) | +0.04 | +0.12 |
| | Gamma | 22 | 4 | Factorised | 14.1 (11.8, 17.5) | 26.7 (22.4, 33.3) | 2.61 (2.43, 2.80) | 5.53 (5.05, 6.05) | 0.43 (0.31, 0.61) | | |
| | | | | Joint | 14.1 (11.8, 17.6) | 26.8 (22.3, 33.5) | 2.62 (2.43, 2.81) | 5.53 (5.05, 6.03) | 0.42 (0.31, 0.61) | +0.03 | +0.06 |
| | Weibull | 22 | 4 | Factorised | 14.2 (11.9, 17.7) | 26.5 (22.2, 33.0) | 2.72 (2.54, 2.91) | 2.35 (2.24, 2.46) | 0.41 (0.30, 0.60) | | |
| | | | | Joint | 14.2 (11.9, 17.5) | 26.4 (22.2, 32.6) | 2.72 (2.54, 2.90) | 2.35 (2.24, 2.46) | 0.40 (0.29, 0.58) | -0.06 | -0.13 |
| | Burr XII | 22 | 4 | Factorised | 13.9 (11.4, 17.8) | 29.5 (24.0, 38.2) | 2.40 (2.19, 2.62) | 5.23 (4.68, 5.85) | 0.46 (0.34, 0.66) | | |
| | | | | Joint | 13.9 (11.4, 17.7) | 29.5 (24.1, 38.1) | 2.40 (2.19, 2.61) | 5.20 (4.64, 5.82) | 0.46 (0.34, 0.66) | +0.01 | -0.00 |
| **Dengue** | Log-normal | 14 | 0 | Factorised | 6.3 (5.4, 7.8) | 8.8 (7.4, 10.9) | 1.79 (1.61, 1.98) | 0.20 (0.18, 0.22) | 0.31 (0.20, 0.52) | | |
| | | | | Joint† | 6.3 (5.4, 7.8) | 8.8 (7.4, 10.9) | 1.79 (1.61, 1.98) | 0.20 (0.18, 0.22) | 0.31 (0.20, 0.52) | — | — |
| | Gamma | 14 | 0 | Factorised | 6.4 (5.4, 7.9) | 8.7 (7.4, 10.9) | 1.82 (1.64, 2.00) | 24.73 (20.06, 30.19) | 0.30 (0.19, 0.51) | | |
| | | | | Joint† | 6.4 (5.4, 7.9) | 8.7 (7.4, 10.9) | 1.82 (1.64, 2.00) | 24.73 (20.06, 30.19) | 0.30 (0.19, 0.51) | — | — |
| | Weibull | 14 | 0 | Factorised | 6.4 (5.5, 7.8) | 8.7 (7.5, 10.7) | 1.90 (1.74, 2.07) | 4.80 (4.31, 5.32) | 0.27 (0.17, 0.47) | | |
| | | | | Joint† | 6.4 (5.5, 7.8) | 8.7 (7.5, 10.7) | 1.90 (1.74, 2.07) | 4.80 (4.31, 5.32) | 0.27 (0.17, 0.47) | — | — |
| | Burr XII | 14 | 0 | Factorised | 6.3 (5.4, 8.0) | 8.9 (7.4, 11.2) | 1.80 (1.59, 2.01) | 9.00 (7.17, 11.56) | 0.32 (0.20, 0.54) | | |
| | | | | Joint† | 6.3 (5.4, 8.0) | 8.9 (7.4, 11.2) | 1.80 (1.59, 2.01) | 9.00 (7.17, 11.56) | 0.32 (0.20, 0.54) | — | — |
| **Influenza** | Log-normal | 13 | 6 | Factorised | 2.6 (1.8, 4.2) | 4.6 (3.2, 7.6) | 0.74 (0.38, 1.10) | 0.35 (0.33, 0.38) | 0.63 (0.43, 0.99) | | |
| | | | | Joint | 2.6 (1.8, 4.2) | 4.6 (3.3, 7.6) | 0.74 (0.39, 1.09) | 0.35 (0.33, 0.38) | 0.63 (0.43, 1.00) | +0.00 | +0.00 |
| | Weibull | 13 | 6 | Factorised | 2.6 (1.9, 4.4) | 4.5 (3.2, 7.5) | 0.91 (0.55, 1.27) | 2.82 (2.63, 3.02) | 0.62 (0.42, 1.00) | | |
| | | | | Joint | 2.7 (1.9, 4.4) | 4.4 (3.2, 7.3) | 0.91 (0.56, 1.27) | 2.87 (2.68, 3.06) | 0.62 (0.42, 0.99) | +0.01 | -0.03 |
| | Burr XII | 13 | 6 | Factorised | 2.6 (1.8, 4.3) | 4.5 (3.2, 7.7) | 0.72 (0.35, 1.11) | 5.59 (4.68, 6.71) | 0.63 (0.43, 1.01) | | |
| | | | | Joint | 2.6 (1.8, 4.4) | 4.6 (3.2, 7.7) | 0.71 (0.34, 1.10) | 5.65 (4.74, 6.78) | 0.64 (0.43, 1.02) | +0.02 | +0.04 |
| **Cholera** | Log-normal | 13 | 2 | Factorised | 1.7 (1.3, 2.5) | 3.9 (2.9, 5.7) | 0.42 (0.13, 0.70) | 0.50 (0.46, 0.55) | 0.47 (0.29, 0.79) | | |
| | | | | Joint | 1.7 (1.3, 2.5) | 3.9 (2.9, 5.7) | 0.41 (0.12, 0.70) | 0.50 (0.46, 0.55) | 0.47 (0.29, 0.79) | -0.01 | -0.01 |
| | Gamma | 13 | 2 | Factorised | 1.8 (1.4, 2.6) | 3.7 (2.9, 5.4) | 0.55 (0.26, 0.83) | 4.08 (3.38, 4.88) | 0.46 (0.28, 0.78) | | |
| | | | | Joint | 1.8 (1.4, 2.6) | 3.7 (2.8, 5.4) | 0.55 (0.26, 0.83) | 4.07 (3.38, 4.88) | 0.46 (0.29, 0.79) | -0.00 | -0.00 |
| | Weibull | 13 | 2 | Factorised | 1.8 (1.4, 2.6) | 3.8 (2.9, 5.4) | 0.68 (0.39, 0.96) | 2.03 (1.83, 2.23) | 0.46 (0.29, 0.79) | | |
| | | | | Joint | 1.8 (1.4, 2.7) | 3.8 (2.9, 5.5) | 0.68 (0.38, 0.97) | 2.03 (1.83, 2.23) | 0.47 (0.29, 0.79) | +0.00 | -0.00 |
| | Burr XII | 13 | 2 | Factorised | 1.7 (1.3, 2.5) | 3.6 (2.7, 5.3) | 0.66 (0.30, 1.05) | 3.12 (2.60, 3.75) | 0.46 (0.29, 0.80) | | |
| | | | | Joint | 1.7 (1.3, 2.5) | 3.6 (2.7, 5.3) | 0.65 (0.28, 1.03) | 3.14 (2.62, 3.77) | 0.47 (0.29, 0.80) | -0.01 | -0.01 |

*continued on next page*

*continued from previous page*

| | | | | | Posterior summary — median (95% CI) | | | | | Δ (joint − fact.) | |
|---|---|---|---|---|---|---|---|---|---|---|---|
| Pathogen | Distribution | $N$ | $n_{12}$ | Model | Median (days) | $p_{95}$ (days) | $\mu_0$ | $\phi$ | $\tau$ | Median | $p_{95}$ |
| **COVID-19** | Log-normal | 44 | 18 | Factorised | 4.8 (4.2, 5.5) | 11.4 (10.0, 13.2) | 1.48 (1.35, 1.61) | 0.53 (0.52, 0.53) | 0.42 (0.33, 0.54) | | |
| | | | | Joint | 4.8 (4.2, 5.5) | 11.4 (10.0, 13.2) | 1.48 (1.35, 1.61) | 0.53 (0.52, 0.53) | 0.41 (0.33, 0.53) | +0.00 | +0.01 |
| | Gamma | 44 | 18 | Factorised | 5.1 (4.5, 5.9) | 10.7 (9.4, 12.4) | 1.62 (1.50, 1.75) | 4.02 (3.95, 4.09) | 0.42 (0.34, 0.54) | | |
| | | | | Joint | 5.1 (4.5, 5.9) | 10.7 (9.5, 12.5) | 1.63 (1.50, 1.76) | 4.01 (3.94, 4.08) | 0.42 (0.33, 0.54) | +0.02 | +0.04 |
| | Weibull | 44 | 18 | Factorised | 5.3 (4.6, 6.2) | 10.5 (9.2, 12.2) | 1.75 (1.62, 1.88) | 2.14 (2.12, 2.16) | 0.42 (0.34, 0.54) | | |
| | | | | Joint | 5.3 (4.7, 6.1) | 10.5 (9.2, 12.2) | 1.75 (1.62, 1.88) | 2.14 (2.12, 2.16) | 0.41 (0.33, 0.53) | +0.01 | +0.01 |
| | Burr XII | 44 | 18 | Factorised | 5.1 (4.5, 5.9) | 10.6 (9.3, 12.3) | 2.02 (1.88, 2.17) | 2.64 (2.58, 2.69) | 0.42 (0.34, 0.54) | | |
| | | | | Joint | 5.1 (4.5, 5.9) | 10.6 (9.4, 12.3) | 2.03 (1.89, 2.17) | 2.63 (2.58, 2.69) | 0.42 (0.33, 0.54) | -0.00 | +0.00 |

† No type-1/2 datasets; joint and factorised likelihoods are identical.

##### B.3. Comparison to classical meta-analysis

**Supplementary Table S4.** Classical meta-analysis of incubation period central estimates (random-effects model, log-transformed mean, `sm = MLN`). $k$ = number of studies. Pooled mean and 95% confidence interval (CI) are back-transformed to days. $I^2$ measures the percentage of total variance due to between-study heterogeneity. $\tau_{\text{meta}}$ is the between-study SD on the log scale (REML). $\tau_{\text{Stan}}$ (median and 95% credible interval) is from the best-fitting Bayesian model ("all data" slot), also on the log scale.

| Pathogen | $k$ | Pooled mean [95% CI] (days) | $I^2$ | $\tau_{\text{meta}}$ | $\tau_{\text{Stan}}$ [95% CrI] | Best model |
|---|---|---|---|---|---|---|
| *Viral haemorrhagic fevers* | | | | | | |
| Ebola (EVD) | 11 | 9.36 [8.44, 10.37] | 81.3% | 0.148 | 0.283 [0.159, 0.552] | Burr XII |
| CCHF | 11 | 5.29 [3.76, 7.44] | 94.8% | 0.546 | 0.634 [0.406, 1.072] | Burr XII |
| *Pandemic respiratory* | | | | | | |
| SARS | 22 | 4.99 [4.54, 5.48] | 88.5% | 0.192 | 0.256 [0.172, 0.392] | Burr XII |
| MERS | 11 | 6.55 [5.99, 7.16] | 75.2% | 0.119 | 0.287 [0.162, 0.551] | Gamma |
| COVID-19 | 50 | 5.28 [4.79, 5.83] | 99.3% | 0.345 | 0.429 [0.349, 0.538] | Gamma |
| Influenza | 14 | 2.09 [1.54, 2.84] | 99.2% | 0.579 | 0.672 [0.464, 1.042] | Log-normal |
| *Arboviral / vector-borne* | | | | | | |
| Dengue | 14 | 6.26 [5.58, 7.02] | 89.3% | 0.200 | 0.307 [0.196, 0.520] | Log-normal |
| *Bat-reservoir zoonoses* | | | | | | |
| Nipah | 11 | 9.30 [8.98, 9.62] | 61.6% | 0.016 | 0.332 [0.269, 0.855] | Gamma |
| *Human-to-human viral* | | | | | | |
| Measles | 12 | 13.94 [13.30, 14.61] | 84.6% | 0.073 | 0.182 [0.103, 0.360] | Log-normal |
| Mpox | 16 | 8.04 [7.28, 8.88] | 85.9% | 0.182 | 0.265 [0.169, 0.446] | Gamma |
| *Environmental / zoonotic bacterial* | | | | | | |
| Cholera | 16 | 1.83 [1.57, 2.13] | 90.7% | 0.280 | 0.376 [0.240, 0.621] | Burr XII |
| Typhoid | 22 | 13.58 [11.56, 15.95] | 97.8% | 0.375 | 0.461 [0.341, 0.663] | Burr XII |

### Appendix C: Pathogen specific data and results

In Tables S5 and S6 we provide an overview of the data collected from the literature. This corresponds to the data available in `ddsynth`. We also provide figures for the fitted distributions for each pathogen and figures displaying the underlying data used for inference. We also provide the forest plots for the meta-analysis, including sub-groups, for each pathogen.

**Supplementary Table S5.** Composition of incubation period datasets included in the analysis. For each pathogen (and subgroup where applicable), $k$ is the number of datasets and $N$ is the total number of individual observations. *Individual-level data* denotes datasets provided as frequency tables (exact or interval-censored); *summary statistics only* denotes datasets reported as mean ± SD, median ± IQR, or median ± range.

| | **Total** | | **Individual-level** | | **Summary statistics only** | |
|---|---|---|---|---|---|---|
| **Pathogen** | $k$ | $N$ | $k$ | $N$ | $k$ | $N$ |
| ***Viral haemorrhagic fevers*** | | | | | | |
| **Ebola (EVD)** | 11 | 575 | 4 | 146 | 7 | 429 |
| *non-West Africa* | 3 | 144 | 1 | 4 | 2 | 140 |
| *West Africa* | 8 | 431 | 3 | 142 | 5 | 289 |
| **Marburg (MVD)** | 2 | 142 | – | – | 2 | 142 |
| **Lassa fever** | 1 | 15 | 1 | 15 | – | – |
| **CCHF** | 11 | 315 | 4 | 30 | 7 | 285 |
| *tick-bite* | 3 | 78 | 1 | 12 | 2 | 66 |
| *Nosocomial* | 3 | 20 | 2 | 15 | 1 | 5 |
| *other* | 4 | 214 | – | – | 4 | 214 |
| *sexual-transmission* | 1 | 3 | 1 | 3 | – | – |
| ***Pandemic respiratory*** | | | | | | |
| **COVID-19** | 51 | 28,302 | 12 | 21,298 | 39 | 7,004 |
| *Wildtype* | 30 | 13,539 | 5 | 8,160 | 25 | 5,379 |
| *Omicron* | 10 | 3,788 | 2 | 2,759 | 8 | 1,029 |
| *Delta* | 7 | 5,328 | 2 | 4,742 | 5 | 586 |
| *Alpha* | 2 | 5,184 | 2 | 5,184 | – | – |
| *Beta* | 2 | 463 | 1 | 453 | 1 | 10 |
| **SARS** | 22 | 1,132 | 6 | 142 | 16 | 990 |
| *China* | 3 | 203 | 1 | 22 | 2 | 181 |
| *Hong Kong* | 5 | 461 | 2 | 78 | 3 | 383 |
| *Canada* | 6 | 215 | 1 | 8 | 5 | 207 |

*continued on next page*

*continued from previous page*

| Pathogen | Total k | Total N | Individual-level k | Individual-level N | Summary statistics only k | Summary statistics only N |
|---|---|---|---|---|---|---|
| *Taiwan* | 2 | 130 | – | – | 2 | 130 |
| *Singapore* | 4 | 85 | 1 | 15 | 3 | 70 |
| *Unclassified* | 2 | 38 | 1 | 19 | 1 | 19 |
| **MERS** | 11 | 547 | 2 | 122 | 9 | 425 |
| *Republic of Korea* | 7 | 286 | 1 | 99 | 6 | 187 |
| *Saudi Arabia* | 2 | 41 | 1 | 23 | 1 | 18 |
| *Unclassified* | 2 | 220 | – | – | 2 | 220 |
| **Influenza** | 14 | 951 | 7 | 334 | 7 | 617 |
| *H5N1* | 4 | 53 | 1 | 8 | 3 | 45 |
| *H2N2* | 1 | 16 | 1 | 16 | – | – |
| *Influenza A* | 2 | 98 | 2 | 98 | – | – |
| *H1N1* | 5 | 706 | 1 | 134 | 4 | 572 |
| *Influenza B* | 2 | 78 | 2 | 78 | – | – |
| ***Arboviral / vector-borne*** | | | | | | |
| **Dengue** | 14 | 204 | 14 | 204 | – | – |
| *Inoculation* | 7 | 63 | 7 | 63 | – | – |
| *mosquito bite* | 4 | 126 | 4 | 126 | – | – |
| *Unclassified* | 3 | 15 | 3 | 15 | – | – |
| **Zika** | 2 | 136 | 1 | 25 | 1 | 111 |
| **Rift Valley fever** | 2 | 18 | 1 | 12 | 1 | 6 |
| **Yellow fever** | 3 | 26 | 3 | 26 | – | – |
| *mosquito bite* | 2 | 18 | 2 | 18 | – | – |
| *Inoculation* | 1 | 8 | 1 | 8 | – | – |
| ***Bat-reservoir zoonoses*** | | | | | | |
| **Nipah** | 11 | 205 | 1 | 11 | 10 | 194 |
| *Bangladesh* | 7 | 139 | 1 | 11 | 6 | 128 |
| *Unclassified* | 4 | 66 | – | – | 4 | 66 |
| ***Human-to-human viral*** | | | | | | |
| **Mpox** | 16 | 1,161 | 4 | 195 | 12 | 966 |
| **Measles** | 12 | 373 | 4 | 195 | 8 | 178 |
| **Smallpox** | 4 | 131 | 4 | 131 | – | – |
| ***Environmental / zoonotic bacterial*** | | | | | | |
| **Cholera** | 16 | 386 | 13 | 334 | 3 | 52 |
| *O139* | 4 | 45 | 2 | 27 | 2 | 18 |
| *O1 Classical* | 6 | 144 | 6 | 144 | – | – |
| *O1 El Tor* | 3 | 123 | 2 | 89 | 1 | 34 |
| *O1 El Tor Ogawa* | 3 | 74 | 3 | 74 | – | – |
| **Typhoid** | 22 | 963 | 18 | 835 | 4 | 128 |
| *Other* | 9 | 211 | 8 | 199 | 1 | 12 |
| *Catered meal* | 4 | 180 | 4 | 180 | – | – |
| *Experimental* | 4 | 258 | 2 | 218 | 2 | 40 |
| *Water* | 4 | 291 | 3 | 215 | 1 | 76 |
| *Unclassified* | 1 | 23 | 1 | 23 | – | – |

**Supplementary Table S6.** Collected incubation period datasets by pathogen.

| Reference | Country | Subgroup | $n$ | Summary | DOI |
|---|---|---|---|---|---|
| ***EVD*** | | | | | |
| Wamala (2010) | Uganda | non-West Africa | 116 | Median: 7.0 (Range: 2.0–20.0) | 10.3201/eid1607.091525 |
| Francesconi (2003) | Uganda | non-West Africa | 24 | Median: 6.0 (Range: 1.0–16.0) | 10.3201/eid0911.030339 |
| Yan (2015) | Sierra Leone | West Africa | 33 | Mean: 9.2 (SD: 6.7) | 10.1007/s10096-015-2457-z |
| Yamin (2016) | Sierra Leone | West Africa | 20 | Mean: 8.6 (SD: 6.1) | 10.1016/j.ajic.2016.04.216 |
| Muoghalu (2017) | Sierra Leone | West Africa | 76 | Mean: 9.5 (SD: 4.0) | 10.3389/fpubh.2017.00160 |
| Faye (2015) | Guinea | West Africa | 152 | Mean: 9.9 (SD: 5.5) | 10.1016/S1473-3099(14)71075-8 |
| Ajelli (2015) | Sierra Leone | West Africa | 8 | Mean: 9.7 (SD: 3.7) | 10.1186/s12916-015-0524-z |
| Chan (2020) | Nigeria | non-West Africa | 4 | *Freq. table.* Median: 8.5 (Range: 8.0–12.0) | 10.1098/rsif.2020.0498 |
| WHO Ebola Response Team (2014) | Guinea | West Africa | 58 | *Freq. table.* Median: 10.0 (Range: 2.0–38.0) | 10.1056/NEJMoa1411100 |
| WHO Ebola Response Team (2014) | Liberia | West Africa | 54 | *Freq. table.* Median: 10.5 (Range: 1.0–42.0) | 10.1056/NEJMoa1411100 |
| WHO Ebola Response Team (2014) | Sierra Leone | West Africa | 30 | *Freq. table.* Median: 10.0 (Range: 3.0–18.0) | 10.1056/NEJMoa1411100 |
| ***MVD*** | | | | | |
| Pavlin (2014) | Mixed | — | 66 | Median: 7.0 (Range: 2.0–14.0) | 10.1186/1756-0500-7-906 |
| Nsanzimana (2025) | Rwanda | — | 76 | Median: 10.0 (IQR: 8.0–13.0) | 10.1056/NEJMoa2415816 |
| ***Lassa*** | | | | | |
| Carey (1972) | Nigeria | — | 15 | *Freq. table (interval-censored).* Range: 1.0–22.0 | 10.1016/0035-9203(72)90271-4 |
| ***CCHF*** | | | | | |
| Arslan (2024) | Turkey | tick-bite | 49 | Mean: 4.0 (SD: 2.4) | 10.14744/nci.2023.09815 |
| Fazlalipour (2024) | Iran | nosocomial | 12 | *Freq. table.* Median: 5.5 (Range: 1.0–22.0) | 10.1186/s12879-024-10199 |
| Kaya (2011) | Turkey | tick-bite | 12 | *Freq. table.* Median: 21.0 (Range: 13.0–53.0) | 10.1016/j.ijid.2011.03.007 |
| Naderi (2013) | Iran | nosocomial | 3 | *Freq. table.* Median: 5.0 (Range: 1.0–8.0) | 10.4269/ajtmh.2012.12-0337 |
| Beştepe Dursun (2021) | Turkey | other | 64 | Median: 3.0 (IQR: 2.0–4.0) | 10.1002/jca.21875 |
| Koksal (2010) | Turkey | other | 64 | Mean: 5.5 (SD: 3.6) | 10.1016/j.jcv.2009.11.007 |
| Koksal (2010) | Turkey | other | 72 | Mean: 4.9 (SD: 3.9) | 10.1016/j.jcv.2009.11.007 |
| Swanepoel (1989) | South Africa | tick-bite | 17 | Median: 2.5 (Range: 2.0–7.0) | 10.1093/clinids/11.Supplement_4.S794 |
| Swanepoel (1989) | South Africa | other | 14 | Median: 4.0 (Range: 2.0–9.0) | 10.1093/clinids/11.Supplement_4.S794 |
| Swanepoel (1989) | South Africa | nosocomial | 5 | Median: 6.0 (Range: 3.0–7.0) | 10.1093/clinids/11.Supplement_4.S794 |
| Pshenichnaya (2016) | Russia | sexual-transmission | 3 | *Freq. table (interval-censored).* Range: 2.0–9.0 | 10.1016/j.ijid.2016.02.1008 |
| ***SARS*** | | | | | |
| Wu (2003) | China | China | 96 | Mean: 5.9 (SD: 3.5) | — |
| Virlogeux (2015) | Hong Kong | Hong Kong | 234 | Mean: 4.7 (SD: 4.6) | 10.1097/ede.0000000000000339 |
| Varia (2003) | Canada | Canada | 42 | Median: 4.0 (Range: 2.0–10.0) | — |
| Wong (2004) | Hong Kong | Hong Kong | 11 | Median: 3.0 (Range: 2.0–6.0) | 10.3201/eid1002.030452 |
| Scales (2003) | Canada | Canada | 7 | Median: 5.0 (Range: 1.0–15.0) | 10.3201/eid0910.030525 |
| Meltzer (2004) | Mixed | — | 19 | Median: 4.0 (Range: 1.0–18.0) | 10.3201/eid1002.030426 |
| McBryde (2006) | China | China | 85 | Mean: 5.3 (SD: 4.5) | 10.1007/s11538-005-9005-4 |
| Liu (2016) | Taiwan | Taiwan | 98 | Median: 6.0 (Range: 1.0–15.0) | 10.1371/journal.pone.0149988 |
| Lee (2003) | Hong Kong | Hong Kong | 138 | Median: 6.0 (Range: 2.0–16.0) | 10.1056/NEJMoa030685 |
| Hsu (2003) | Singapore | Singapore | 7 | Median: 4.0 (Range: 2.0–8.0) | 10.3201/eid0906.030264 |
| Hsu (2003) | Singapore | Singapore | 13 | Median: 7.0 (Range: 4.0–12.0) | 10.3201/eid0906.030264 |
| Goh (2006) | Singapore | Singapore | 50 | Mean: 5.1 (SD: 2.2) | — |

continued on next page

continued from previous page

| Reference | Country | Subgroup | $n$ | Summary | DOI |
|---|---|---|---|---|---|
| Chen (2003) | Taiwan | Taiwan | 32 | Median: 4.0 (Range: 3.0–6.0) | 10.1001/archotol.129.11.1157 |
| Booth (2003) | Canada | Canada | 144 | Median: 6.0 (IQR: 3.0–10.0) | 10.1001/jama.289.21.JOC30885 |
| Avendano (2003) | Canada | Canada | 4 | Mean: 4.0 (SD: 3.0) | — |
| Avendano (2003) | Canada | Canada | 10 | Mean: 3.5 (SD: 3.0) | — |
| Meltzer (2004) | Mixed | — | 19 | *Freq. table (interval-censored).* Range: 1.0–18.0 | 10.3201/eid1002.030426 |
| Farewell (2005) | Hong Kong | Hong Kong | 67 | *Freq. table (interval-censored).* Range: 0.1–14.5 | 10.1002/sim.2206 |
| Chow (2004) | Singapore | Singapore | 15 | *Freq. table.* Median: 4.0 (Range: 3.0–8.0) | 10.1136/bmj.37939.465729.44 |
| Olsen (2003) | China | China | 22 | *Freq. table.* Median: 3.5 (Range: 2.0–8.0) | 10.1056/NEJMoa031349 |
| Wong (2004) | Hong Kong | Hong Kong | 11 | *Freq. table.* Median: 3.0 (Range: 2.0–6.0) | 10.3201/eid1002.030452 |
| Dwosh (2003) | Canada | Canada | 8 | *Freq. table (interval-censored).* Range: 1.0–12.0 | — |
| ***MERS*** | | | | | |
| Cho (2016) | Republic of Korea | Republic of Korea | 73 | Median: 7.0 (IQR: 5.0–10.0) | 10.1016/S0140-6736(16)30623-7 |
| Nam (2017) | Republic of Korea | Republic of Korea | 14 | Median: 8.0 (IQR: 6.5–10.5) | 10.1016/j.ijid.2017.02.008 |
| Nam (2017) | Republic of Korea | Republic of Korea | 11 | Median: 4.0 (IQR: 3.0–8.0) | 10.1016/j.ijid.2017.02.008 |
| Al-Jasser (2019) | Saudi Arabia | Saudi Arabia | 18 | Mean: 6.3 (SD: 4.3) | 10.1016/j.jiph.2018.09.008 |
| Kim (2015) | Republic of Korea | Republic of Korea | 36 | Median: 5.0 (Range: 2.0–13.0) | 10.1177/1010539515610036 |
| Kim (2016) | Republic of Korea | Republic of Korea | 17 | Median: 7.0 (Range: 2.0–14.0) | 10.1016/j.phrp.2016.01.001 |
| Park (2015) | Republic of Korea | Republic of Korea | 36 | Median: 6.0 (Range: 2.0–15.0) | 10.2807/1560-7917.es2015.20.25.21169 |
| Liu (2016) | Middle East | — | 92 | Median: 5.0 (Range: 2.0–15.0) | 10.1371/journal.pone.0149988 |
| Liu (2016) | Middle East | — | 128 | Median: 7.0 (Range: 3.0–11.0) | 10.1371/journal.pone.0149988 |
| Assiri (2013) | Saudi Arabia | Saudi Arabia | 23 | *Freq. table (interval-censored).* Range: 1.0–19.0 | 10.1056/NEJMoa1306742 |
| Virlogeux (2016) | Republic of Korea | Republic of Korea | 99 | *Freq. table (interval-censored).* Range: 0.1–27.0 | 10.3201/eid2203.151437 |
| ***COVID_19*** | | | | | |
| Shen (2020) | China | Wildtype | 8 | Median: 7.0 (Range: 4.0–12.0) | 10.1093/ofid/ofaa231 |
| Bender (2021) | Germany | Wildtype | 53 | Median: 4.3 (IQR: 2.5–6.5) | 10.3201/eid2704.204576 |
| Liu (2020) | China | Wildtype | 27 | Median: 6.0 (Range: 1.0–13.0) | 10.1097/JCMA.0000000000000411 |
| Song (2020) | China | Wildtype | 22 | Mean: 8.2 (SD: 3.6) | 10.1016/j.jinf.2020.04.018 |
| Ki (2020) | South Korea | Wildtype | 10 | Median: 3.0 (Range: 0.1–15.0) | 10.4178/epih.e2020007 |
| Mao (2020) | China | Wildtype | 28 | Median: 8.5 (Range: 1.0–24.0) | 10.1186/s12889-020-09606-4 |
| Zhang (2020) | China | Wildtype | 8 | Median: 7.0 (Range: 2.0–12.0) | 10.1186/s12879-020-05570-x |
| Zhang (2020) | China | Wildtype | 23 | Median: 8.0 (Range: 4.0–13.0) | 10.1186/s12879-020-05570-x |
| Zhang (2020) | China | Wildtype | 46 | Median: 10.0 (Range: 7.0–15.0) | 10.1186/s12879-020-05570-x |
| Guo (2020) | China | Wildtype | 85 | Median: 9.0 (Range: 6.0–13.0) | 10.1186/s12916-020-01719-2 |
| Nie (2020) | China | Wildtype | 2907 | Median: 5.0 (Range: 2.0–8.0) | 10.1093/infdis/jiaa211 |
| Du (2021) | China | Wildtype | 75 | Median: 8.5 (Range: 6.0–12.0) | 10.7883/yoken.JJID.2021.274 |
| Hua (2020) | China | Wildtype | 43 | Mean: 9.1 (SD: 3.7) | 10.1002/jmv.26180 |
| Wong (2020) | Brunei | Wildtype | 15 | Median: 5.0 (Range: 1.0–11.0) | 10.4269/ajtmh.20-0771 |
| Hu (2021) | China | Wildtype | 268 | Median: 5.7 (IQR: 3.2–8.8) | 10.1038/s41467-021-21710-6 |

continued on next page

continued from previous page

| **Reference** | **Country** | **Subgroup** | $n$ | **Summary** | **DOI** |
|---|---|---|---|---|---|
| Zhao (2021a), doi: 10.1016/j.epidem.2... | China | Wildtype | 254 | Mean: 6.8 (SD: 4.1) | 10.1016/j.epidem.2021.100482 |
| Böhm (2021) | Germany | Wildtype | 256 | *Freq. table (interval-censored).* Range: 0.1–43.0 | 10.1017/S0950268821000510 |
| Yang (2020) | China | Wildtype | 178 | Median: 5.4 (Range: 1.0–21.0) | 10.1017/S0950268820001338 |
| Bui (2020) | Vietnam | Wildtype | 19 | Median: 5.6 (Range: 1.4–13.0) | 10.1371/journal.pone.0243889 |
| Han (2020) | South Korea | Wildtype | 8 | Median: 5.5 (Range: 3.0–12.0) | 10.4178/epih.e2020056 |
| Kong (2020) | China | Wildtype | 136 | *Freq. table.* Median: 8.5 (Range: 1.0–17.0) | 10.1002/agm2.12114 |
| Xiao (2021) | China | Wildtype | 217 | *Freq. table.* Median: 8.0 (Range: 1.0–18.0) | 10.1186/s12199-021-00935-3 |
| Dai (2020) | China | Wildtype | 180 | Mean: 5.8 (SD: 3.7) | 10.2147/RMHP.S257907 |
| Huang (2020) | China | Wildtype | 6 | Median: 2.0 (Range: 1.0–4.0) | 10.1016/j.jinf.2020.03.006 |
| Zhao (2021b), doi: 10.1097/MD.0000000... | China | Wildtype | 102 | Mean: 6.5 (SD: 4.6) | 10.1097/MD.0000000000027846 |
| Li (2020) | China | Wildtype | 957 | Mean: 8.7 (SD: 5.2) | 10.3389/fpubh.2020.577431 |
| Bernal Lopez (2022) | UK | Wildtype | 45 | Median: 4.0 (Range: 0.1–11.0) | 10.2807/1560-7917.ES.2022.27.15.2001551 |
| Bernal Lopez (2022) | UK | Wildtype | 12 | Median: 4.0 (Range: 2.0–11.0) | 10.2807/1560-7917.ES.2022.27.15.2001551 |
| Mefsin (2022) | China | Omicron | 57 | Mean: 4.6 (SD: 1.7) | 10.3201/eid2809.220613 |
| Mefsin (2022) | China | Omicron | 23 | Mean: 4.4 (SD: 1.4) | 10.3201/eid2809.220613 |
| Brandal (2021) | Norway | Omicron | 81 | Median: 3.0 (Range: 0.1–8.0) | 10.2807/1560-7917.ES.2021.26.50.2101147 |
| Backer (2022) | Netherlands | Omicron | 258 | Mean: 3.2 (SD: 2.2) | 10.2807/1560-7917.ES.2022.27.6.2200042 |
| Tanaka (2022) | Japan | Omicron | 77 | *Freq. table.* Median: 3.0 (Range: 1.0–7.0) | 10.3390/ijerph19106330 |
| Liu (2022) | South Korea | Omicron | 22 | Mean: 3.5 (SD: 1.4) | 10.1016/j.onehlt.2022.100425 |
| Zeng (2023) | Singapore | Omicron | 36 | Median: 3.0 (IQR: 2.0–4.0) | 10.3201/eid2904.220854 |
| Xiong (2023) | China | Omicron | 500 | Mean: 3.3 (SD: 1.1) | 10.3201/eid2902.221243 |
| Wei (2023) | China | Omicron | 52 | Mean: 4.6 (SD: 2.1) | 10.1111/irv.13097 |
| Backer (2022) | Netherlands | Delta | 255 | Mean: 4.4 (SD: 2.5) | 10.2807/1560-7917.ES.2022.27.6.2200042 |
| McAleavey (2022) | Ireland | Delta | 171 | Median: 3.0 (Range: 1.0–13.0) | 10.1016/j.puhe.2022.06.023 |
| Zhang (2021) | China | Delta | 47 | Mean: 4.4 (SD: 1.9) | 10.46234/ccdcw2021.148 |
| Li (2022) | China | Delta | 136 | *Freq. table (interval-censored).* Range: 0.1–15.0 | 10.1371/journal.pntd.0010048 |
| Zeng (2023) | Singapore | Delta | 42 | Median: 4.0 (IQR: 3.0–7.0) | 10.3201/eid2904.220854 |
| Luo (2023) | China | Delta | 71 | Mean: 5.3 (SD: 3.4) | 10.46234/ccdcw2023.011 |
| Tanaka (2022) | Japan | Alpha | 51 | *Freq. table.* Median: 5.0 (Range: 1.0–12.0) | 10.3390/ijerph19106330 |
| Investigation team (2021) | France | Beta | 10 | Median: 4.5 (Range: 2.0–7.0) | 10.2807/1560-7917.ES.2021.26.13.2100333 |
| Böhmer (2020) | Germany | Wildtype | 12 | *Freq. table (interval-censored).* Range: 1.0–7.0 | 10.1016/S1473-3099(20)30314-5 |
| Galmiche (2023) | France | Alpha | 5133 | *Freq. table.* Median: 5.0 (Range: 1.0–15.0) | 10.1016/S2666-5247(23)00005-8 |
| Galmiche (2023) | France | Beta | 453 | *Freq. table.* Median: 5.0 (Range: 1.0–15.0) | 10.1016/S2666-5247(23)00005-8 |
| Galmiche (2023) | France | Delta | 4606 | *Freq. table.* Median: 4.0 (Range: 1.0–15.0) | 10.1016/S2666-5247(23)00005-8 |
| Galmiche (2023) | France | Omicron | 2682 | *Freq. table.* Median: 3.0 (Range: 1.0–13.0) | 10.1016/S2666-5247(23)00005-8 |
| Galmiche (2023) | France | Wildtype | 7539 | *Freq. table.* Median: 4.0 (Range: 1.0–15.0) | 10.1016/S2666-5247(23)00005-8 |
| ***Flu*** | | | | | |
| Beigel (2005) | Thailand | H5N1 | 17 | Median: 4.0 (Range: 2.0–8.0) | 10.1056/NEJMra052211 |
| Beigel (2005) | Vietnam | H5N1 | 10 | Median: 3.0 (Range: 2.0–4.0) | 10.1056/NEJMra052211 |
| Huai (2008) | China | H5N1 | 18 | Median: 5.0 (Range: 2.0–9.5) | 10.3201/eid1411.080509 |
| Oner (2006) | Turkey | H5N1 | 8 | *Freq. table.* Median: 4.5 (Range: 4.0–7.0) | 10.1056/NEJMoa060601 |
| Knight (1965) | USA | H2N2 | 16 | *Freq. table.* Median: 2.0 (Range: 1.0–3.0) | — |
| Moser (1979) | USA | Influenza A | 37 | *Freq. table (interval-censored).* Range: 1.0–4.0 | 10.1093/oxfordjournals.aje.a112781 |

continued on next page

continued from previous page

| Reference | Country | Subgroup | $n$ | Summary | DOI |
|---|---|---|---|---|---|
| Cao (2009) | China | H1N1 | 426 | Median: 2.0 (Range: 1.0–7.0) | 10.1056/NEJMoa0906612 |
| Shen (2012) | China | H1N1 | 23 | Median: 2.0 (IQR: 1.0–3.0) | 10.1186/1743-422X-9-20 |
| Wang (2012) | China | H1N1 | 79 | Median: 1.6 (Range: 0.4–4.2) | 10.1016/j.puhe.2011.11.008 |
| Lessler (2009) | USA | H1N1 | 134 | *Freq. table (interval-censored).* Range: 0.1–11.2 | 10.1056/NEJMoa0908481 |
| Canini (2011) | Mixed | H1N1 | 44 | Mean: 1.9 (SD: 0.7) | 10.1128/jvi.01318-10 |
| Henle (1946) | USA | Influenza A | 61 | *Freq. table (interval-censored).* Range: 0.5–2.5 | 10.4049/jimmunol.52.2.145 |
| Henle (1946) | USA | Influenza B | 24 | *Freq. table (interval-censored).* Range: 0.5–2.0 | 10.4049/jimmunol.52.2.145 |
| Francis (1944) | USA | Influenza B | 54 | *Freq. table (interval-censored).* Range: 0.5–1.0 | — |
| ***RVF*** | | | | | |
| Hoogstraal (1979) | Mixed | — | 12 | *Freq. table.* Median: 3.5 (Range: 3.0–6.0) | — |
| Archer (2011) | South Africa | — | 6 | Median: 4.0 (Range: 4.0–5.0) | 10.7196/samj.4544 |
| ***Zika*** | | | | | |
| Sharma (2019) | India | — | 111 | Median: 6.0 (Range: 2.0–10.0) | 10.4103/INJMS.INJMS_65_19 |
| Lessler (2016) | Mixed | — | 25 | *Freq. table (interval-censored).* Range: 0.0–185.0 | 10.2471/BLT.16.174540 |
| ***Dengue*** | | | | | |
| Ashburn & Craig 1907, https://www.jst... | Philippines | inoculation | 9 | *Freq. table.* Median: 3.5 (Range: 2.5–7.0) | — |
| Cleland (1919) | Australia | inoculation | 9 | *Freq. table.* Median: 7.0 (Range: 4.8–10.0) | 10.1017/S0022172400007476 |
| Cleland (1918) | Australia | inoculation | 16 | *Freq. table (interval-censored).* Range: 4.9–9.4 | 10.1017/S0022172400006690 |
| Siler (1926) | Philippines | mosquito bite | 47 | *Freq. table (interval-censored).* Range: 4.0–11.0 | — |
| Siler (1926) | Philippines | inoculation | 3 | *Freq. table.* Median: 6.8 (Range: 6.5–7.2) | — |
| Graham (1903) | Lebanon | inoculation | 3 | *Freq. table (interval-censored).* Range: 4.0–6.0 | — |
| Schule PA (1928) | Philippines | mosquito bite | 8 | *Freq. table (interval-censored).* Range: 5.0–20.0 | 10.4269/ajtmh.1928.s1-8.203 |
| Chandler (1923) | USA | mosquito bite | 4 | *Freq. table.* Median: 6.0 (Range: 4.1–6.5) | 10.4269/ajtmh.1923.s1-3.233 |
| Chandler (1923) | USA | inoculation | 2 | *Freq. table.* Median: 5.5 (Range: 5.1–5.8) | 10.4269/ajtmh.1923.s1-3.233 |
| Blanc (1930) | Greece | inoculation | 21 | *Freq. table.* Median: 7.0 (Range: 5.0–10.0) | — |
| Simmons (1931) | Philippines | mosquito bite | 67 | *Freq. table (interval-censored).* Range: 1.0–13.0 | — |
| Snijders (1931) | — | — | 3 | *Freq. table (interval-censored).* Range: 3.0–6.0 | — |
| CDC (1982) | — | — | 5 | *Freq. table (interval-censored).* Range: 0.1–11.0 | — |
| Anderson (2011) | — | — | 7 | *Freq. table (interval-censored).* Range: 3.0–14.0 | — |
| ***YFV*** | | | | | |
| Reed (1902) | Cuba | mosquito bite | 10 | *Freq. table.* Median: 3.6 (Range: 2.9–5.7) | — |
| Reed (1902) | Cuba | mosquito bite | 8 | *Freq. table.* Median: 3.8 (Range: 3.0–5.9) | — |
| Reed (1902) | Cuba | inoculation | 8 | *Freq. table (interval-censored).* Range: 0.5–5.5 | — |
| ***Nipah*** | | | | | |
| Rahman (2012) | Bangladesh | Bangladesh | 4 | Median: 10.0 (Range: 9.0–12.0) | 10.1089/vbz.2011.0656 |
| Rahman (2012) | Bangladesh | Bangladesh | 6 | Median: 4.0 (Range: 2.0–7.0) | 10.1089/vbz.2011.0656 |
| Nikolay (2019) | Bangladesh | Bangladesh | 11 | Median: 9.0 (Range: 6.0–14.0) | 10.1056/NEJMoa1805376 |
| Pallivalappil (2020) | India | — | 22 | Median: 9.5 (Range: 4.0–14.0) | 10.4103/jgid.jgid_4_19 |
| Ching (2015) | Philippines | — | 15 | Median: 8.0 (Range: 3.0–20.0) | 10.3201/eid2102.141433 |
| Luby (2009) | Bangladesh | Bangladesh | 14 | Median: 9.0 (Range: 6.0–11.0) | 10.3201/eid1508.081237 |
| Chandni (2019) | India | — | 11 | Median: 10.0 (Range: 8.0–15.0) | 10.1093/cid/ciz789 |
| Hossain (2008) | Bangladesh | Bangladesh | 11 | Median: 9.0 (Range: 6.0–11.0) | 10.1086/529147 |
| Nikolay (2019) | Bangladesh | Bangladesh | 82 | Median: 9.0 (IQR: 8.0–11.0) | 10.1056/NEJMoa1805376 |
| Thomas (2019) | India | — | 18 | Mean: 9.3 (SD: 1.9) | 10.4103/ijcm.IJCM_198_19 |

continued on next page

continued from previous page

| Reference | Country | Subgroup | $n$ | Summary | DOI |
|---|---|---|---|---|---|
| Nikolay (2019) | Bangladesh | Bangladesh | 11 | *Freq. table.* Median: 9.0 (Range: 6.0–14.0) | 10.1056/NEJMoa1805376 |
| ***Mpox*** | | | | | |
| Angelo (2023) | Mixed | IIb | 78 | Median: 8.0 (Range: 2.0–40.0) | 10.1016/S1473-3099(22)00651-X |
| Catala (2022) | Spain | IIb | 77 | Median: 6.0 (IQR: 4.0–9.0) | 10.1111/bjd.21790 |
| Choudury (2022) | Germany | IIb | 179 | Median: 7.0 (IQR: 4.0–10.0) | 10.3238/arztebl.m2022.0340 |
| Gaspari (2022) | Italy | IIb | 30 | Median: 9.0 (Range: 4.0–15.0) | 10.1128/jcm.01365-22 |
| Kroger (2023) | Germany | IIb | 209 | Mean: 8.2 (SD: 4.7) | 10.1007/s15010-023-01997-x |
| Mailhe (2023) | France | IIb | 86 | Median: 6.0 (IQR: 3.0–8.0) | 10.1016/j.cmi.2022.08.012 |
| Muira (2023) | Netherlands | IIb | 18 | Mean: 8.1 (SD: 4.4) | 10.1093/infdis/jiad091 |
| Moschese (2023) | Italy | IIb | 16 | Median: 11.0 (IQR: 11.0–16.0) | 10.1016/j.jinf.2022.08.019 |
| Nunez (2023) | Mexico | IIb | 18 | Median: 8.0 (IQR: 4.0–9.0) | 10.1016/j.lana.2022.100392 |
| Tarin-Vicente (2022) | Spain | IIb | 181 | Median: 7.0 (IQR: 5.0–11.0) | 10.1016/S0140-6736(22)01436-2 |
| Thornhill (2022) | Mixed | IIb | 51 | Median: 7.0 (IQR: 4.0–11.0) | 10.1016/S0140-6736(22)02187-0 |
| Thornhill (2022a), doi: 10.1056/NEJMo... | Mixed | IIb | 23 | Median: 7.0 (Range: 3.0–20.0) | 10.1056/NEJMoa2207323 |
| Charniga (2022) | USA | IIb | 22 | *Freq. table (interval-censored).* Range: 0.1–29.0 | 10.1101/2022.06.22.22276713 |
| Cobos (2023) | Spain | IIb | 19 | *Freq. table.* Median: 8.0 (Range: 2.0–17.0) | 10.37201/req/112.2022 |
| Guzzetta (2022) | Italy | IIb | 33 | *Freq. table (interval-censored).* Range: 0.1–24.0 | 10.3201/eid2810.221126 |
| McFarland (2023) | Mixed | IIb | 121 | *Freq. table (interval-censored).* Range: 0.1–35.0 | 10.2807/1560-7917.ES.2023.28.27.2200806 |
| ***Smallpox*** | | | | | |
| Clinical Society (1892) | UK | — | 39 | *Freq. table.* Median: 12.0 (Range: 9.0–20.0) | — |
| Robertson (1914) | Australia | — | 26 | *Freq. table.* Median: 12.0 (Range: 6.0–16.0) | — |
| Friedmann (1920) | Mixed | — | 25 | *Freq. table.* Median: 13.0 (Range: 9.0–17.0) | — |
| Downie (1972) | Mixed | — | 41 | *Freq. table.* Median: 13.0 (Range: 8.0–17.0) | — |
| ***Measles*** | | | | | |
| Goodall (1931) | United Kingdom | — | 116 | *Freq. table.* Median: 12.0 (Range: 6.0–25.0) | — |
| Goodall (1931) | United Kingdom | — | 25 | *Freq. table.* Median: 12.0 (Range: 7.0–25.0) | — |
| Ehresmann (1995) | USA | — | 16 | *Freq. table.* Median: 14.5 (Range: 11.0–22.0) | 10.1093/infdis/171.3.679 |
| Bloch (1985) | USA | — | 8 | Median: 14.0 (Range: 12.0–18.0) | — |
| Fielding (2005) | Australia | — | 21 | Median: 12.0 (Range: 8.0–17.0) | — |
| Kobayashi (2020) | Japan | — | 30 | Median: 16.0 (Range: 11.0–21.0) | 10.1016/j.vaccine.2020.05.067 |
| Ehresmann (1995) | USA | — | 16 | Median: 14.0 (Range: 11.0–22.0) | 10.1093/infdis/171.3.679 |
| Papania (1999) | USA | — | 34 | Median: 14.0 (Range: 10.0–21.0) | 10.1542/peds.104.5.e59 |
| Komabayashi (2018) | Japan | — | 22 | Mean: 13.8 (SD: 2.7) | 10.7883/yoken.JJID.2018.083 |
| Komabayashi (2018) | Japan | — | 38 | Mean: 14.2 (SD: 2.9) | 10.7883/yoken.JJID.2018.083 |
| Sheline (1987) | USA | — | 9 | Median: 14.0 (Range: 13.0–17.0) | — |
| Panum (1847) | Faroe Islands | — | 38 | *Freq. table (interval-censored).* Range: 12.0–15.0 | — |
| ***Cholera*** | | | | | |
| Morris (1995) | USA | O139 | 7 | Median: 0.9 (Range: 0.8–2.0) | 10.1093/infdis/171.4.903 |
| Morris (1995) | USA | O139 | 11 | Median: 1.3 (Range: 0.8–2.7) | 10.1093/infdis/171.4.903 |
| Snow (1855) | United Kingdom | O1 Classical | 25 | *Freq. table (interval-censored).* Range: 0.1–11.0 | — |
| Cash (1974) | USA | O1 Classical | 40 | *Freq. table (interval-censored).* Range: 0.8–6.7 | 10.1093/infdis/129.1.45 |
| Cash (1974) | USA | O1 Classical | 22 | *Freq. table (interval-censored).* Range: 0.4–3.8 | 10.1093/infdis/129.1.45 |
| Eberhart-Phillips (1996) | — | O1 El Tor | 66 | *Freq. table (interval-censored).* Range: 0.1–7.0 | 10.1017/S0950268800058891 |
| Goh (1984) | Singapore | O1 El Tor Ogawa | 18 | *Freq. table.* Median: 1.6 (Range: 0.2–8.5) | 10.1093/ije/13.2.210 |

continued on next page

continued from previous page

| Reference | Country | Subgroup | $n$ | Summary | DOI |
|---|---|---|---|---|---|
| Levine (1979) | USA | O1 Classical | 9 | *Freq. table (interval-censored).* Range: 0.6–4.6 | 10.1016/0035-9203(79)90119-6 |
| Levine (1979) | USA | O1 Classical | 11 | *Freq. table (interval-censored).* Range: 0.6–4.3 | 10.1016/0035-9203(79)90119-6 |
| Schiraldi (1974) | Italy | O1 El Tor Ogawa | 53 | *Freq. table.* Median: 2.0 (Range: 0.0–6.0) | — |
| Sutton (1974) | USA | O1 El Tor | 23 | *Freq. table (interval-censored).* Range: 0.0–6.2 | 10.1017/S0022172400023688 |
| Cohen (1999) | USA | O139 | 22 | *Freq. table (interval-censored).* Range: 0.9–2.4 | 10.1128/IAI.67.12.6346-6349.1999 |
| Coster (1995) | USA | O139 | 5 | *Freq. table.* Median: 0.9 (Range: 0.8–1.9) | 10.1016/S0140-6736(95)90698-3 |
| Hornick (1971) | USA | O1 Classical | 37 | *Freq. table.* Median: 2.0 (Range: 1.5–2.0) | — |
| Sack (1998) | USA | O1 El Tor | 34 | Mean: 1.2 (SD: 0.4) | 10.1128/iai.66.5.1968-1972.1998 |
| Taylor (1993) | USA | O1 El Tor Ogawa | 3 | *Freq. table (interval-censored).* Range: 0.3–3.0 | 10.1093/infdis/167.6.1330 |
| ***Typhoid*** | | | | | |
| Anita (2012) | Malaysia | Other | 12 | *Freq. table.* Median: 18.0 (Range: 3.0–21.0) | — |
| Caraway (1961) | USA | Catered meal | 33 | *Freq. table.* Median: 20.0 (Range: 7.0–34.0) | — |
| Coté (1995) | USA | Other | 24 | *Freq. table.* Median: 14.0 (Range: 1.0–28.0) | — |
| Kobayashi (2016) | Japan | Other | 7 | *Freq. table.* Median: 17.0 (Range: 10.0–27.0) | — |
| Michel (2005) | Ivory Coast | Other | 24 | *Freq. table.* Median: 23.0 (Range: 9.0–34.0) | 10.1007/s10654-005-7454-6 |
| Naylor GR (1983) | Mixed | Experimental | 109 | *Freq. table.* Median: 9.0 (Range: 3.0–41.0) | — |
| Pradier (2000) | France | Catered meal | 28 | *Freq. table.* Median: 17.0 (Range: 9.0–33.0) | — |
| Ramsey (1926) | USA | Catered meal | 35 | *Freq. table.* Median: 13.0 (Range: 2.0–41.0) | — |
| Sawyer (1914) | USA | Catered meal | 84 | *Freq. table.* Median: 7.0 (Range: 3.0–29.0) | — |
| Waddington (2014) | Mixed | Experimental | 20 | Median: 9.0 (IQR: 6.5–13.0) | 10.1093/cid/ciu078 |
| Waddington (2014) | Mixed | Experimental | 20 | Median: 8.0 (IQR: 6.0–9.0) | 10.1093/cid/ciu078 |
| Moore (1950) | UK | Other | 41 | *Freq. table.* Median: 9.0 (Range: 6.0–21.0) | — |
| Cumming (1917) | USA | Other | 23 | *Freq. table.* Median: 7.0 (Range: 5.0–12.0) | — |
| Brooks (2012) | USA | Other | 12 | Median: 6.0 (IQR: 4.0–11.0) | 10.1089/fpd.2011.0992 |
| Naylor (1983) | Mixed | Experimental | 109 | *Freq. table.* Median: 9.0 (Range: 3.0–41.0) | 10.1016/S0140-6736(83)91395-8 |
| Caraway (1961) | USA | Other | 33 | *Freq. table.* Median: 20.0 (Range: 7.0–34.0) | — |
| Couper (1956) | UK | — | 23 | *Freq. table.* Median: 8.0 (Range: 6.0–20.0) | 10.1016/S0140-6736(56)90818-2 |
| Ramsey (1926) | USA | Other | 35 | *Freq. table.* Median: 13.0 (Range: 2.0–41.0) | — |
| Hill (cited in Miner JR, 1922), doi: ... | USA | Water | 21 | *Freq. table.* Median: 15.0 (Range: 5.0–29.0) | 10.1093/infdis/31.3.296 |
| Lumsden (cited in Miner JR, 1922), do... | USA | Water | 13 | *Freq. table.* Median: 16.0 (Range: 7.0–38.0) | 10.1093/infdis/31.3.296 |
| Ferguson (cited in Miner JR, 1922), d... | USA | Water | 181 | *Freq. table (interval-censored).* Range: 7.0–40.0 | 10.1093/infdis/31.3.296 |
| Egoz (1988) | Israel | Water | 76 | Median: 22.0 (Range: 12.0–40.0) | — |

##### C.1. Ebola (EVD)

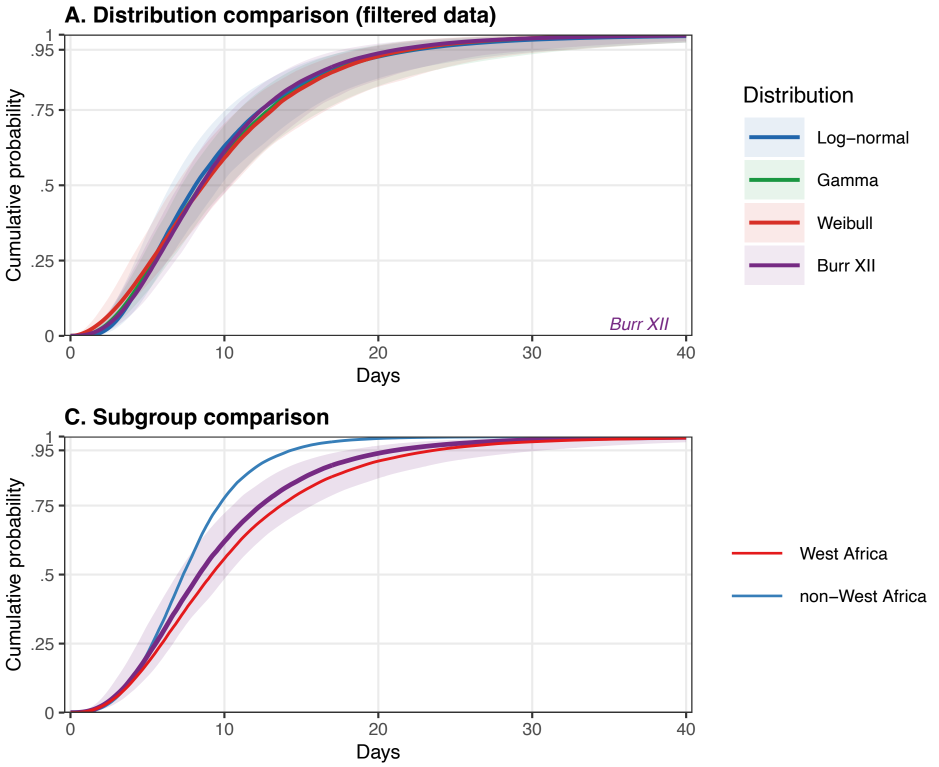

**Supplementary Figure S5. Incubation period model fits for Ebola (EVD):** (A) Posterior predictive cumulative distribution functions (CDFs) for all converged parametric distributions fitted to the filtered dataset. Ribbons indicate 95% credible intervals; the best-fitting distribution is annotated. (B) Comparison of CDFs fitted to all data vs. the filtered dataset for the best-fitting distribution; shown only when filtering removed at least one dataset. (C) Subgroup CDFs for the best-fitting distribution; the overall estimate (shaded ribbon) is shown alongside subgroup-specific estimates; shown only when subgroup analyses were performed.

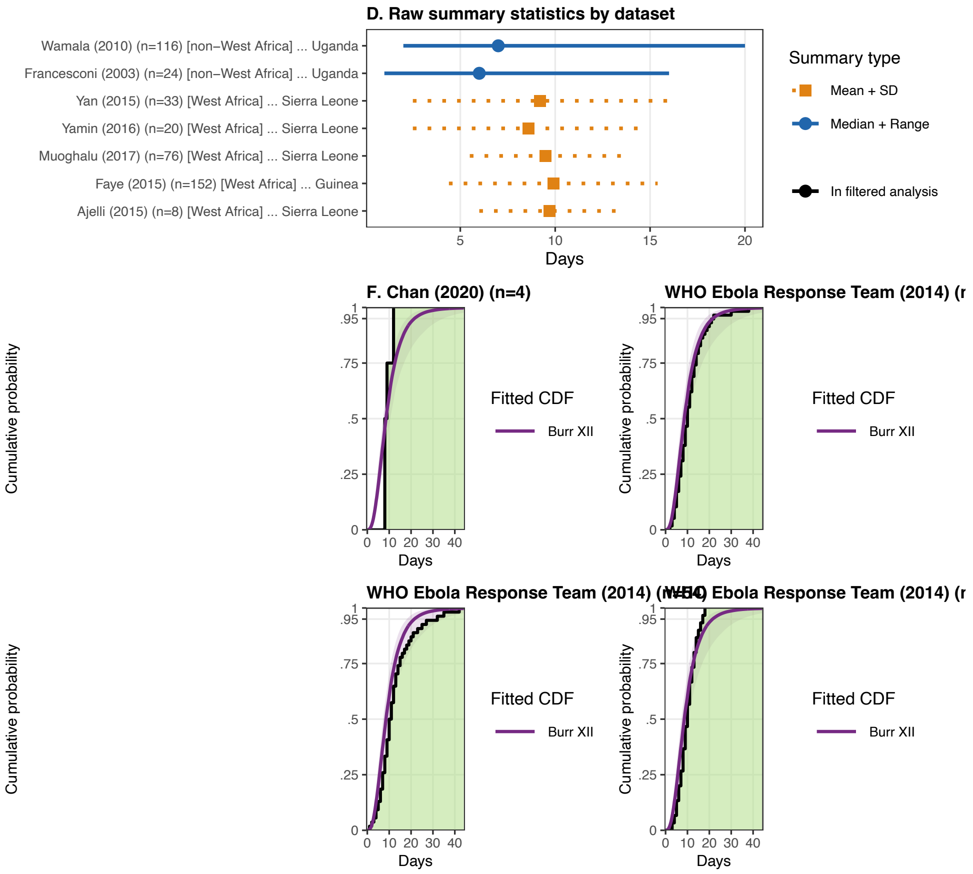

**Supplementary Figure S6. Raw incubation period data for Ebola (EVD):** (D) Raw summary statistics reported in source datasets. Points indicate the central estimate (circle: median; square: mean) and lines the reported uncertainty interval (solid: range; dashed: IQR; dotted: SD). Faded entries were excluded by the data-quality filter. (F) Empirical CDFs for datasets reporting frequency tables or interval-censored observations, overlaid with the posterior predictive CDF for the best-fitting distribution (coloured ribbon and line). For interval-censored data, the solid step line is the conservative ECDF at interval upper bounds; the dashed step line is the ECDF at interval lower bounds; the shaded band represents the uncertainty region.

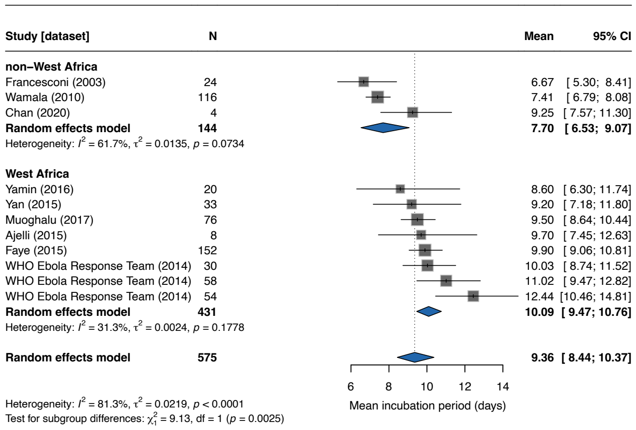

**Supplementary Figure S7. Forest plot of mean incubation period for Ebola (EVD):** Squares represent study-specific estimates with 95% confidence intervals (CIs); square size is proportional to study weight. The diamond shows the overall pooled estimate from a random-effects model with log-transformed mean (back-transformed to days). Studies are stratified by location; subgroup pooled estimates are shown as separate diamonds. Heterogeneity is quantified by $I^2$ and $\tau^2$.

##### C.2. Marburg (MVD)

Only two estimates of the incubation period, both in summary type format, were found in the literature. However, published estimates of longer incubation periods point to inferred incubation period potentially underestimating the 95th percentile; outliers reported in Schneider et al. [24] indicate that the 95th percentile of the incubation period distribution might be similar to that of Ebola Virus Disease.

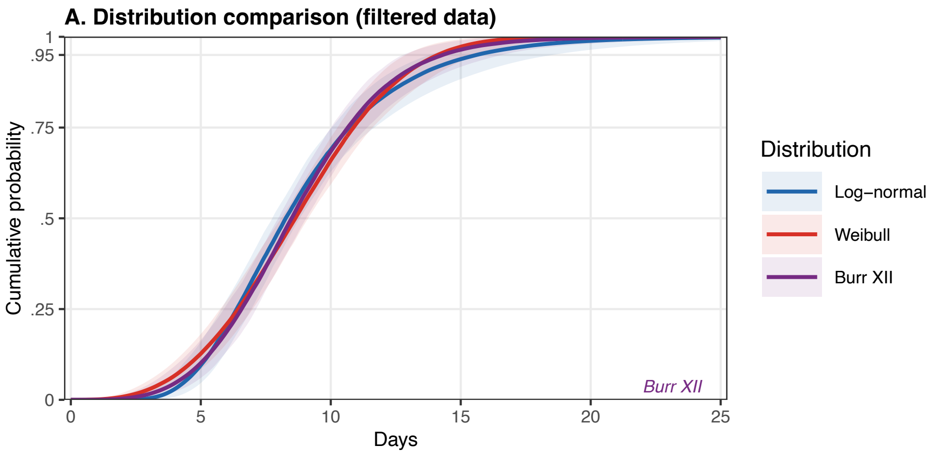

**Supplementary Figure S8. Incubation period model fits for Marburg (MVD):** (A) Posterior predictive cumulative distribution functions (CDFs) for all converged parametric distributions fitted to the filtered dataset. Ribbons indicate 95% credible intervals; the best-fitting distribution is annotated. (B) Comparison of CDFs fitted to all data vs. the filtered dataset for the best-fitting distribution; shown only when filtering removed at least one dataset. (C) Subgroup CDFs for the best-fitting distribution; the overall estimate (shaded ribbon) is shown alongside subgroup-specific estimates; shown only when subgroup analyses were performed.

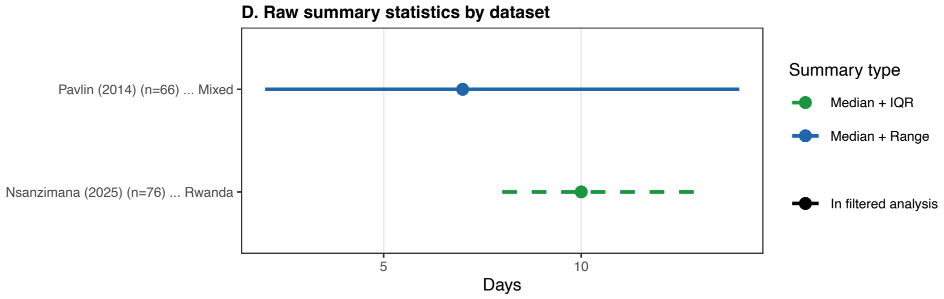

**Supplementary Figure S9. Raw incubation period data for Marburg (MVD):** (D) Raw summary statistics reported in source datasets. Points indicate the central estimate (circle: median; square: mean) and lines the reported uncertainty interval (solid: range; dashed: IQR; dotted: SD). Faded entries were excluded by the data-quality filter. (F) Empirical CDFs for datasets reporting frequency tables or interval-censored observations, overlaid with the posterior predictive CDF for the best-fitting distribution (coloured ribbon and line). For interval-censored data, the solid step line is the conservative ECDF at interval upper bounds; the dashed step line is the ECDF at interval lower bounds; the shaded band represents the uncertainty region.

##### C.3. Lassa Fever

The incubation period distribution estimate is based on only one dataset of individual level interval censored data. It is also important to note that this is from a nosocomial dataset from 1972 [53] and that this is not the dominant transmission route for Lassa fever. More information on Lassa fever is contained in Doohan et al. [18].

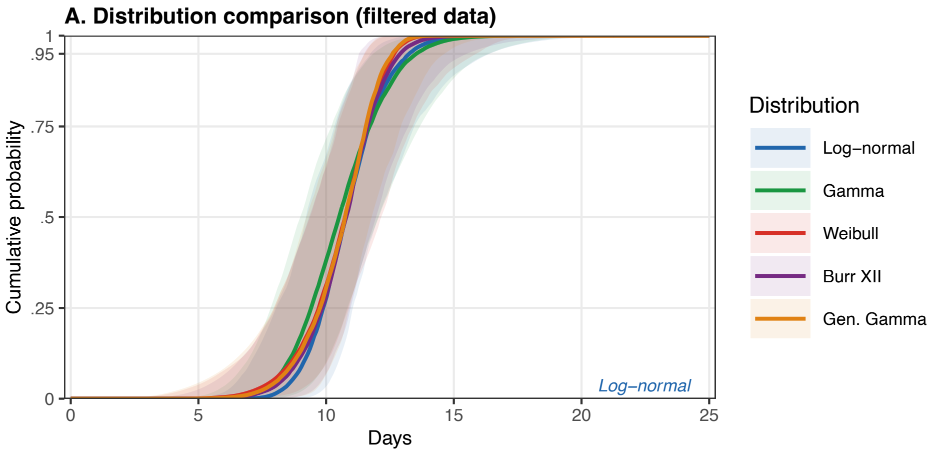

**Supplementary Figure S10. Incubation period model fits for Lassa:** (A) Posterior predictive cumulative distribution functions (CDFs) for all converged parametric distributions fitted to the filtered dataset. Ribbons indicate 95% credible intervals; the best-fitting distribution is annotated. (B) Comparison of CDFs fitted to all data vs. the filtered dataset for the best-fitting distribution; shown only when filtering removed at least one dataset. (C) Subgroup CDFs for the best-fitting distribution; the overall estimate (shaded ribbon) is shown alongside subgroup-specific estimates; shown only when subgroup analyses were performed.

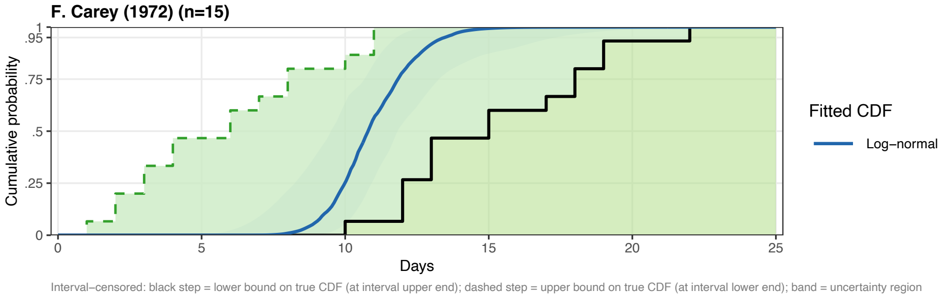

**Supplementary Figure S11. Raw incubation period data for Lassa:** (D) Raw summary statistics reported in source datasets. Points indicate the central estimate (circle: median; square: mean) and lines the reported uncertainty interval (solid: range; dashed: IQR; dotted: SD). Faded entries were excluded by the data-quality filter. (F) Empirical CDFs for datasets reporting frequency tables or interval-censored observations, overlaid with the posterior predictive CDF for the best-fitting distribution (coloured ribbon and line). For interval-censored data, the solid step line is the conservative ECDF at interval upper bounds; the dashed step line is the ECDF at interval lower bounds; the shaded band represents the uncertainty region.

##### C.4. CCHF

For CCHF we consider a subgroup analysis of nosocomial transmission vs tick-bites.

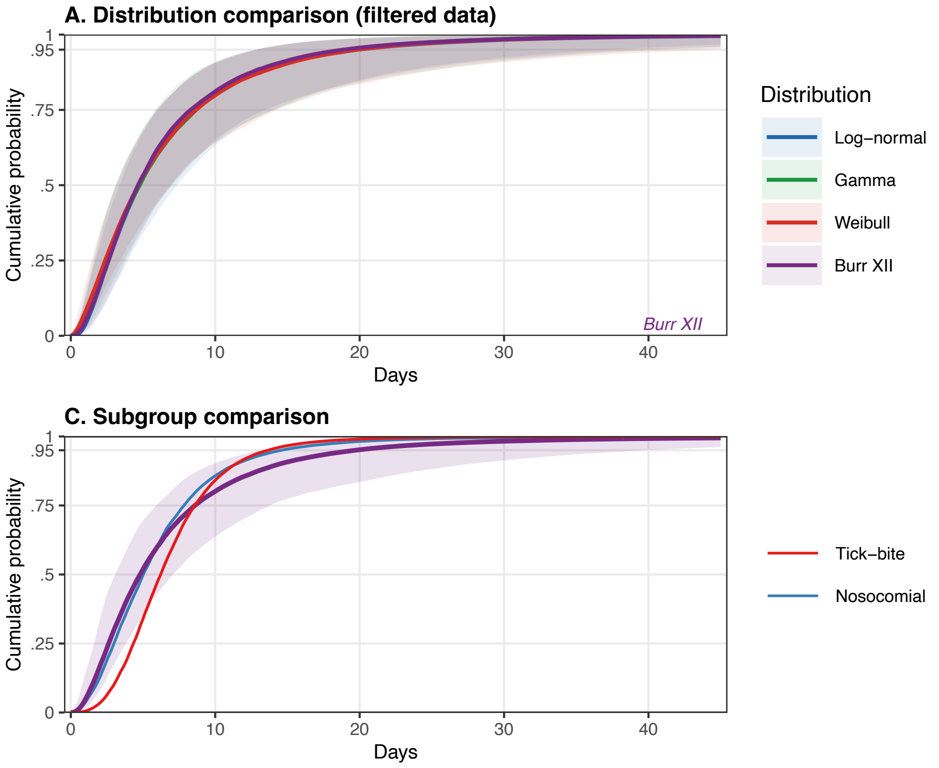

**Supplementary Figure S12. Incubation period model fits for CCHF:** (A) Posterior predictive cumulative distribution functions (CDFs) for all converged parametric distributions fitted to the filtered dataset. Ribbons indicate 95% credible intervals; the best-fitting distribution is annotated. (B) Comparison of CDFs fitted to all data vs. the filtered dataset for the best-fitting distribution; shown only when filtering removed at least one dataset. (C) Subgroup CDFs for the best-fitting distribution; the overall estimate (shaded ribbon) is shown alongside subgroup-specific estimates; shown only when subgroup analyses were performed.

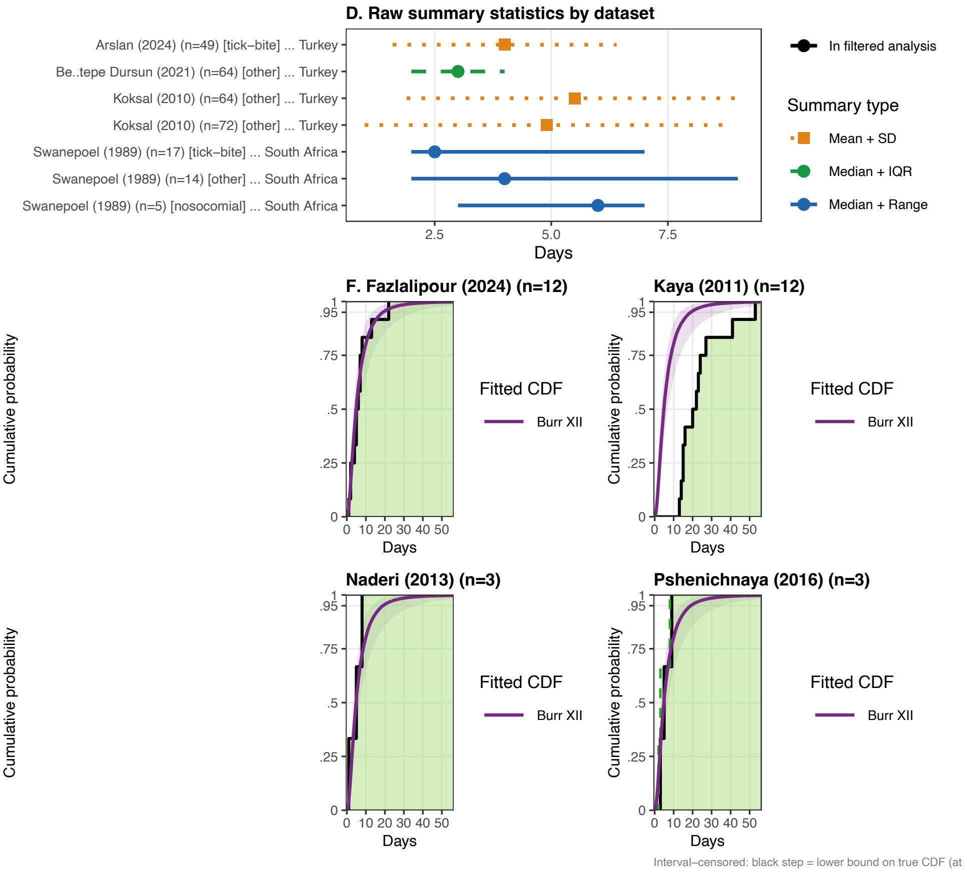

**Supplementary Figure S13. Raw incubation period data for CCHF:** (D) Raw summary statistics reported in source datasets. Points indicate the central estimate (circle: median; square: mean) and lines the reported uncertainty interval (solid: range; dashed: IQR; dotted: SD). Faded entries were excluded by the data-quality filter. (F) Empirical CDFs for datasets reporting frequency tables or interval-censored observations, overlaid with the posterior predictive CDF for the best-fitting distribution (coloured ribbon and line). For interval-censored data, the solid step line is the conservative ECDF at interval upper bounds; the dashed step line is the ECDF at interval lower bounds; the shaded band represents the uncertainty region.

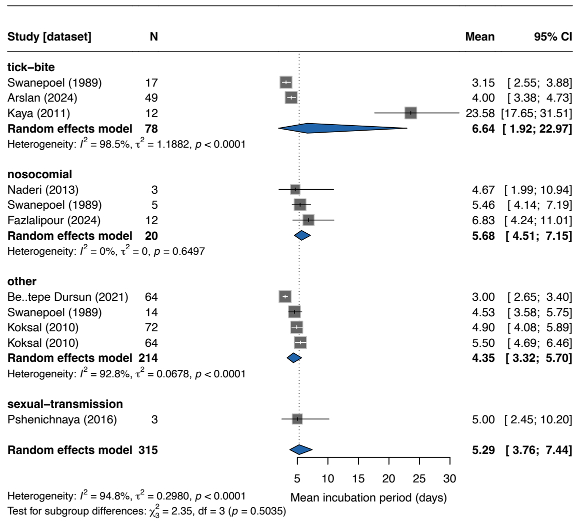

**Supplementary Figure S14. Forest plot of mean incubation period for CCHF:** Squares represent study-specific estimates with 95% confidence intervals (CIs); square size is proportional to study weight. The diamond shows the overall pooled estimate from a random-effects model with log-transformed mean (back-transformed to days). Studies are stratified by transmission route; subgroup pooled estimates are shown as separate diamonds. Heterogeneity is quantified by $I^2$ and $\tau^2$.

##### C.5. COVID-19

COVID-19 is unique in the amount of data available. Subgroup analysis was performed for SARS-CoV-2 variants but other subgroup analyses are possible (e.g., by country for the Wildtype variant).

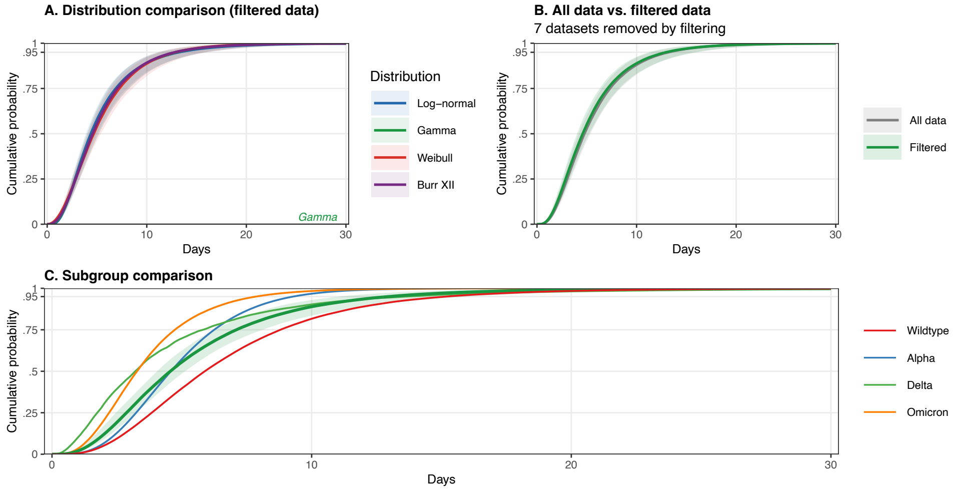

**Supplementary Figure S15. Incubation period model fits for COVID-19:** (A) Posterior predictive cumulative distribution functions (CDFs) for all converged parametric distributions fitted to the filtered dataset. Ribbons indicate 95% credible intervals; the best-fitting distribution is annotated. (B) Comparison of CDFs fitted to all data vs. the filtered dataset for the best-fitting distribution; shown only when filtering removed at least one dataset. (C) Subgroup CDFs for the best-fitting distribution; the overall estimate (shaded ribbon) is shown alongside subgroup-specific estimates; shown only when subgroup analyses were performed.

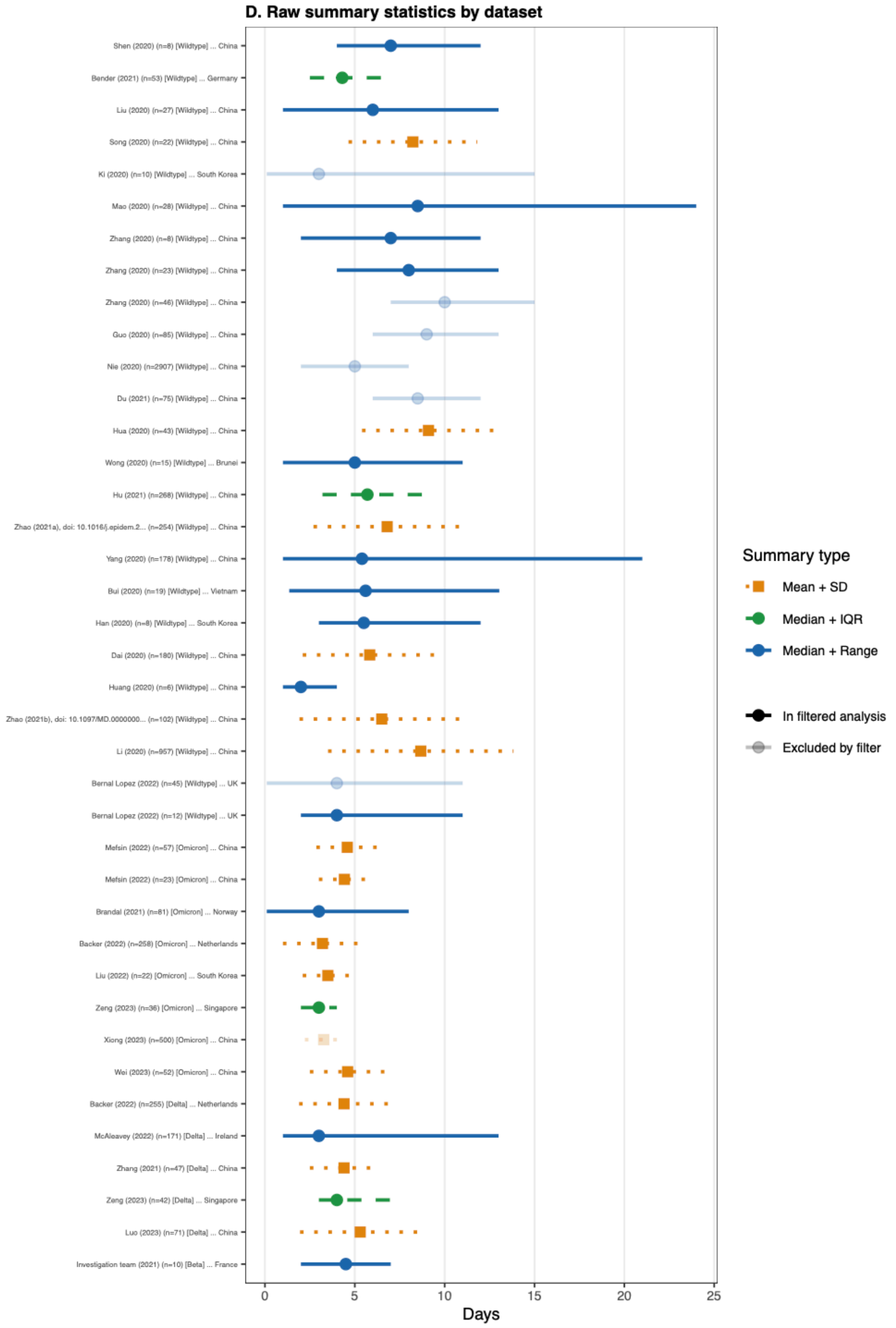

**Supplementary Figure S16. Raw incubation period data for COVID-19:** (D) Raw summary statistics reported in source datasets. Points indicate the central estimate (circle: median; square: mean) and lines the reported uncertainty interval (solid: range; dashed: IQR; dotted: SD). Faded entries were excluded by the data-quality filter.

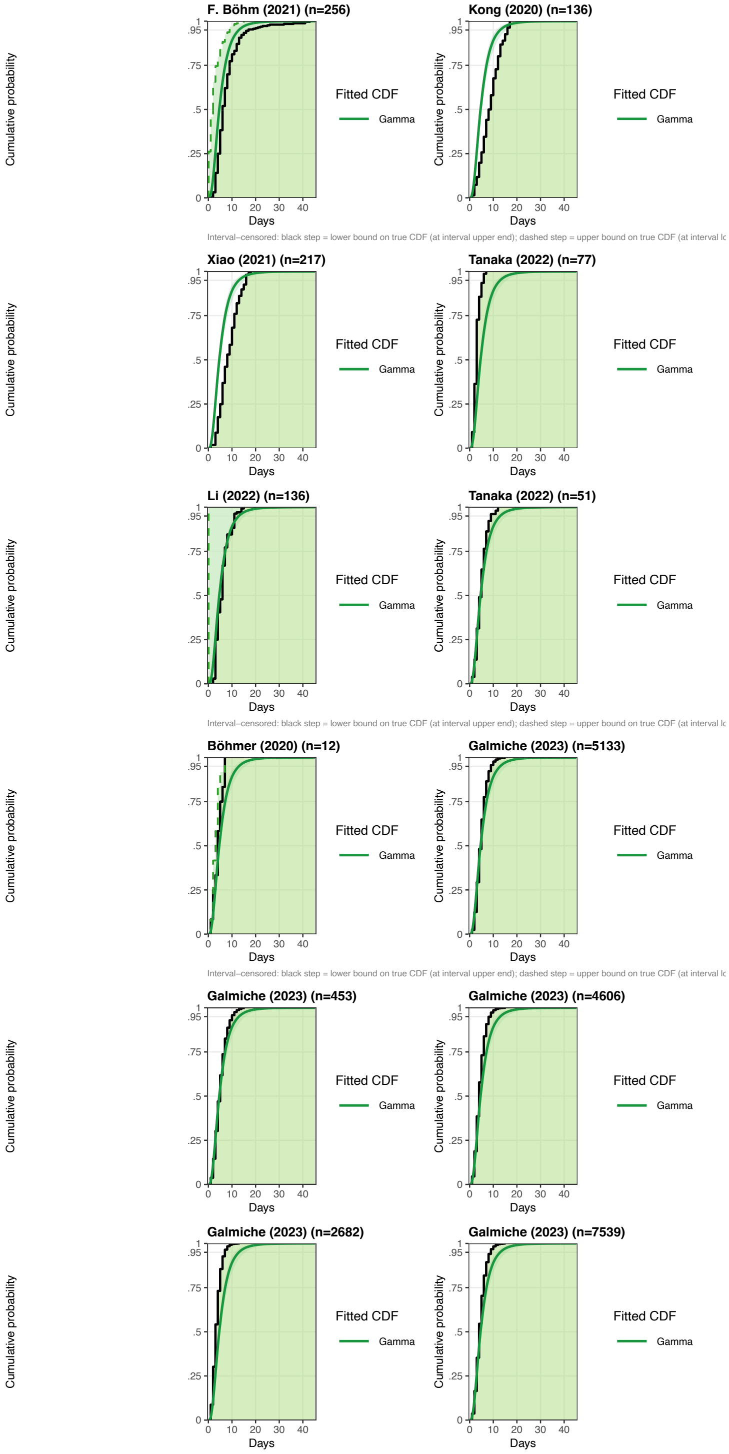

**Supplementary Figure S17. Raw incubation period data for COVID-19:** (F) Empirical CDFs for datasets reporting frequency tables or interval-censored observations, overlaid with the posterior predictive CDF for the best-fitting distribution (coloured ribbon and line). For interval-censored data, the solid step line is the conservative ECDF at interval upper bounds; the dashed step line is the ECDF at interval lower bounds; the shaded band represents the uncertainty region.

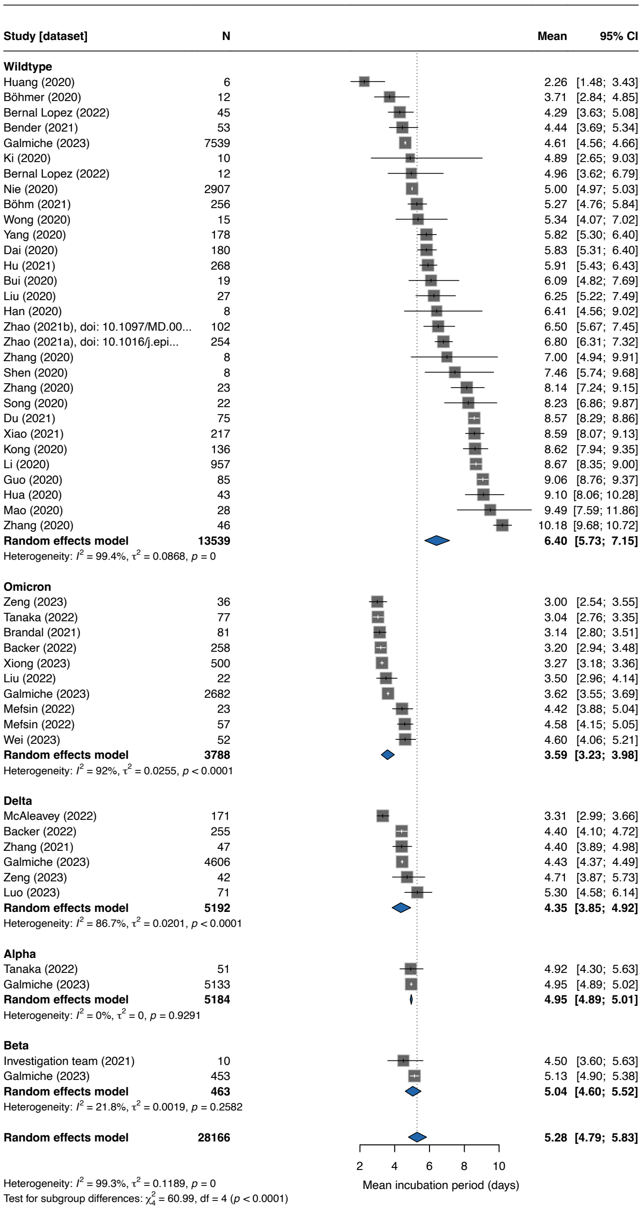

**Supplementary Figure S18. Forest plot of mean incubation period for COVID-19:** Squares represent study-specific estimates with 95% confidence intervals (CIs); square size is proportional to study weight. The diamond shows the overall pooled estimate from a random-effects model with log-transformed mean (back-transformed to days). Studies are stratified by SARS-CoV-2 variant; subgroup pooled estimates are shown as separate diamonds. Heterogeneity is quantified by $I^2$ and $\tau^2$.

##### C.6. SARS

We performed subgroup analysis by country for SARS. There was a single outbreak of SARS-CoV-1, but differences incubation period estimates arose due to different underlying patient populations and healthcare systems [19].

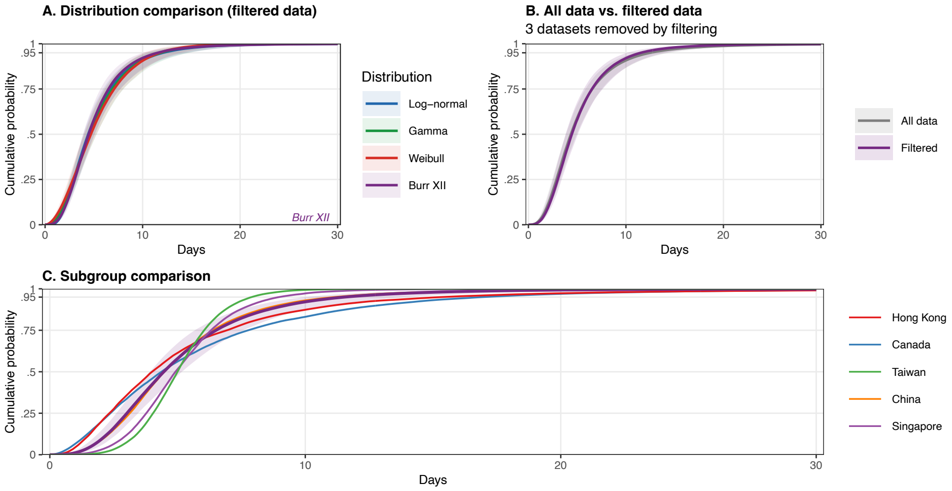

**Supplementary Figure S19. Incubation period model fits for SARS:** (A) Posterior predictive cumulative distribution functions (CDFs) for all converged parametric distributions fitted to the filtered dataset. Ribbons indicate 95% credible intervals; the best-fitting distribution is annotated. (B) Comparison of CDFs fitted to all data vs. the filtered dataset for the best-fitting distribution; shown only when filtering removed at least one dataset. (C) Subgroup CDFs for the best-fitting distribution; the overall estimate (shaded ribbon) is shown alongside subgroup-specific estimates; shown only when subgroup analyses were performed.

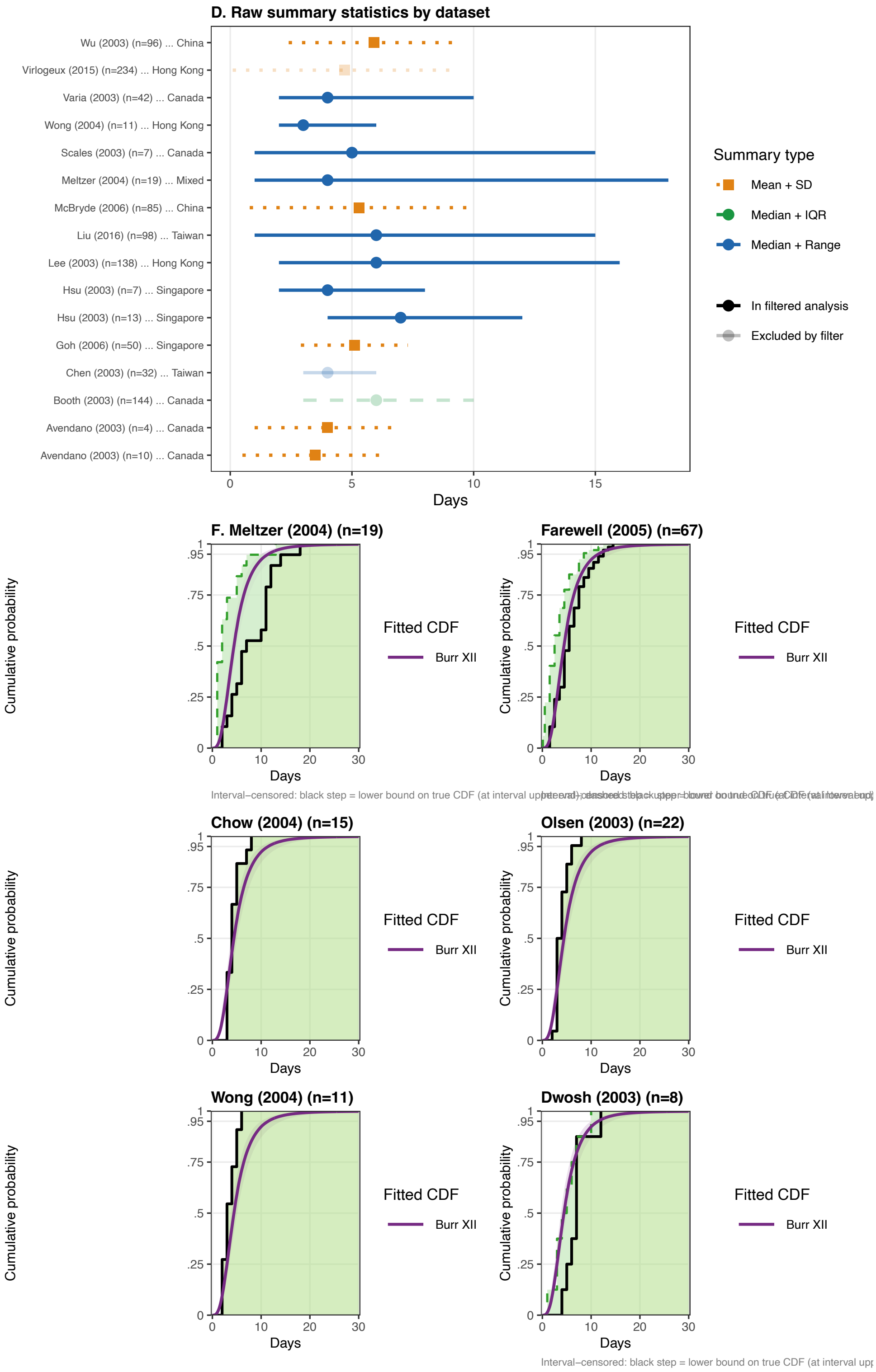

**Supplementary Figure S20. Raw incubation period data for SARS:** (D) Raw summary statistics reported in source datasets. Points indicate the central estimate (circle: median; square: mean) and lines the reported uncertainty interval (solid: range; dashed: IQR; dotted: SD). Faded entries were excluded by the data-quality filter. (F) Empirical CDFs for datasets reporting frequency tables or interval-censored observations, overlaid with the posterior predictive CDF for the best-fitting distribution (coloured ribbon and line). For interval-censored data, the solid step line is the conservative ECDF at interval upper bounds; the dashed step line is the ECDF at interval lower bounds; the shaded band represents the uncertainty region.

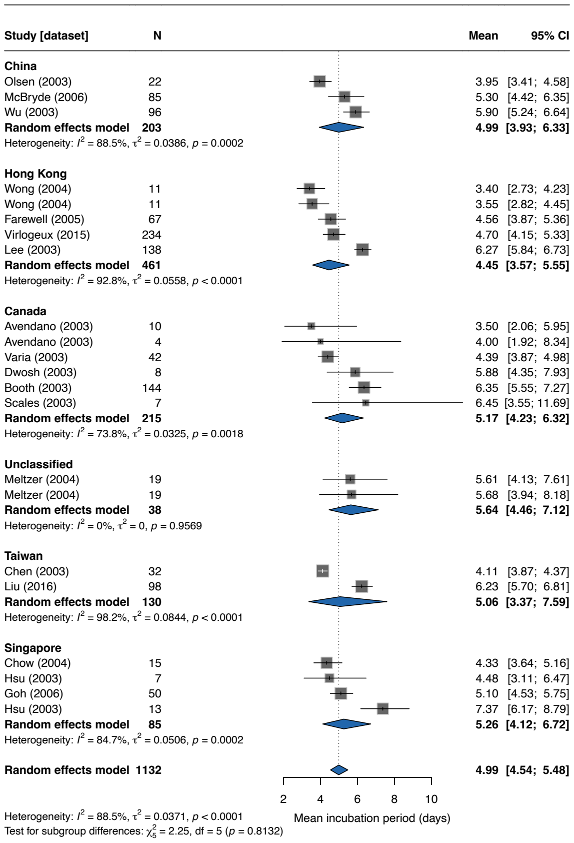

**Supplementary Figure S21. Forest plot of mean incubation period for SARS:** Squares represent study-specific estimates with 95% confidence intervals (CIs); square size is proportional to study weight. The diamond shows the overall pooled estimate from a random-effects model with log-transformed mean (back-transformed to days). Studies are stratified by country; subgroup pooled estimates are shown as separate diamonds. Heterogeneity is quantified by $I^2$ and $\tau^2$.

#### C.7. MERS

In the subgroup analysis we compare all data with the estimate of the incubation period distribution restricted to data from only Saudi Arabia.

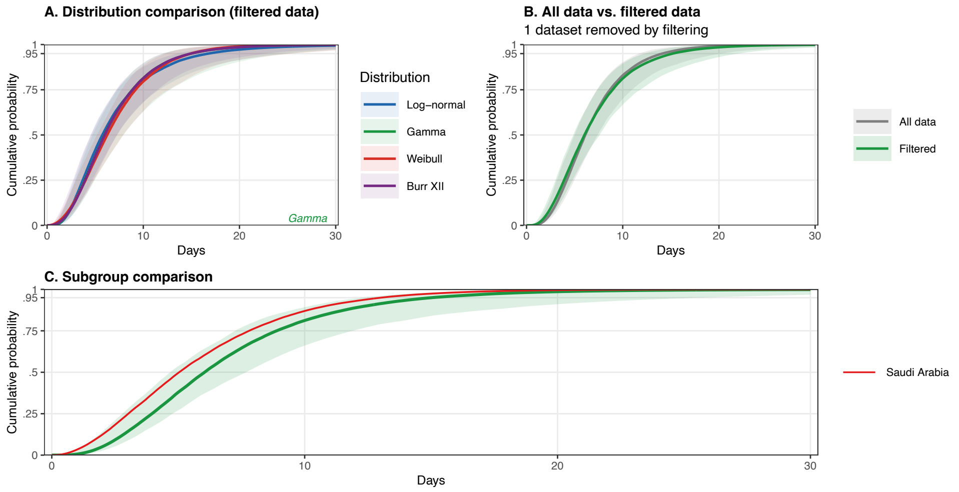

**Supplementary Figure S22. Incubation period model fits for MERS:** (A) Posterior predictive cumulative distribution functions (CDFs) for all converged parametric distributions fitted to the filtered dataset. Ribbons indicate 95% credible intervals; the best-fitting distribution is annotated. (B) Comparison of CDFs fitted to all data vs. the filtered dataset for the best-fitting distribution; shown only when filtering removed at least one dataset. (C) Subgroup CDFs for the best-fitting distribution; the overall estimate (shaded ribbon) is shown alongside subgroup-specific estimates; shown only when subgroup analyses were performed.

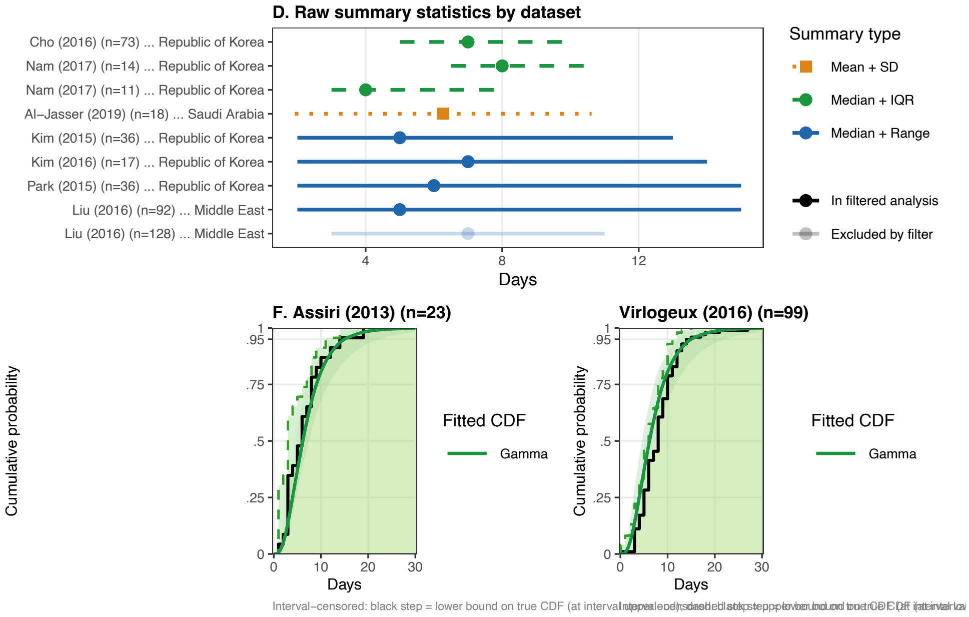

**Supplementary Figure S23. Raw incubation period data for MERS:** (D) Raw summary statistics reported in source datasets. Points indicate the central estimate (circle: median; square: mean) and lines the reported uncertainty interval (solid: range; dashed: IQR; dotted: SD). Faded entries were excluded by the data-quality filter. (F) Empirical CDFs for datasets reporting frequency tables or interval-censored observations, overlaid with the posterior predictive CDF for the best-fitting distribution (coloured ribbon and line). For interval-censored data, the solid step line is the conservative ECDF at interval upper bounds; the dashed step line is the ECDF at interval lower bounds; the shaded band represents the uncertainty region.

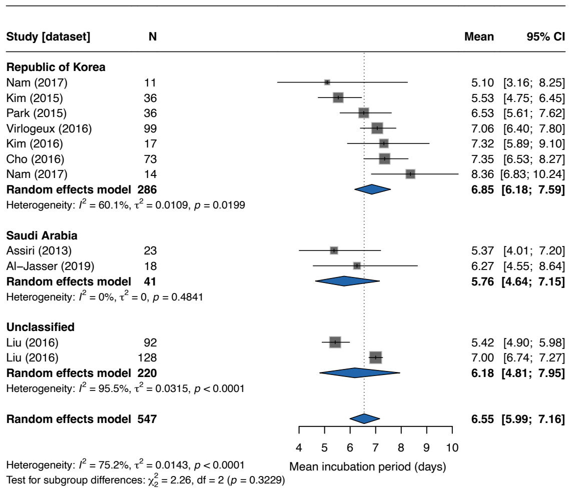

**Supplementary Figure S24. Forest plot of mean incubation period for MERS:** Squares represent study-specific estimates with 95% confidence intervals (CIs); square size is proportional to study weight. The diamond shows the overall pooled estimate from a random-effects model with log-transformed mean (back-transformed to days). Studies are stratified by country; subgroup pooled estimates are shown as separate diamonds. Heterogeneity is quantified by $I^2$ and $\tau^2$.

#### C.8. Influenza

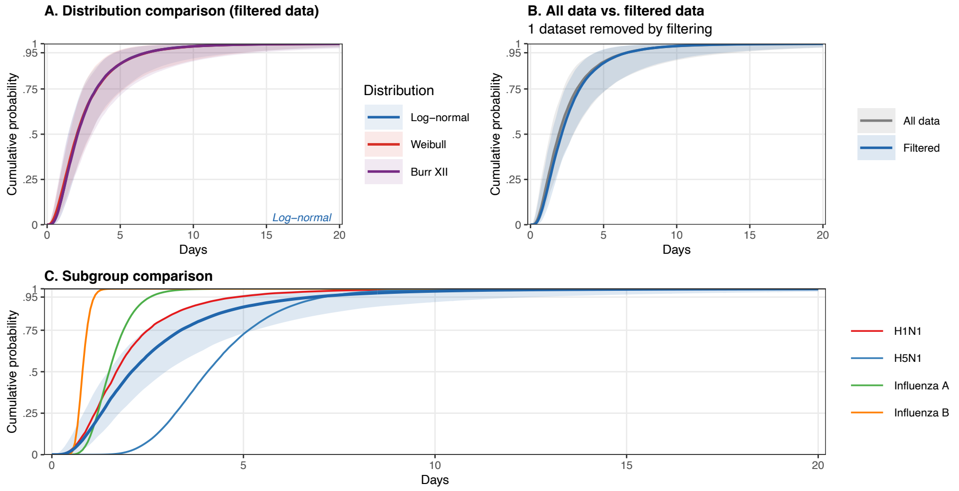

**Supplementary Figure S25. Incubation period model fits for Influenza:** (A) Posterior predictive cumulative distribution functions (CDFs) for all converged parametric distributions fitted to the filtered dataset. Ribbons indicate 95% credible intervals; the best-fitting distribution is annotated. (B) Comparison of CDFs fitted to all data vs. the filtered dataset for the best-fitting distribution; shown only when filtering removed at least one dataset. (C) Subgroup CDFs for the best-fitting distribution; the overall estimate (shaded ribbon) is shown alongside subgroup-specific estimates; shown only when subgroup analyses were performed.

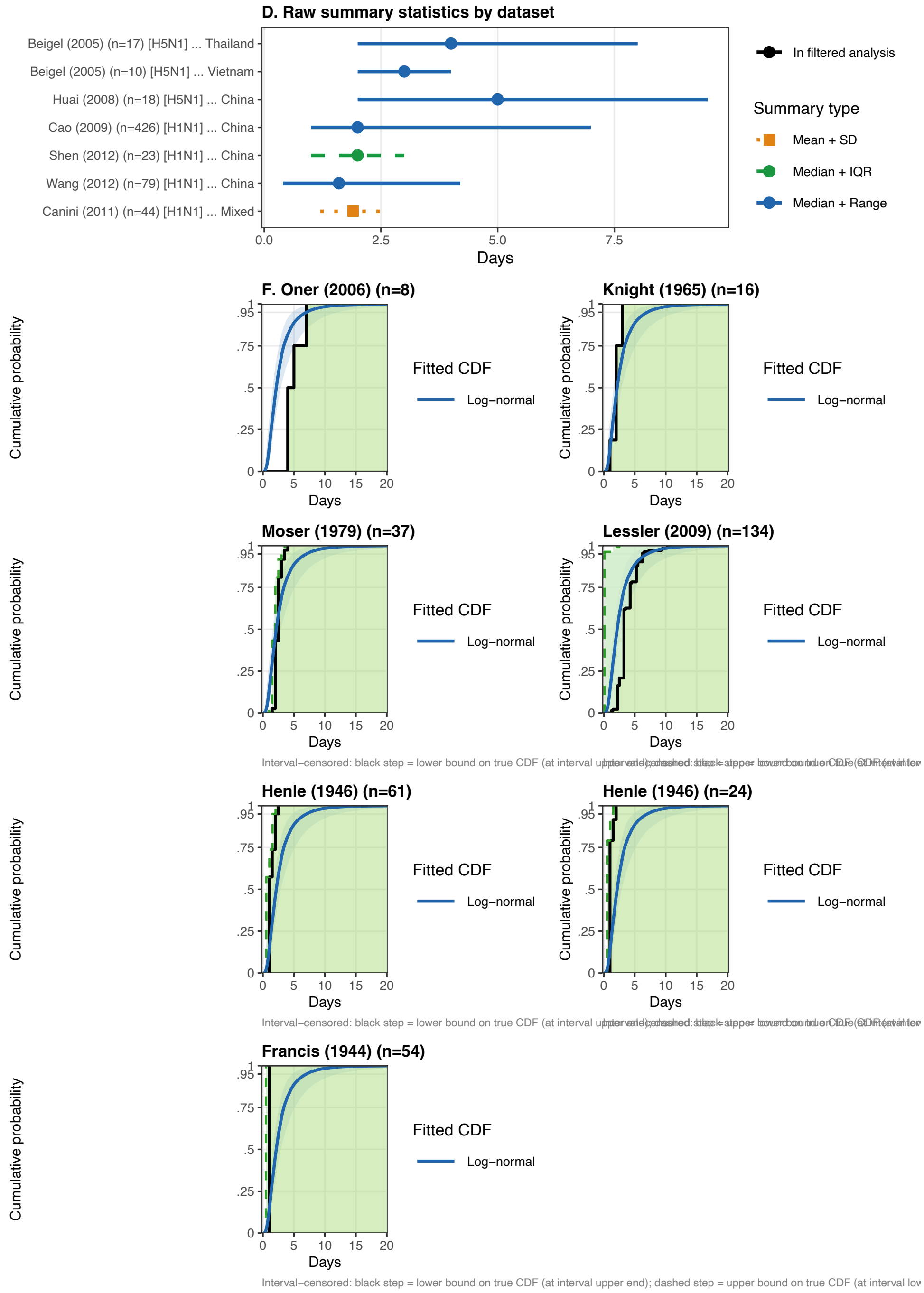

**Supplementary Figure S26. Raw incubation period data for Influenza:** (D) Raw summary statistics reported in source datasets. Points indicate the central estimate (circle: median; square: mean) and lines the reported uncertainty interval (solid: range; dashed: IQR; dotted: SD). Faded entries were excluded by the data-quality filter. (F) Empirical CDFs for datasets reporting frequency tables or interval-censored observations, overlaid with the posterior predictive CDF for the best-fitting distribution (coloured ribbon and line). For interval-censored data, the solid step line is the conservative ECDF at interval upper bounds; the dashed step line is the ECDF at interval lower bounds; the shaded band represents the uncertainty region.

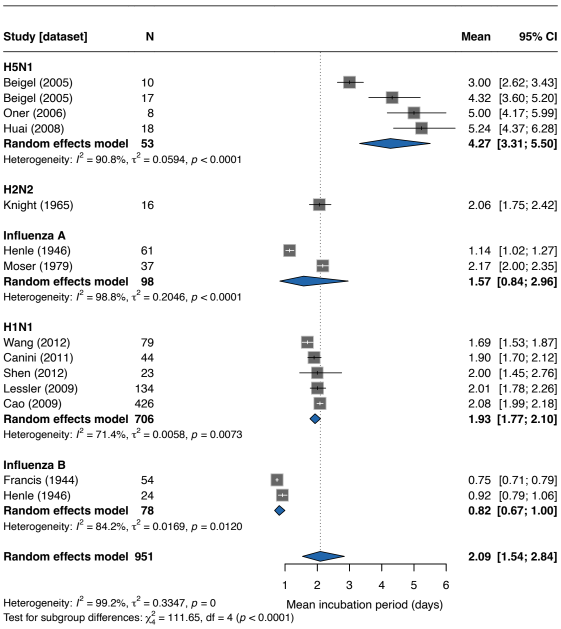

**Supplementary Figure S27. Forest plot of mean incubation period for Influenza:** Squares represent study-specific estimates with 95% confidence intervals (CIs); square size is proportional to study weight. The diamond shows the overall pooled estimate from a random-effects model with log-transformed mean (back-transformed to days). Studies are stratified by subtype/genus; subgroup pooled estimates are shown as separate diamonds. Heterogeneity is quantified by $I^2$ and $\tau^2$.

**C.9. Dengue**

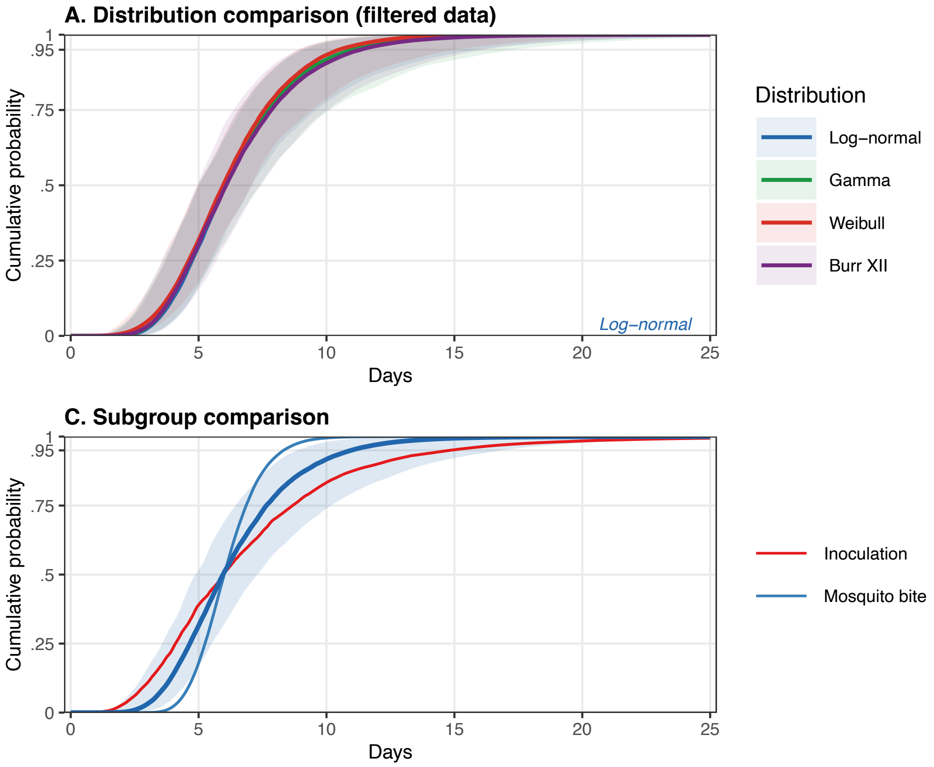

**Supplementary Figure S28. Incubation period model fits for Dengue:** (A) Posterior predictive cumulative distribution functions (CDFs) for all converged parametric distributions fitted to the filtered dataset. Ribbons indicate 95% credible intervals; the best-fitting distribution is annotated. (B) Comparison of CDFs fitted to all data vs. the filtered dataset for the best-fitting distribution; shown only when filtering removed at least one dataset. (C) Subgroup CDFs for the best-fitting distribution; the overall estimate (shaded ribbon) is shown alongside subgroup-specific estimates; shown only when subgroup analyses were performed.

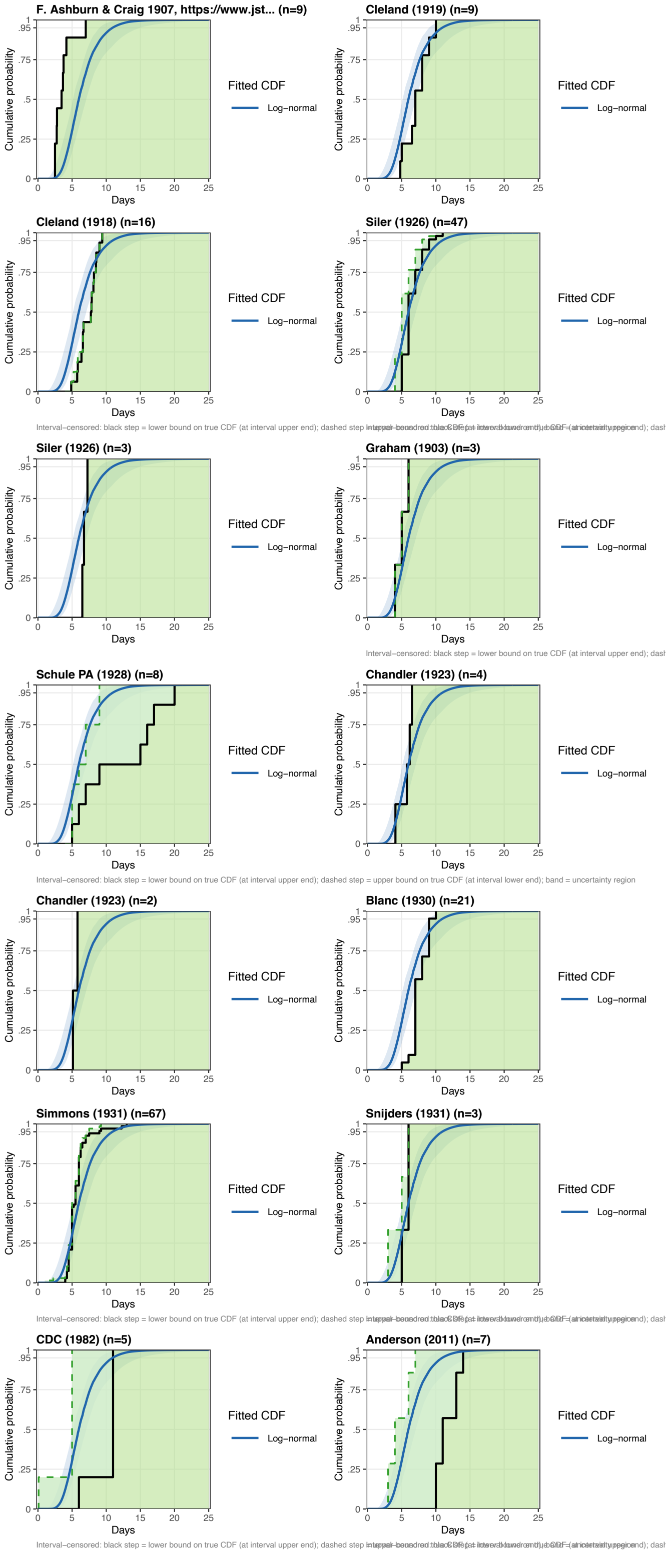

**Supplementary Figure S29. Raw incubation period data for Dengue:** (F) Empirical CDFs for datasets reporting frequency tables or interval-censored observations, overlaid with the posterior predictive CDF for the best-fitting distribution (coloured ribbon and line). For interval-censored data, the solid step line is the conservative ECDF at interval upper bounds; the dashed step line is the ECDF at interval lower bounds; the shaded band represents the uncertainty region.

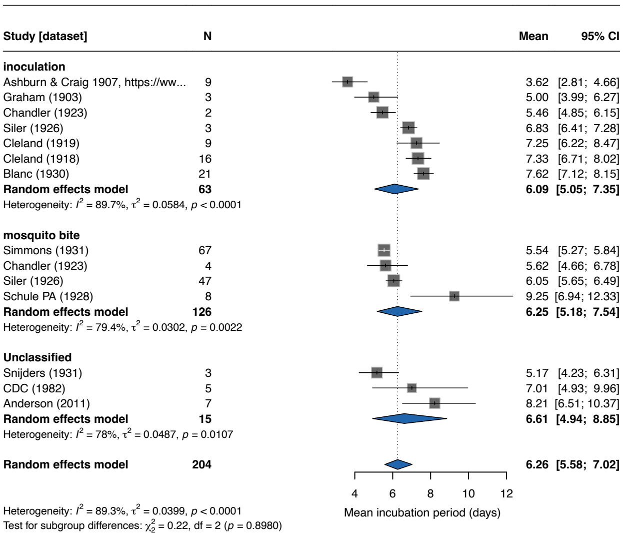

**Supplementary Figure S30. Forest plot of mean incubation period for Dengue:** Squares represent study-specific estimates with 95% confidence intervals (CIs); square size is proportional to study weight. The diamond shows the overall pooled estimate from a random-effects model with log-transformed mean (back-transformed to days). Studies are stratified by exposure route; subgroup pooled estimates are shown as separate diamonds. Heterogeneity is quantified by $I^2$ and $\tau^2$.

##### C.10. Zika

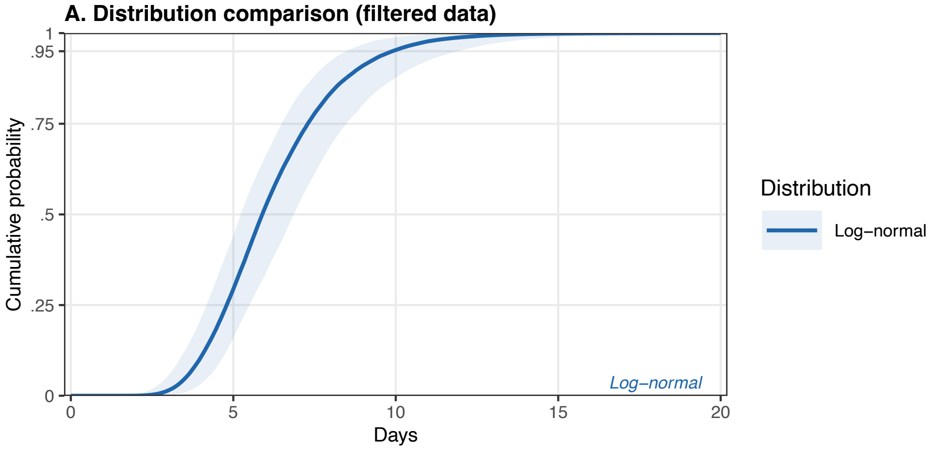

**Supplementary Figure S31. Incubation period model fits for Zika:** (A) Posterior predictive cumulative distribution functions (CDFs) for all converged parametric distributions fitted to the filtered dataset. Ribbons indicate 95% credible intervals; the best-fitting distribution is annotated.

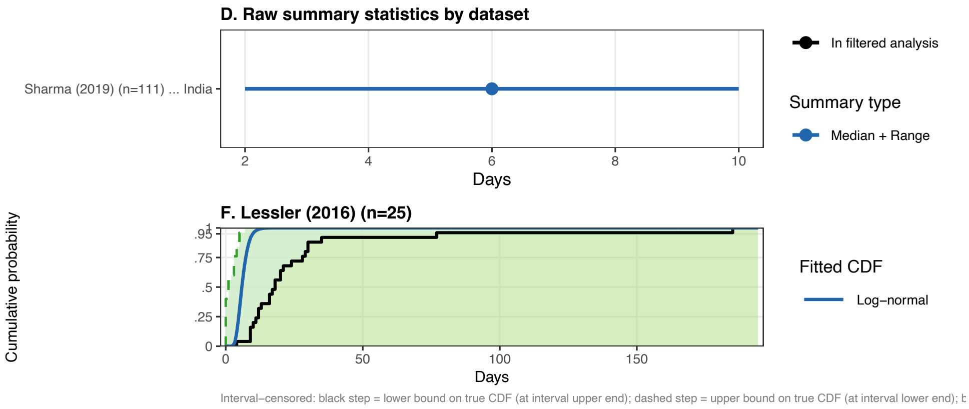

**Supplementary Figure S32. Raw incubation period data for Zika:** (D) Raw summary statistics reported in source datasets. Points indicate the central estimate (circle: median; square: mean) and lines the reported uncertainty interval (solid: range; dashed: IQR; dotted: SD). Faded entries were excluded by the data-quality filter. (F) Empirical CDFs for datasets reporting frequency tables or interval-censored observations, overlaid with the posterior predictive CDF for the best-fitting distribution (coloured ribbon and line). For interval-censored data, the solid step line is the conservative ECDF at interval upper bounds; the dashed step line is the ECDF at interval lower bounds; the shaded band represents the uncertainty region.

##### C.11. Rift Valley fever

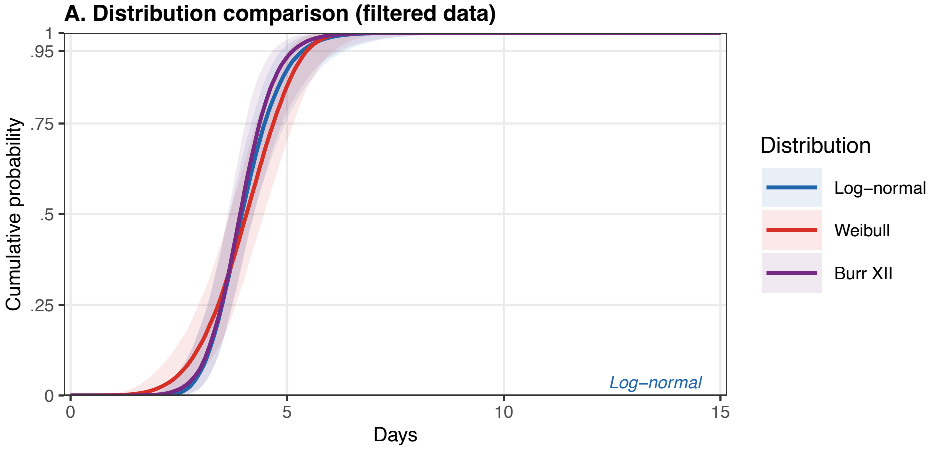

**Supplementary Figure S33. Incubation period model fits for Rift Valley Fever:** (A) Posterior predictive cumulative distribution functions (CDFs) for all converged parametric distributions fitted to the filtered dataset. Ribbons indicate 95% credible intervals; the best-fitting distribution is annotated.

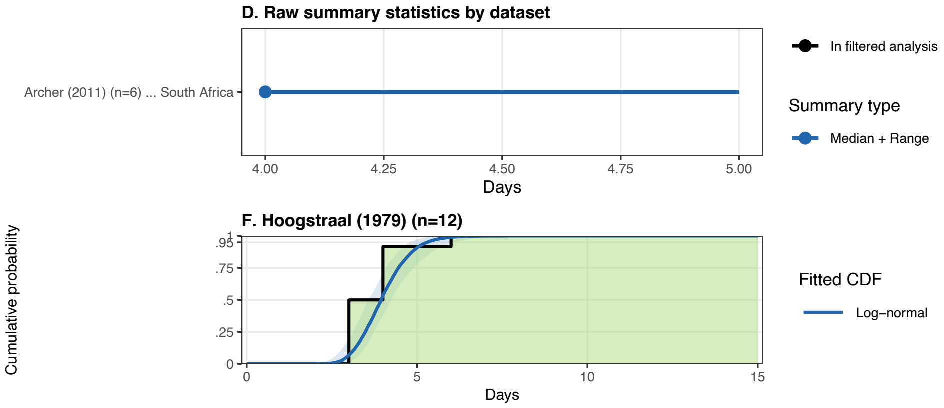

**Supplementary Figure S34. Raw incubation period data for Rift Valley Fever:** (D) Raw summary statistics reported in source datasets. Points indicate the central estimate (circle: median; square: mean) and lines the reported uncertainty interval (solid: range; dashed: IQR; dotted: SD). Faded entries were excluded by the data-quality filter. (F) Empirical CDFs for datasets reporting frequency tables or interval-censored observations, overlaid with the posterior predictive CDF for the best-fitting distribution (coloured ribbon and line). For interval-censored data, the solid step line is the conservative ECDF at interval upper bounds; the dashed step line is the ECDF at interval lower bounds; the shaded band represents the uncertainty region.

###### C.12. Nipah

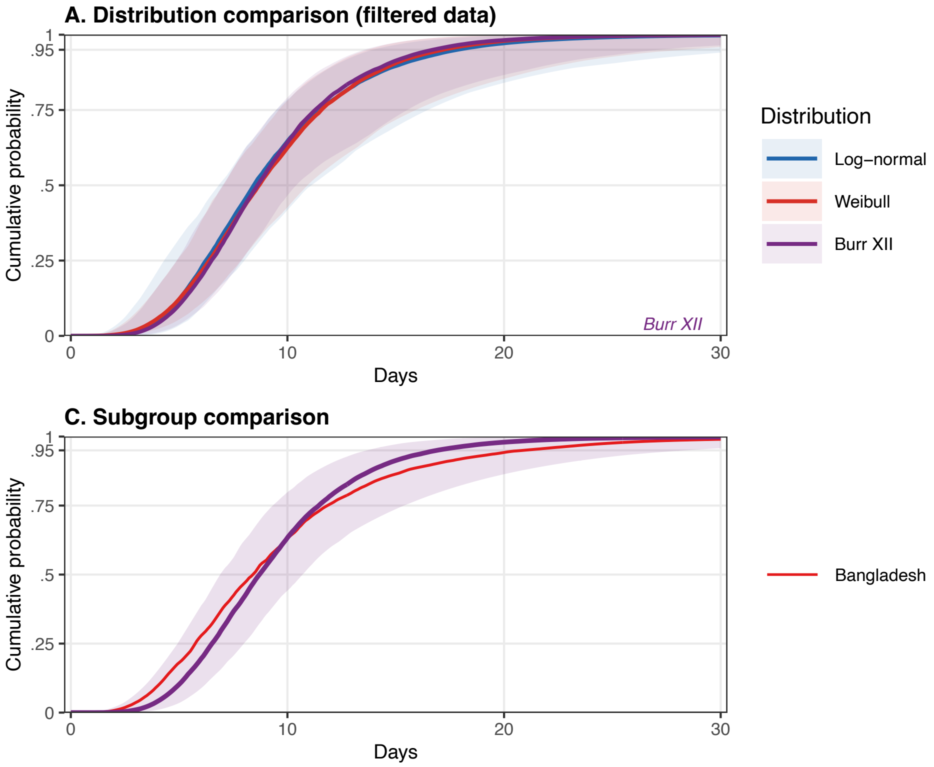

**Supplementary Figure S35. Incubation period model fits for Nipah:** (A) Posterior predictive cumulative distribution functions (CDFs) for all converged parametric distributions fitted to the filtered dataset. Ribbons indicate 95% credible intervals; the best-fitting distribution is annotated. (B) Comparison of CDFs fitted to all data vs. the filtered dataset for the best-fitting distribution; shown only when filtering removed at least one dataset. (C) Subgroup CDFs for the best-fitting distribution; the overall estimate (shaded ribbon) is shown alongside subgroup-specific estimates; shown only when subgroup analyses were performed.

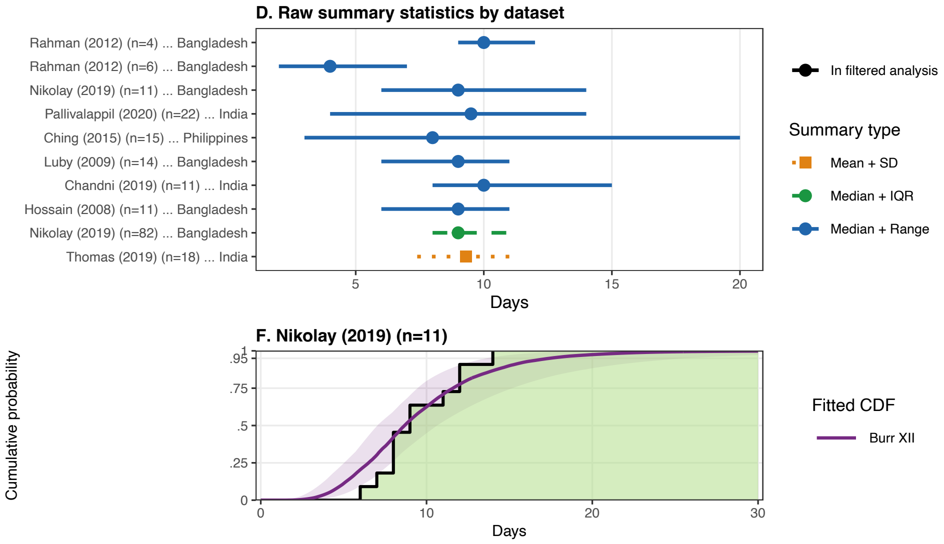

**Supplementary Figure S36. Raw incubation period data for Nipah:** (D) Raw summary statistics reported in source datasets. Points indicate the central estimate (circle: median; square: mean) and lines the reported uncertainty interval (solid: range; dashed: IQR; dotted: SD). Faded entries were excluded by the data-quality filter. (F) Empirical CDFs for datasets reporting frequency tables or interval-censored observations, overlaid with the posterior predictive CDF for the best-fitting distribution (coloured ribbon and line). For interval-censored data, the solid step line is the conservative ECDF at interval upper bounds; the dashed step line is the ECDF at interval lower bounds; the shaded band represents the uncertainty region.

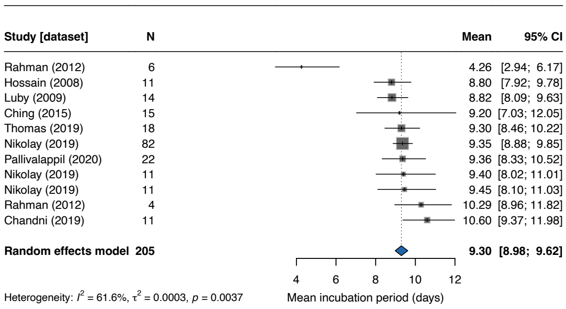

**Supplementary Figure S37. Forest plot of mean incubation period for Nipah:** Squares represent study-specific estimates with 95% confidence intervals (CIs); square size is proportional to study weight. The diamond shows the overall pooled estimate from a random-effects model with log-transformed mean (back-transformed to days). Heterogeneity is quantified by $I^2$ and $\tau^2$.

###### C.13. Mpox

We note that the inferred incubation period distribution is based on a single (global) outbreak of mpox in 2022. Most data is derived from a specific population group (MSM) and may not be representative for other settings and clades.

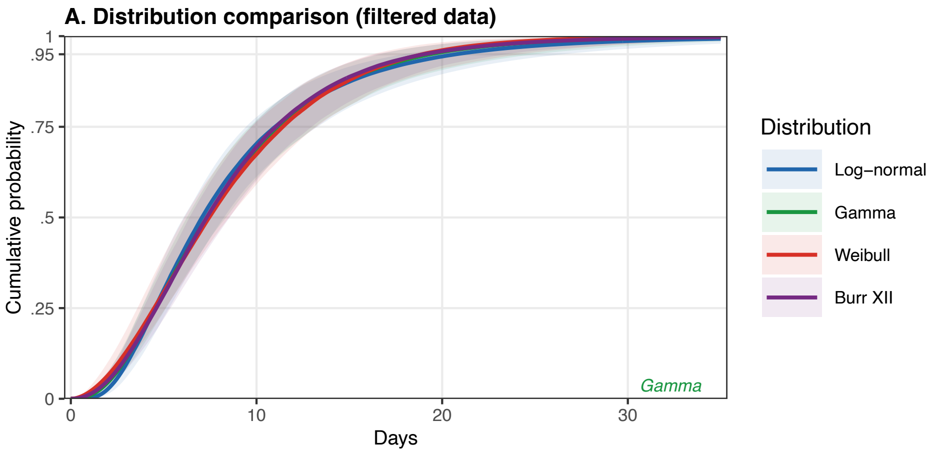

**Supplementary Figure S38. Incubation period model fits for mpox:** (A) Posterior predictive cumulative distribution functions (CDFs) for all converged parametric distributions fitted to the filtered dataset. Ribbons indicate 95% credible intervals; the best-fitting distribution is annotated.

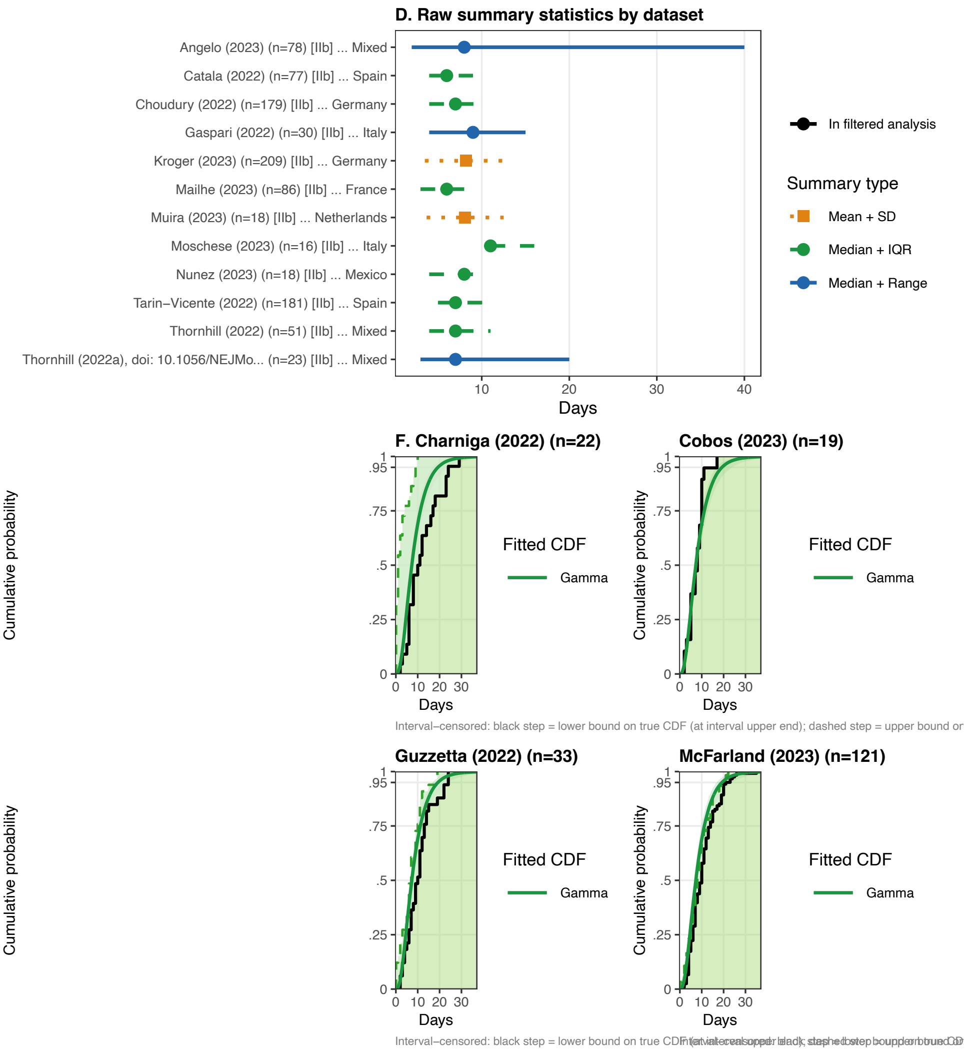

**Supplementary Figure S39. Raw incubation period data for mpox:** (D) Raw summary statistics reported in source datasets. Points indicate the central estimate (circle: median; square: mean) and lines the reported uncertainty interval (solid: range; dashed: IQR; dotted: SD). Faded entries were excluded by the data-quality filter. (F) Empirical CDFs for datasets reporting frequency tables or interval-censored observations, overlaid with the posterior predictive CDF for the best-fitting distribution (coloured ribbon and line). For interval-censored data, the solid step line is the conservative ECDF at interval upper bounds; the dashed step line is the ECDF at interval lower bounds; the shaded band represents the uncertainty region.

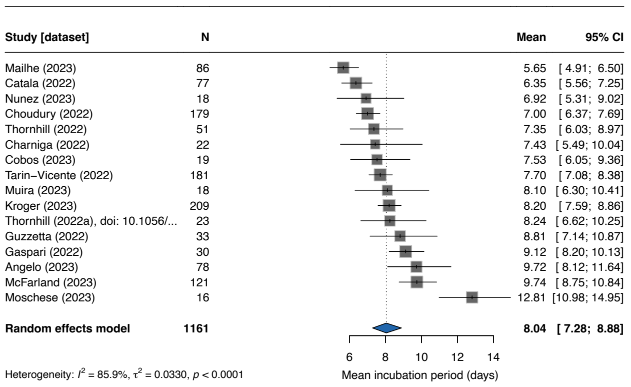

**Supplementary Figure S40. Forest plot of mean incubation period for mpox:** Squares represent study-specific estimates with 95% confidence intervals (CIs); square size is proportional to study weight. The diamond shows the overall pooled estimate from a random-effects model with log-transformed mean (back-transformed to days). Heterogeneity is quantified by $I^2$ and $\tau^2$.

#### C.14. Measles

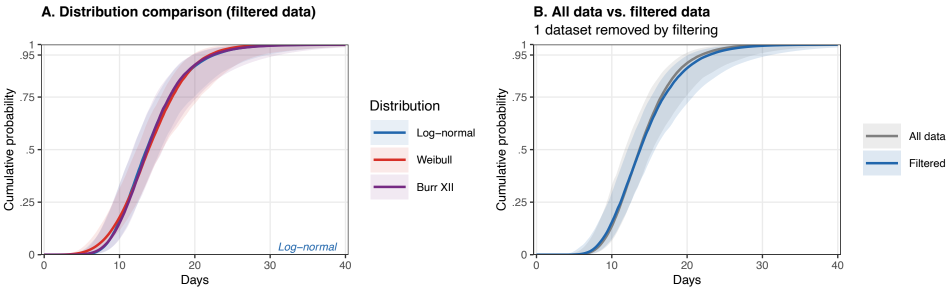

**Supplementary Figure S41. Incubation period model fits for Measles:** (A) Posterior predictive cumulative distribution functions (CDFs) for all converged parametric distributions fitted to the filtered dataset. Ribbons indicate 95% credible intervals; the best-fitting distribution is annotated. (B) Comparison of CDFs fitted to all data vs. the filtered dataset for the best-fitting distribution; shown only when filtering removed at least one dataset. (C) Subgroup CDFs for the best-fitting distribution; the overall estimate (shaded ribbon) is shown alongside subgroup-specific estimates; shown only when subgroup analyses were performed.

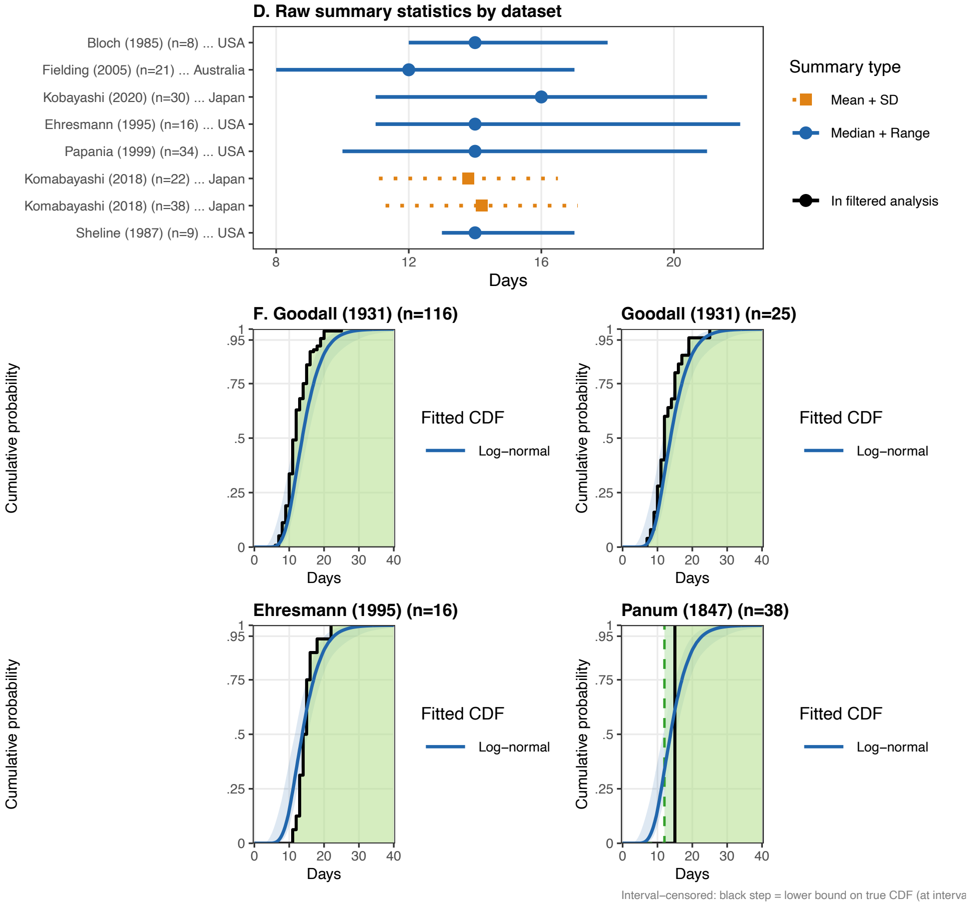

**Supplementary Figure S42. Raw incubation period data for Measles:** (D) Raw summary statistics reported in source datasets. Points indicate the central estimate (circle: median; square: mean) and lines the reported uncertainty interval (solid: range; dashed: IQR; dotted: SD). Faded entries were excluded by the data-quality filter. (F) Empirical CDFs for datasets reporting frequency tables or interval-censored observations, overlaid with the posterior predictive CDF for the best-fitting distribution (coloured ribbon and line). For interval-censored data, the solid step line is the conservative ECDF at interval upper bounds; the dashed step line is the ECDF at interval lower bounds; the shaded band represents the uncertainty region.

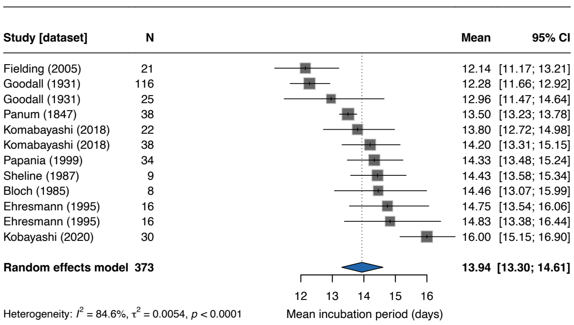

**Supplementary Figure S43. Forest plot of mean incubation period for Measles:** Squares represent study-specific estimates with 95% confidence intervals (CIs); square size is proportional to study weight. The diamond shows the overall pooled estimate from a random-effects model with log-transformed mean (back-transformed to days). Heterogeneity is quantified by $I^2$ and $\tau^2$.

**C.15. Smallpox**

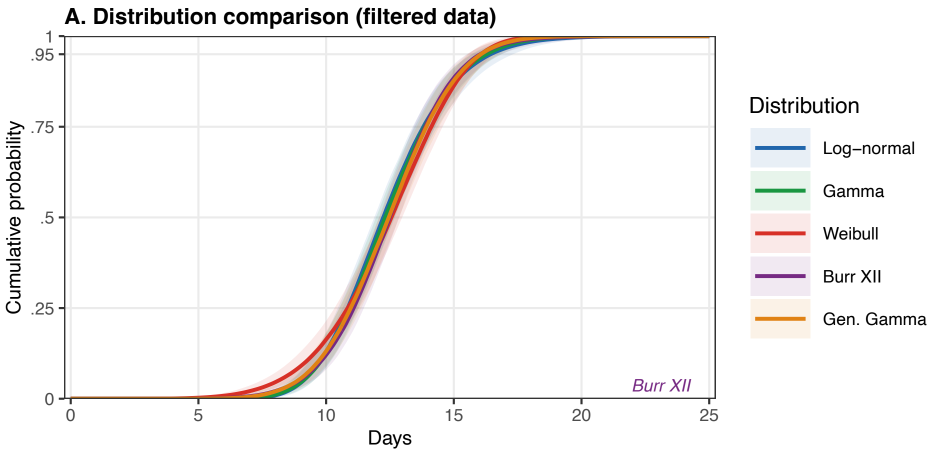

**Supplementary Figure S44. Incubation period model fits for Smallpox:** (A) Posterior predictive cumulative distribution functions (CDFs) for all converged parametric distributions fitted to the filtered dataset. Ribbons indicate 95% credible intervals; the best-fitting distribution is annotated.

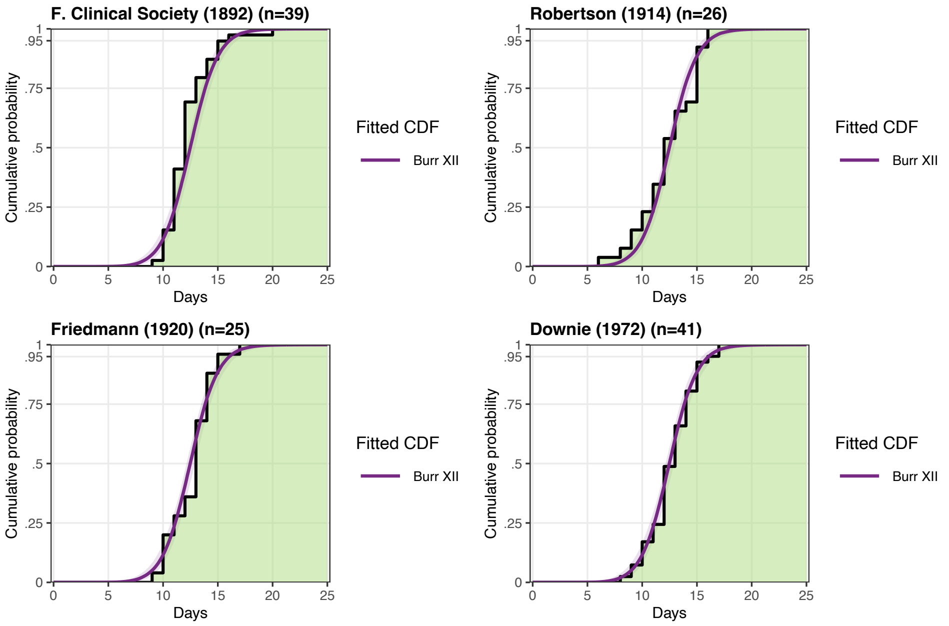

**Supplementary Figure S45. Raw incubation period data for Smallpox:** (F) Empirical CDFs for datasets reporting frequency tables or interval-censored observations, overlaid with the posterior predictive CDF for the best-fitting distribution (coloured ribbon and line). For interval-censored data, the solid step line is the conservative ECDF at interval upper bounds; the dashed step line is the ECDF at interval lower bounds; the shaded band represents the uncertainty region.

##### C.16. Cholera

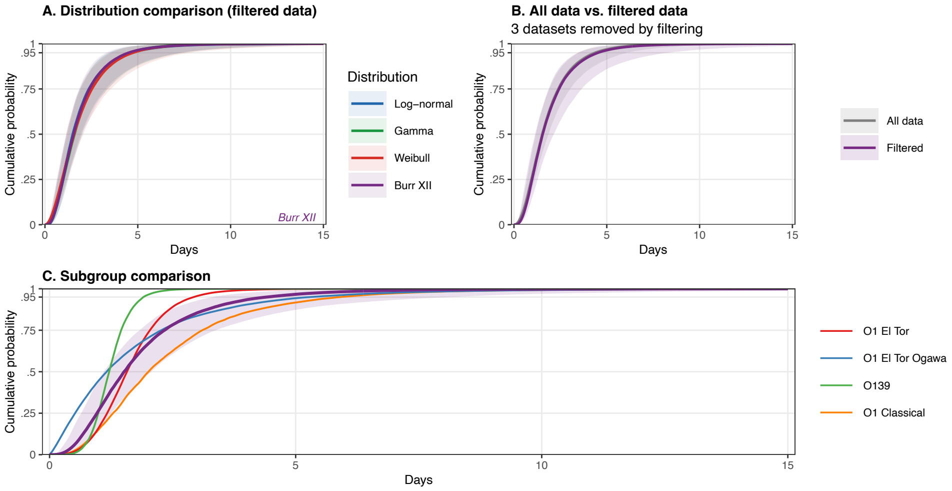

**Supplementary Figure S46. Incubation period model fits for Cholera:** (A) Posterior predictive cumulative distribution functions (CDFs) for all converged parametric distributions fitted to the filtered dataset. Ribbons indicate 95% credible intervals; the best-fitting distribution is annotated. (B) Comparison of CDFs fitted to all data vs. the filtered dataset for the best-fitting distribution; shown only when filtering removed at least one dataset. (C) Subgroup CDFs for the best-fitting distribution; the overall estimate (shaded ribbon) is shown alongside subgroup-specific estimates; shown only when subgroup analyses were performed.

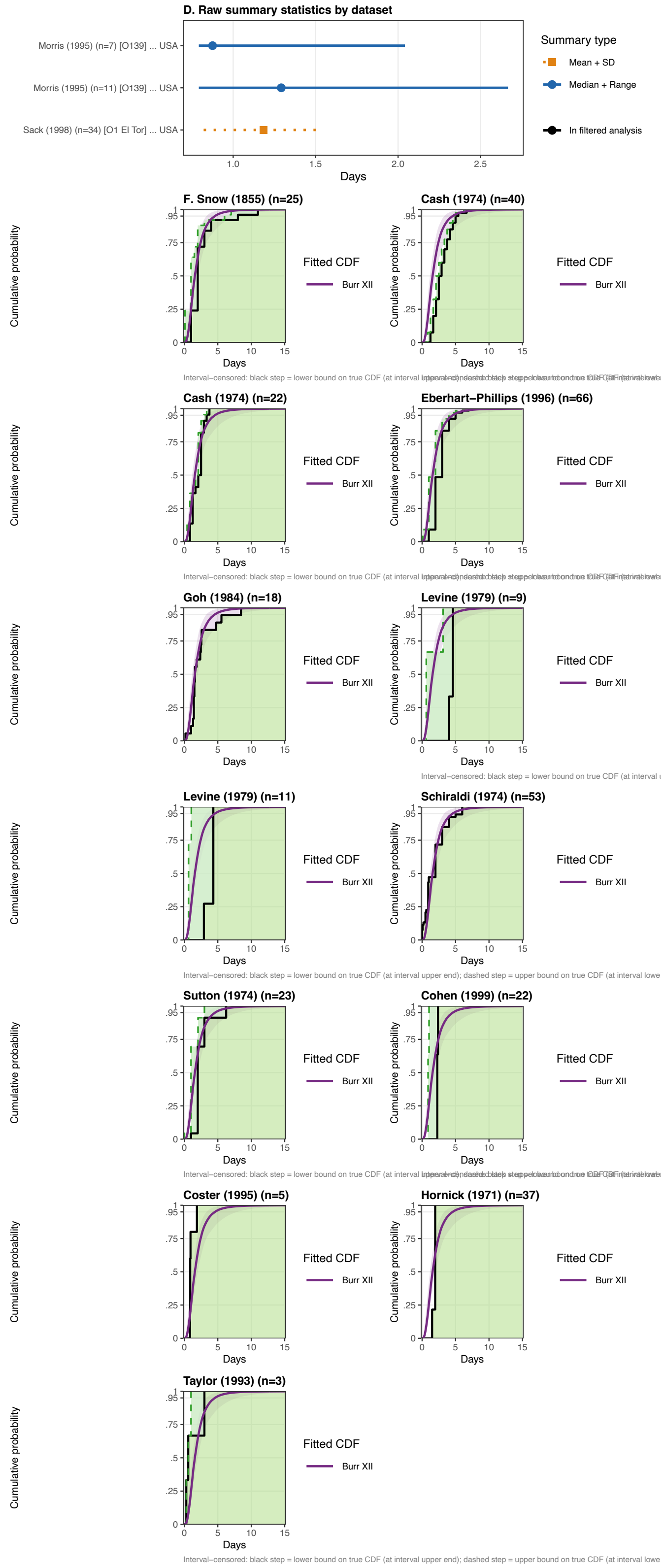

**Supplementary Figure S47. Raw incubation period data for Cholera:** (D) Raw summary statistics reported in source datasets. Points indicate the central estimate (circle: median; square: mean) and lines the reported uncertainty interval (solid: range; dashed: IQR; dotted: SD). Faded entries were excluded by the data-quality filter. (F) Empirical CDFs for datasets reporting frequency tables or interval-censored observations, overlaid with the posterior predictive CDF for the best-fitting distribution (coloured ribbon and line). For interval-censored data, the solid step line is the conservative ECDF at interval upper bounds; the dashed step line is the ECDF at interval lower bounds; the shaded band represents the uncertainty region.

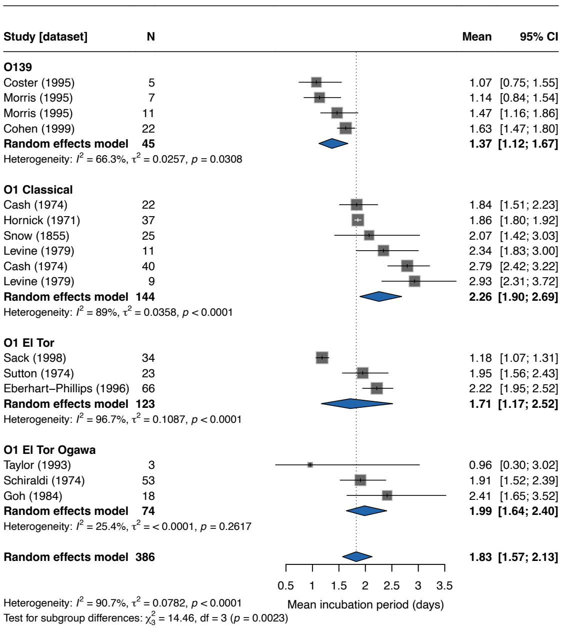

**Supplementary Figure S48. Forest plot of mean incubation period for Cholera:** Squares represent study-specific estimates with 95% confidence intervals (CIs); square size is proportional to study weight. The diamond shows the overall pooled estimate from a random-effects model with log-transformed mean (back-transformed to days). Studies are stratified by biotype; subgroup pooled estimates are shown as separate diamonds. Heterogeneity is quantified by $I^2$ and $\tau^2$.

##### C.17. Typhoid

Typhoid fever is an excellent example of the impact of dose on the incubation period, which we demonstrate with the subgroup analysis by exposure route.

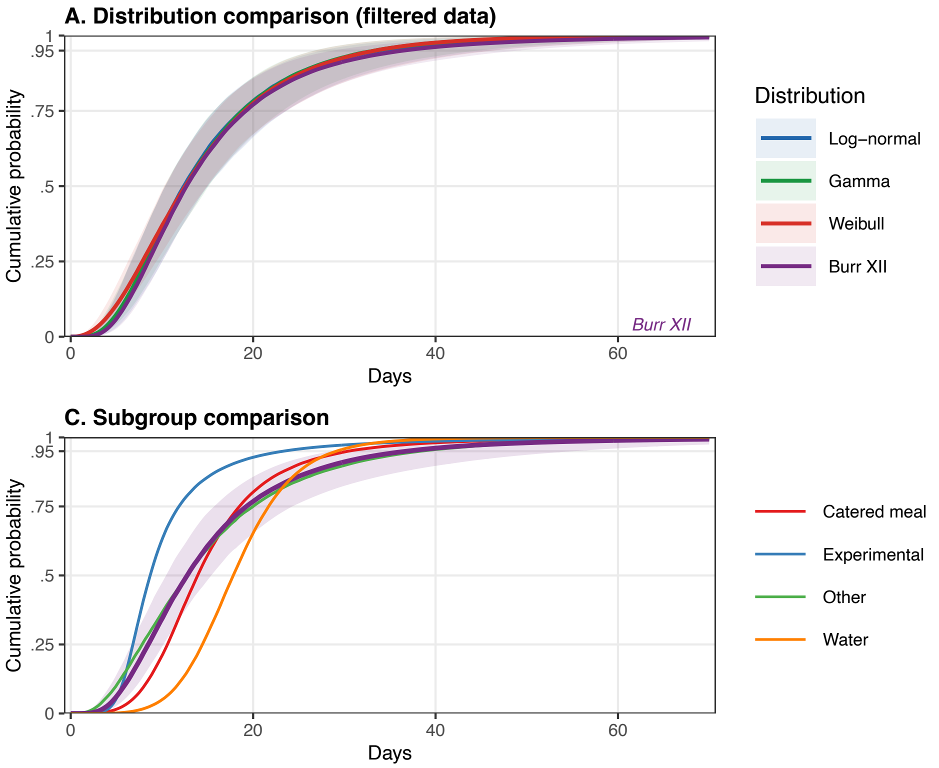

**Supplementary Figure S49. Incubation period model fits for Typhoid:** (A) Posterior predictive cumulative distribution functions (CDFs) for all converged parametric distributions fitted to the filtered dataset. Ribbons indicate 95% credible intervals; the best-fitting distribution is annotated. (B) Comparison of CDFs fitted to all data vs. the filtered dataset for the best-fitting distribution; shown only when filtering removed at least one dataset. (C) Subgroup CDFs for the best-fitting distribution; the overall estimate (shaded ribbon) is shown alongside subgroup-specific estimates; shown only when subgroup analyses were performed.

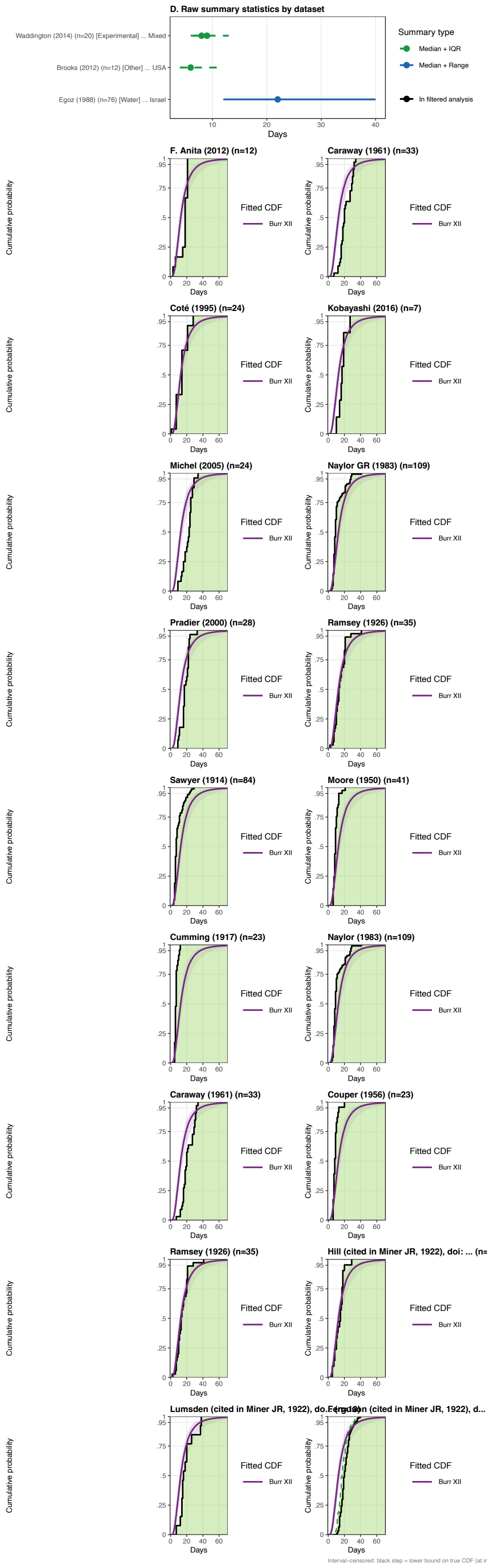

**Supplementary Figure S50. Raw incubation period data for Typhoid:** (D) Raw summary statistics reported in source datasets. Points indicate the central estimate (circle: median; square: mean) and lines the reported uncertainty interval (solid: range; dashed: IQR; dotted: SD). Faded entries were excluded by the data-quality filter. (F) Empirical CDFs for datasets reporting frequency tables or interval-censored observations, overlaid with the posterior predictive CDF for the best-fitting distribution (coloured ribbon and line). For interval-censored data, the solid step line is the conservative ECDF at interval upper bounds; the dashed step line is the ECDF at interval lower bounds; the shaded band represents the uncertainty region.

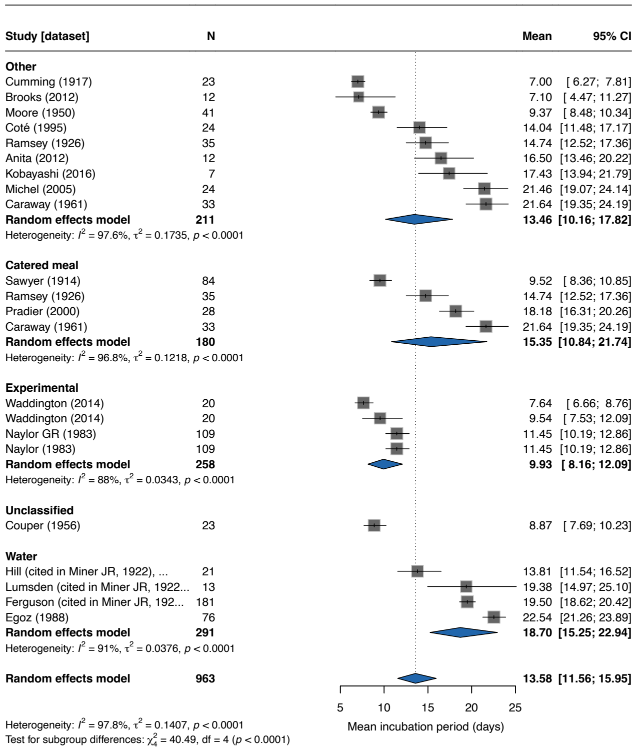

**Supplementary Figure S51. Forest plot of mean incubation period for Typhoid:** Squares represent study-specific estimates with 95% confidence intervals (CIs); square size is proportional to study weight. The diamond shows the overall pooled estimate from a random-effects model with log-transformed mean (back-transformed to days). Studies are stratified by exposure route; subgroup pooled estimates are shown as separate diamonds. Heterogeneity is quantified by $I^2$ and $\tau^2$.

#### C.18. Yellow fever

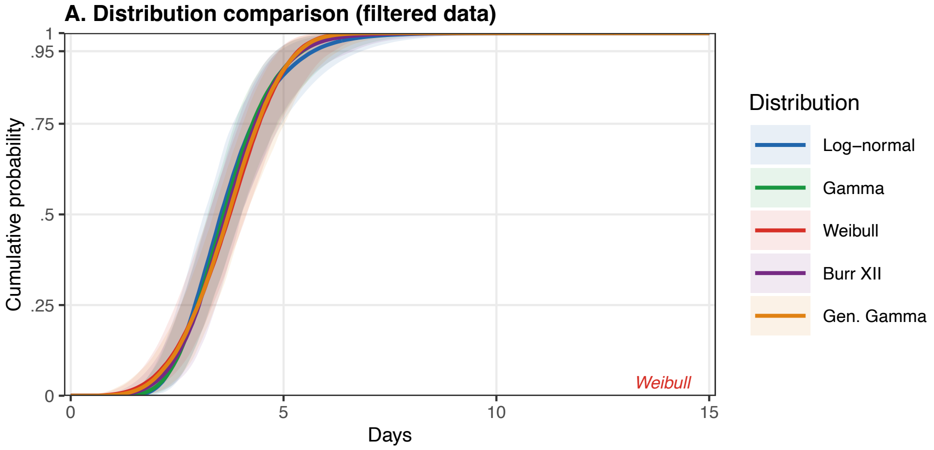

**Supplementary Figure S52. Incubation period model fits for Yellow fever:** (A) Posterior predictive cumulative distribution functions (CDFs) for all converged parametric distributions fitted to the filtered dataset. Ribbons indicate 95% credible intervals; the best-fitting distribution is annotated.

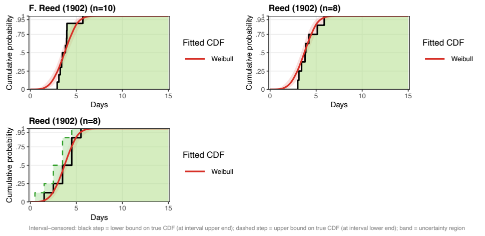

**Supplementary Figure S53. Raw incubation period data for Yellow fever:** (F) Empirical CDFs for datasets reporting frequency tables or interval-censored observations, overlaid with the posterior predictive CDF for the best-fitting distribution (coloured ribbon and line). For interval-censored data, the solid step line is the conservative ECDF at interval upper bounds; the dashed step line is the ECDF at interval lower bounds; the shaded band represents the uncertainty region.
